# Improved functional mapping with GSA-MiXeR implicates biologically specific gene-sets and estimates enrichment magnitude

**DOI:** 10.1101/2022.12.08.22283159

**Authors:** Oleksandr Frei, Guy Hindley, Alexey A. Shadrin, Dennis van der Meer, Bayram C. Akdeniz, Weiqiu Cheng, Kevin S. O’Connell, Shahram Bahrami, Nadine Parker, Olav B. Smeland, Dominic Holland, Schizophrenia Working Group of the Psychiatric Genomics Consortium, Christiaan de Leeuw, Danielle Posthuma, Ole A. Andreassen, Anders M. Dale

**Author notes:** Corresponding author: Oleksandr Frei.

## Abstract

While genome-wide association studies (GWAS) are increasingly successful in discovering genomic loci associated with complex human traits and disorders, the biological interpretation of these findings remains challenging. We developed the GSA-MiXeR analytical tool for gene-set analysis (GSA), which fits a model for gene-set heritability enrichments for complex traits, accounting for linkage disequilibrium across variants, and allowing the quantification of partitioned heritability and fold enrichment for small gene-sets. We validate the method using extensive simulations and sensitivity analyses. When applied to height and schizophrenia, GSA-MiXeR implicates gene-sets with greater biological specificity compared to standard GSA approaches, including insulin-like growth factor for height, as well as calcium channel function, GABAergic and dopaminergic signaling for schizophrenia. Such biologically relevant gene-sets, often with less than ten genes, are more likely to provide new insights into the pathobiology of complex diseases and highlight potential drug targets.

## Introduction

Genome-wide association studies (GWAS) have discovered thousands of genomic loci associated with complex human traits and disorders, highlighting their polygenic nature and the predominance of small individual effects of common genetic variants^1^. Gene-set analysis (GSA) has become a powerful tool for understanding biological implications of GWAS findings^2^. Building upon large databases of gene-sets such as the Molecular Signatures Database (MSigDB)^3^, Gene Ontology (GO)^4^, Synaptic Gene Ontologies (SynGO)^5^ and others, GSA has provided many prominent findings^6-8^, implicating the role of biological pathways and yielding relevant tissue- and cell type-specific insights into complex human traits and disorders.

The most popular analytical tools for GSA analysis of GWAS data are MAGMA^9^, Fisher’s exact (hypergeometric) test^10^, and the stratified linkage disequilibrium (LD) score regression (sLDSC)^11^. MAGMA implements a two-stage approach, where GWAS p-values of single nucleotide polymorphisms (SNPs) are first aggregated into gene-level p-values, which are further tested for association with predefined gene-sets. The hypergeometric test evaluates whether the genes implicated in GWAS is over-represented within predefined gene-sets. Both tools implement competitive GSA^12^, testing the null hypothesis that the genes in question are no more strongly associated with the phenotype than other genes. A limitation to both approaches is that they only provide a measure of statistical significance (“enrichment p-value”), which largely depends both on the sample size of the GWAS, and on the size of a gene-set. Such enrichment p-values do not constitute a biologically meaningful measure of enrichment magnitude, limiting our ability to compare the relative strength of enrichment across or within traits. Importantly, enrichment p-values correlate with gene-set size, tending to be more significant for larger gene-sets, thus limiting our ability to discover more informative biological pathways, with greater specificity to a given phenotype.

The sLDSC method quantifies enrichment by partitioning heritability across genomic regions. Specifically, such partitioning allows one to compute a trait’s “fold enrichment” score for a genomic region. This is given by the ratio of the trait’s heritability attributed to the region (as estimated by the model) to the heritability of the region estimated from a baseline model which assumes that heritability is uniformly distributed across the genome. The sLDSC method uses GWAS summary statistics to quantify fold enrichments in functional categories. For example, regions conserved in mammals have a disproportionately large contribution to heritability for many traits and disorders^11^. However, for smaller annotations it had been shown that block jackknife-based significance testing used in sLDSC does not always control type 1 error for annotations with approximately 0.5% of SNPs or less^13^. Furthermore, sLDSC is based on an infinitesimal model with additional simplifying assumptions about the distribution of genetic effects with respect to minor allele frequency (MAF) and LD^14^, which in certain cases has been shown to bias fold enrichment estimates^15^. Finally, the sLDSC method has so far been only applied to tissue- and cell type-specific annotations, but not to gene ontologies, both due to computational challenges (such as excessive memory usage and runtime) and algorithmic failures (such as non-invertible matrices arising in block jackknife-based significance testing) which prevent application of sLDSC tool to enrichment analysis of gene ontologies.

Here, building on previously developed MiXeR framework^16-19^, we introduce GSA-MiXeR, a novel method for competitive GSA which quantifies partitioned heritability attributed to each gene-set and its fold enrichment with respect to the baseline model (see Online Methods). Efficient design and implementation, enabled by stochastic gradient-based log-likelihood optimization^20^, allow GSA-MiXeR to jointly model more than 18,000 genes in a single model, while accounting simultaneously for the trait’s polygenicity, MAF- and LD-dependency of genetic effects, and differential enrichment of functional categories, thus taking into account the unique genetic architecture of each trait. We evaluated the accuracy of GSA-MiXeR’s fold enrichment estimates in extensive simulations using real UK Biobank genotypes. We also performed replication analysis using independent GWAS for discovery and replication, which confirmed that ranking gene-sets according to fold enrichment (GSA-MiXeR estimate) results in a more stable order of gene-sets compared to ranking based on enrichment p-values from MAGMA, even though GSA-MiXeR promoted smaller gene-sets of potentially higher biological relevance. We also performed extensive sensitivity analyses, testing robustness of the GSA-MiXeR method against a mis-specified model of the genetic architecture and out-of-sample LD information.

To demonstrate GSA-MiXeR performance with real phenotypic data, we applied it to height and schizophrenia, two highly polygenic phenotypes^21^. In our main analysis, we report GSA-MiXeR results for significant gene-sets first identified by MAGMA. This shows how the addition of fold enrichments enhances the biological interpretation of GSA results, allowing comparisons of gene-set effect size within and across phenotypes. Application to height demonstrates GSA-MiXeR’s ability to reorder gene-sets in a way that promotes smaller, and potentially more biologically relevant gene-sets, in particular highlighting insulin-like growth factor pathways, which have an established role in the biology of height^22^. In schizophrenia, GSA-MiXeR reveals that gene-sets related to calcium channel function have greater fold enrichment than larger gene-sets related to post-synaptic functioning. In a subsequent exploratory analysis of schizophrenia without filtering on significant MAGMA associations, we demonstrate that GSA-MiXeR implicates other highly enriched and biologically plausible gene-sets, including biological processes related to dopaminergic and GABAergic neurons, pointing to potential key biological pathways perturbed in schizophrenia, and supporting prior hypotheses of schizophrenia pathogenesis^23^.

## Results

### Simulation studies

To evaluate the accuracy of fold enrichment estimates from GSA-MiXeR, we conducted simulations by synthesizing a quantitative trait with pre-defined SNP-heritability (h2=0.1, 0.4 or 0.7) fully attributed to the genic regions using real UK Biobank genotypes from N=337,145 subjects after quality control (QC). In simulations without enrichment, all genetic effects were randomly distributed across the genic regions. Then, to simulate enrichment, a certain number of genes (n=1, 10, or 100) received a 3, 10 or 30-fold enrichment in heritability attributed to those genes. We used boundaries of real protein-coding and pseudo-genes in these simulations and selected an enriched subset of genes at random. Then we fitted two models: the baseline model with two variance parameters, allowing for different effect size variance in genic regions versus non-genic regions; and the full model, where, in addition to the baseline model, each gene was allowed to have its own effect size variance. Estimated fold enrichment of a gene-set was calculated as a ratio of its heritability estimate from the full model versus the baseline model (see Online Methods).

The results (Figure 1, Supplementary Figure 1 and Supplementary Table 1) show that GSA-MiXeR model provides an unbiased estimate of fold enrichments in scenarios where GWAS summary statistics have sufficiently strong signal (h2=0.4 and h2=0.7). Notably, this also applies to gene-sets which comprise only one gene (N=1 scenario). The results for a weaker GWAS signal (h2=0.1) indicate that the estimates of fold enrichment are biased downwards, underestimating real enrichment by up to 50%. The scenario without enrichment had no inflation in enrichment estimates, with an average fold enrichment close to 1.0.

**1.**
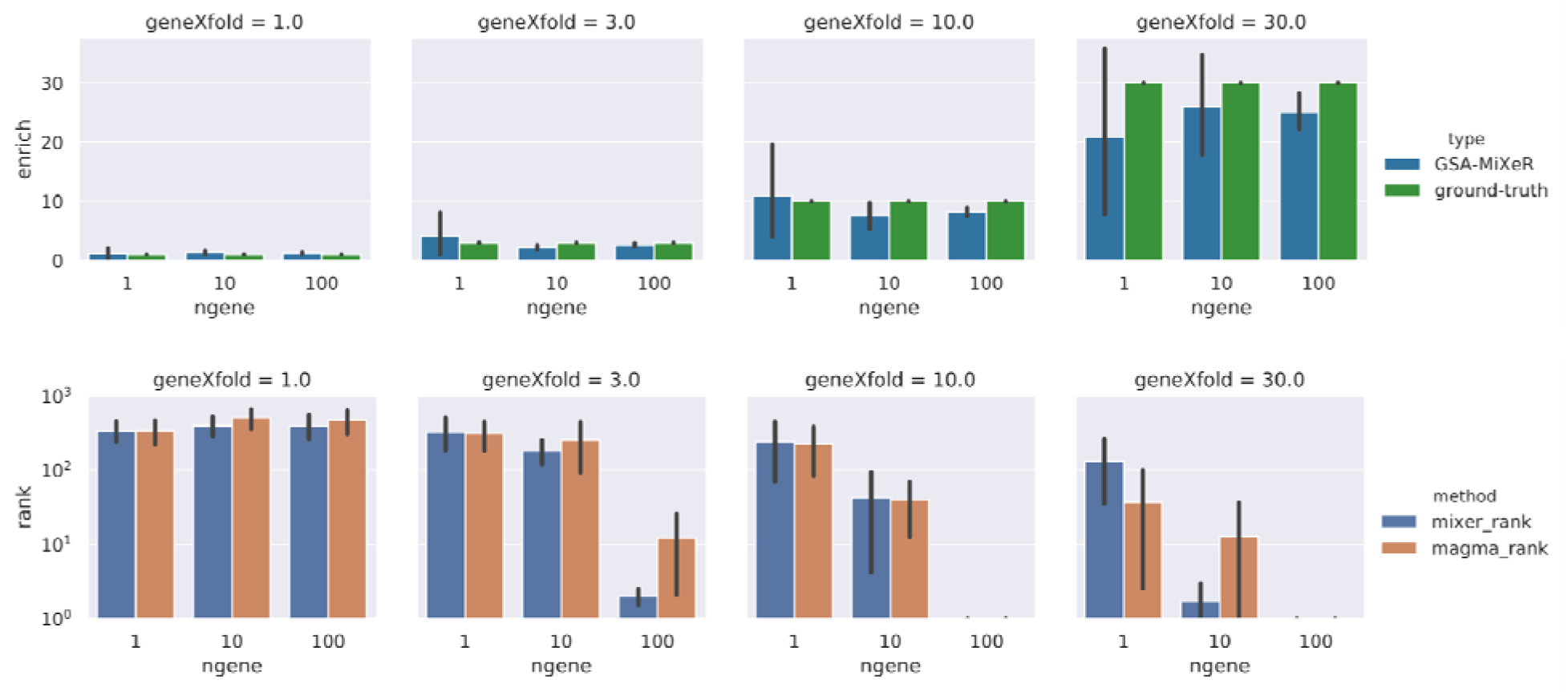
Selected results from simulations. The top row of figures compares fold enrichment estimated by GSA-MiXeR versus ground truth. The bottom row of figures visualizes accuracy of sorting gene-sets according to the MAGMA enrichment p-values, or according to the GSA-MiXeR fold enrichment, showing the position of the truly enriched gene-set among 1000 randomly generated gene-sets in simulations (so tat expected rank is 1 for scenarios with enrichment). The simulations cover four levels of enrichment: 1 (null enrichment), and 3, 10 and 30-fold enrichment; and vary in the size of enriched gene-set: 1, 10 and 100 genes. In all simulations the trait was simulated with SNP h2 of 40%. An extended set of simulation results are available in Supplementary Table 1 and 2.

We performed additional simulations to compare GSA-MiXeR and MAGMA’s ability to sort gene-sets in a way that promotes enriched gene-sets. For each of the previously conducted simulations we generated a further 999 gene-sets, each with the same number of genes as the enriched gene-set. We then computed the position of the one truly enriched gene-set on the list, sorted either according to GSA-MiXeR fold enrichment or according to MAGMA p-values. The results show that GSA-MiXeR generally outperforms MAGMA for gene-sets with N=10 and N=100 genes (Figure 1, Supplementary Table 2). The difference between the two methods was most prominent for smaller gene-sets (N=10 genes). For instance, in simulations with 30x fold enrichment and h2=0.7, the truly enriched gene-set was on average ranked on 10^th^ position by GSA-MiXeR, and on 146^th^ position according to MAGMA. Nevertheless, for the individual genes (gene-sets with N=1 gene), MiXeR only ranked them at approximately the 100-th position, indicating that the error of estimating gene-level fold enrichment currently appears to be too large to make reliable inferences at the level of individual genes.

### Application to real phenotypic data

We applied GSA-MiXeR to GWAS of height^24^ and schizophrenia^25^, two human phenotypes with distinct genetic architectures and biological underpinnings. In our main analysis, we first identified all enriched gene-sets with MAGMA p-values below 0.05 after Bonferroni correction. Next, we re-ranked gene-sets based on GSA-MiXeR fold enrichment and compared it to the original MAGMA p-value-based ranking. We also compared fold enrichment and partitioned heritability estimated by GSA-MiXeR within and across the phenotypes, and examined gene-level enrichments with a particular focus on schizophrenia.

When applied to height, GSA-MiXeR ranking promoted smaller gene-sets (Figure 2, Supplementary Figure 2) as compared to MAGMA results. The top ten GSA-MiXeR gene-sets had a median size of 43 genes, with an average fold enrichment of 4.25 (Supplementary Table 3); the top ten MAGMA gene-sets had a median size of 1164 genes with an average fold enrichment of 2.28 (Supplementary Table 4). The average fraction of SNP-heritability attributed to a given gene-set was 2.0% among the top-ten GSA-MiXeR gene-sets and 9.1% among the top ten MAGMA gene-sets. Despite capturing a smaller fraction of heritability, the top ten GSA-MiXeR gene-sets exhibited greater granularity, promoting to the top of the list gene-sets related to insulin-like growth factor, bone mineralization, and musculoskeletal development. In contrast, six of the top ten MAGMA gene-sets were related to non-specific basic cellular processes such as regulation of cellular biosynthetic processes.

**2.**
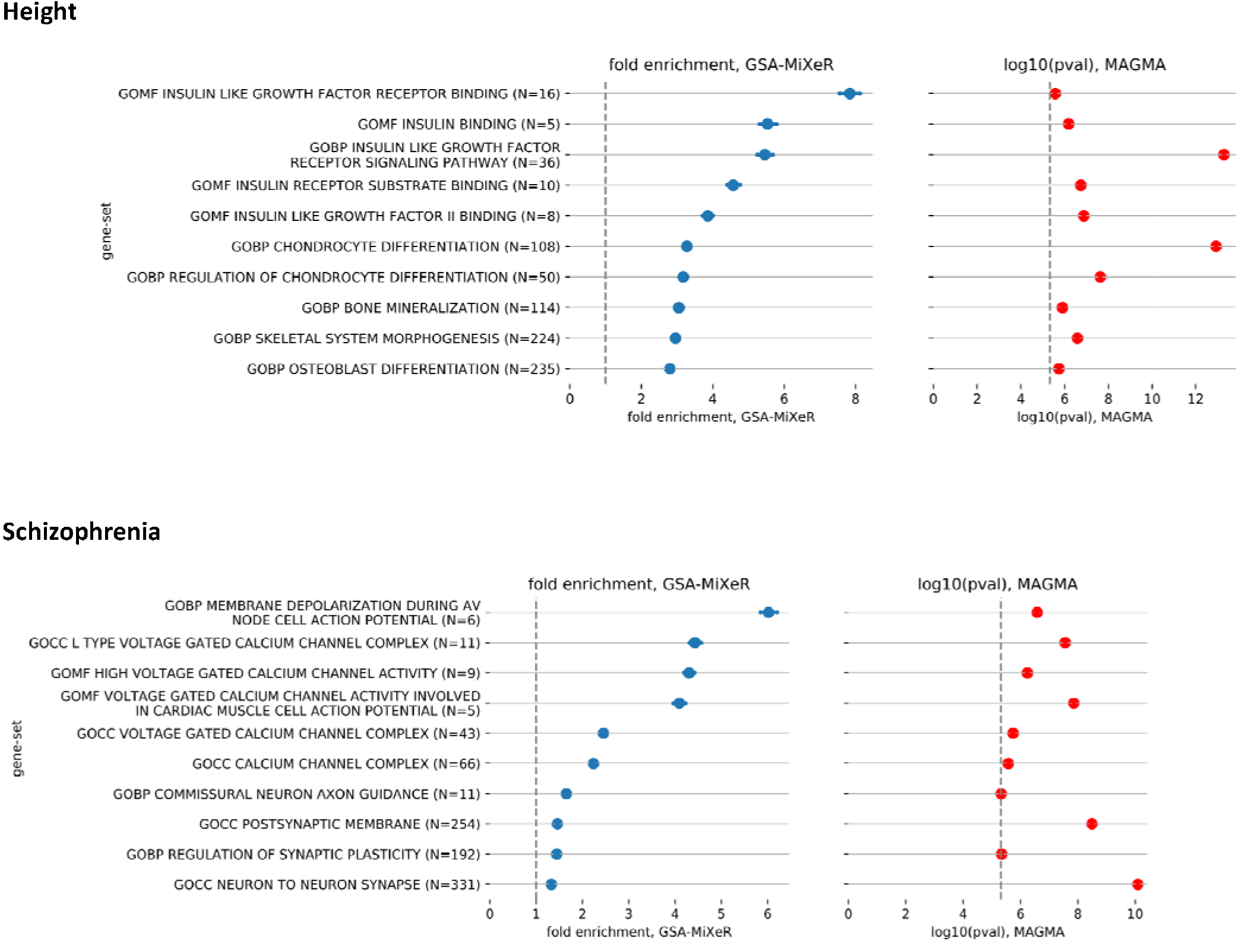
Top-10 gene-sets associated with height and schizophrenia ordered by fold enrichment. Top 10 gene-sets associated with height (top panel) and schizophrenia (bottom panel) according to GSA-MiXeR fold enrichment, filtered on significant MAGMA associations. Left-hand panels show GSA-MiXeR fold enrichment estimates, with standard deviation shown as error bars. Right-hand panels show log10 of MAGMA’s enrichment p-value. The size of gene-sets is indicated in parenthesis after gene-set name. Vertical dashed line indicate no enrichment (for GSA-MiXeR figures) and Bonferroni -corrected p-value threshold (for MAGMA figures). Full results without constraining to top 10 gene-sets are shown in Supplementary Figures 2, 3.

When applied to schizophrenia, the top ten GSA-MiXeR gene-sets had a median size of 27 genes with an average fold enrichment of 2.94 (Supplementary Table 5), versus a median size of 581 genes and an average fold enrichment of 1.85 for the top ten MAGMA gene-sets (Supplementary Table 6). Six out of the top ten GSA-MiXeR gene-sets for schizophrenia were related to voltage-gated calcium channels and membrane depolarization (Figure 1, Supplementary Figure 3), which are more specific biological processes compared to the synaptic and post-synaptic gene-sets prioritized by MAGMA. For the voltage-gated calcium channels gene-sets, the larger enrichment was partly attributable to the *CACNA1C* gene which was the most highly enriched gene in all related gene-sets. In a leave-one-gene-out analysis we show that after exclusion of *CACNA1C* gene the average fold enrichment across those gene sets reduces from 4.71 to 2.67 (Supplementary Table 5), however the gene-sets remain enriched.

In an exploratory analysis of schizophrenia with GSA-MiXeR we considered all gene-sets without filtering on significant MAGMA associations. The top ten gene-sets implicated by the exploratory analysis had a median size of 7 genes with an average fold enrichment of 18.0 (Supplementary Table 7). Interestingly, the top two most fold enriched gene-sets were both related to the dopaminergic system, the main pharmacological target in schizophrenia treatment^26^, which were not significant in the MAGMA analysis. There were also several other gene-sets within the top ten which were related to systems hypothesized to play a role in schizophrenia pathogenesis, including cerebrocortical GABAergic interneuron migration^27^, and interneuron migration^28^. However, as shown by leave-one-gene-out analysis (Supplementary Table 7), the enrichment in these gene-sets appears to be driven largely by the *DRD2* gene, rather than by the joint effect of the entire gene-set. After DRD2 exclusion, the enrichment of those gene-sets reduces from 19.1 to 2.0 (Supplementary Table 7). Despite the high fold enrichments of the gene-sets discussed here, it is important to note that they individually contribute to less than 0.5% of schizophrenia’s heritability, further emphasizing its broad polygenic architecture.

For schizophrenia, we additionally compared gene-level fold enrichments starting within a few selected gene-sets (first implicated either as a significant MAGMA association, or through GSA-MiXeR-based ranking with enrichment of at least 15-fold; then constrained to gene-sets with up to 25 genes). Among those genes, only a subset showed strong enrichment (Supplementary Table 8), with no more than half of the genes being responsible for the high fold enrichment. This was confirmed by QQ plots constrained to genetic variants within 10 kilobases (KB) up/down of each gene (Supplementary Figure 4), which showed a “null” QQ plot for a subset of genes, while other genes – particularly *DRD2* and *CACNA1C* – showed strong enrichment. This suggests large heterogeneity in enrichment across genes also within highly enriched gene-sets.

### Sensitivity analysis

We performed additional analyses to test the sensitivity of our results to GSA-MiXeR modeling assumptions. For the enrichment model we compared the three alternatives: (A) the default GSA-MiXeR model, allowing effect size variance to depend on genes; (B) “gene-set” model, allowing effect size variance to depend on gene-sets, but not on individual genes; (C) a “hybrid” model allowing effect size variance to depend on both genes and gene-sets. We expected that model (B) would yield less noisy results, at a price of higher model misspecification as it doesn’t model heterogeneity across individual genes. Another downside is that model (B) must deal with overlap across gene-sets, which we handle through an additive model (see Online Methods). Model (C) was expected to combine the flexibility of model (A) with more robust inference of parameters in model (B). The results are presented in Supplementary Figure 5, with models (A) and (C) yielding similar results, showing that modeling enrichments at the gene level is superior to modeling enrichments at the gene-set level, in line with observations highlighted in Supplementary Figure 4 and Supplementary Table 8. Our choice of model (A) over model (C) for the final analysis was to ensure that enrichment estimate for a particular gene-set does not depend on whether other gene-sets were included in the analysis.

Further comparison of two baseline models (with and without accounting for enrichment in functional categories) showed that these two strategies are generally consistent, with slight inflation in estimates for height if the baseline model does not account for functional categories (Supplementary Figure 5). This suggests that controlling for such enrichment is important, as otherwise some gene-sets might appear enriched simply because of a higher content of enriched functional categories. Additional sensitivity analyses are presented in Supplementary Figure (6), showing only minor effects from out of sample LD reference.

### Replication analysis

In our analysis of real GWAS data we first used MAGMA to filter on significantly associated gene-sets, followed by GSA-MiXeR to re-rank these gene-sets. Here, we present the results of an independent replication analysis using either MAGMA p-values or GSA-MiXeR fold enrichments to rank gene-sets. As a discovery sample, we used half of the UK Biobank panel for the height phenotype, and half of the Psychiatric Genomics Consortium (PGC) schizophrenia sub-studies. The remaining subjects or studies were used as replication data sets. Using independently obtained results for MAGMA and GSA-MiXeR from the discovery and replication samples, we computed the fraction of top-N gene-sets in the discovery samples that also appeared within the top-N gene-sets in the replication samples, allowing N to be 10, 20, 50 or 100 gene-sets. Further, we investigated how the replication rate depended on gene-set size, by computing replication rate for a stratum of gene-sets with up to 25 genes per set, and a second stratum of gene-sets with more than 25 genes per set.

The results are presented in Figure 3 and, in more detail, in Supplementary Figure 7. GSA-MiXeR and MAGMA methods had a comparable replication rate when evaluated on all gene-sets, or in a stratum with more than 25 genes. The biggest differences were observed in the stratum of up to 25 genes, where we consistently observed better replication rates for GSA MiXeR ranking, compared to MAGMA ranking, both for schizophrenia and for height. Overall, the results indicate that replication of GSA-MiXeR based ranking is equivalent to MAGMA based ranking for all gene-sets, and outperforms MAGMA for smaller gene-sets.

**3.**
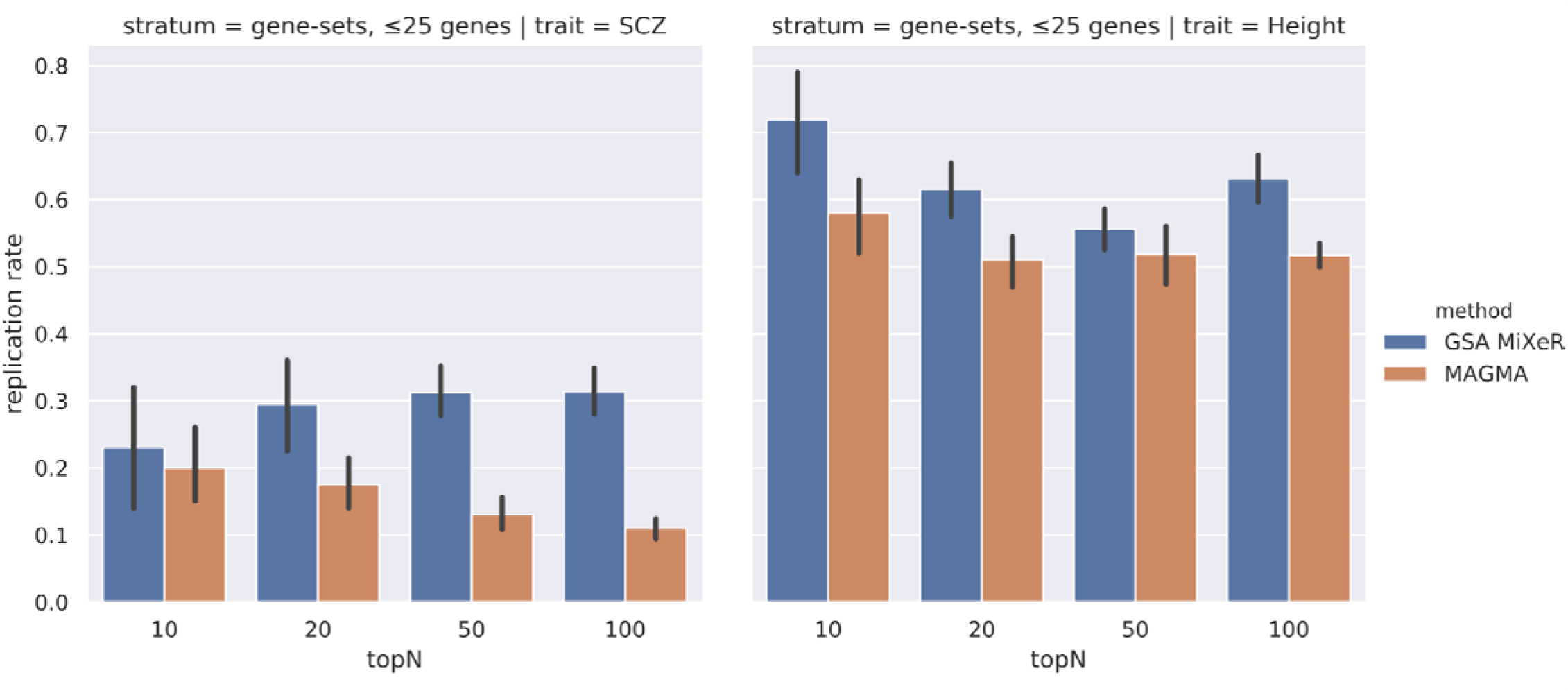
Replication analysis for height (UKB) and schizophrenia (PGC) Replication analysis using schizophrenia (SCZ; PGC sub-studies) and height (UK Biobank) data, with replication rate assessed among gene-sets with up to 25 genes. Replication rate is computed for GSA-MiXeR and MAGMA analyses using two-fold cross-validation, i.e. after applying both methods to two independent and equally sized GWAS sub-samples. After ranking gene-sets according to their fold enrichment of heritability (GSA-MiXeR) and enrichment p-value (MAGMA) obtained in the discovery sample, the replication rate (shown on the vertical axis) was then defined as the fraction of gene-sets that remain within topN gene-sets (N= 10, 20, 50, 100, on the horizontal axis) in the replication half of the dataset.

### Validate prediction accuracy in an independent schizophrenia sample

To further demonstrate the relevance of GSA-MiXeR findings, we tested whether GSA-MiXeR could improve the selection of SNPs for polygenic risk scoring (PGS) over standard p-value based selection. We therefore constructed an enhanced PGS model by re-ranking and clumping SNPs based on posterior effect size estimates from the baseline GSA-MiXeR model. The top-N SNPs were then included in the PGS model, using the original GWAS effect size estimates as weights. We compared our MiXeR-informed PGS to a conventional pruning and thresholding PGS in an independent clinical sample of 743 individuals with schizophrenia and 1074 healthy control subjects, with mean age 32.5 (SD 10.0) years, 52.6% male. This showed that the best-performing model achieved a 13% increase in accuracy, reaching 18.02% Nagelkerke R2 with GSA-MiXeR versus 15.98% for conventional PGS (Figure 4). Most notably, the based-performing GSA-MiXeR-enhanced PGS model included fewer SNPs than the conventional PGS. This indicates that GSA-MiXeR prioritizes SNPs which are more informative for case/control prediction in an independent sample.

**4.**
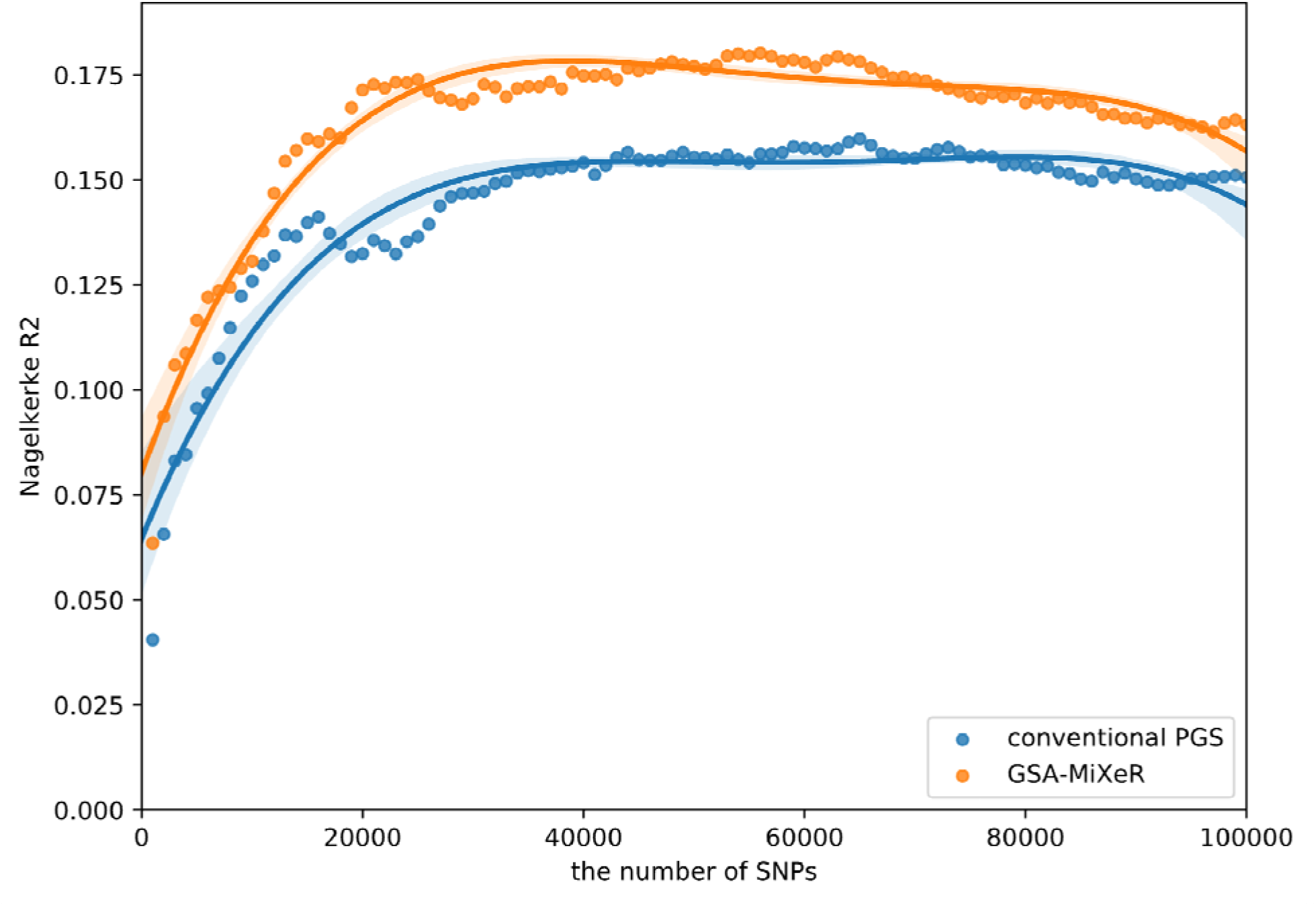
Improved prediction in an independent sample using posterior effect sizes from the baseline GSA-MiXeR model. Comparison between conventional polygenic score (PGS) based on pruning + thresholding strategy and PGS enhanced using posterior effect size estimates from the baseline GSA-MiXeR model, to inform the order in which SNPs were pruned, while keeping the original GWAS effect size estimates as weights. The conventional PGS and GSA-MiXeR enhanced PGS were validated in an independent clinical sample of 743 individuals with schizophrenia and 1074 healthy control subjects, with predictive accuracy measured by Nagelkerke R2 shown on the vertical axis. The horizontal axis indicates the number of SNPs included in each model. The results show that the best-performing model reaching 18.02% with GSA-MiXeR versus 15.98% accuracy of a conventional PGS, at the same time allowing for fewer SNPs in the model.

## Discussion

Here, we present GSA-MiXeR, a novel method for competitive gene-set enrichment analysis which applies stochastic gradient-based optimization for maximum likelihood estimation from GWAS summary statistics. We show that GSA-MiXeR can accurately model the contribution of more than 18,000 genes to SNP-heritability of complex polygenic traits, while controlling for polygenicity, MAF- and LD-dependent genetic architecture, and functional categories. This enables the robust estimation of gene-set fold enrichment alongside partitioned heritability, thus estimating the magnitude of the enrichment, which was previously unfeasible due to computational and methodological limitations. We show through real data from schizophrenia and height how GSA-MiXeR enhances the biological interpretation of GWAS data.

In both simulated and real data, we show GSA-MiXeR’s capability to reorder gene-sets in a way that promotes smaller gene-sets while yielding an equivalent or higher replication rate compared to current standards in the field. When applied to sufficiently powered GWAS study, GSA-MiXeR provides an unbiased estimate of fold enrichments even for small gene-sets with 10 genes or less. GSA-MiXeR’s ability to estimate fold enrichment of a gene-set, rather than its statistical significance (p-value), represents a key advance in GSA, allowing direct comparison of the relative biological effect of implicated gene-sets. This facilitates the promotion of smaller gene-sets with more specific and well-defined functions that can be more readily interrogated in downstream experimental analysis.

When applied to height and schizophrenia, two biologically and genetically distinct complex traits^21^, we demonstrate how fold enrichments can add new, biologically relevant information to MAGMA-based GSA. For height, GSA-MiXeR based ranking promoted highly specific and biologically relevant gene-sets, including gene-sets related to insulin-like growth factor pathways, chondrocyte and osteoblast differentiation and bone mineralization. This contrasted with larger and less specific gene-sets prioritized by MAGMA, such as “negative regulation of signaling”. While a similar pattern was observed for schizophrenia, GSA-MiXeR’s potential for novel mechanistic insights was most clearly illustrated in our exploratory analysis without filtering on significant MAGMA associations. Despite recent advances in schizophrenia GWAS^8,29^, results from GSA analyses have not yet mapped closely onto the primary clinical theories of schizophrenia pathoetiology^23^. It is therefore remarkable that the top two most fold enriched gene-sets were related to dopaminergic neurotransmission, the leading theory of schizophrenia pathogenesis^30-32^. In addition, GSA-MiXeR implicated several other relevant gene-sets including “interneuron migration” and “cerebral cortex GABAergic interneuron development”, both of which may affect parvalbumin-positive interneurons, a class of GABAergic interneurons which have been implicated in schizophrenia pathogenesis [9]. A limitation to these findings is that they are based solely on ranking, which does not constitute statistical inference. It is also worth noting that enrichment in those gene-sets appear to be driven largely by a single gene, *DRD2*. By contrast, the high enrichment of the gene-sets related to voltage-gated calcium channels appear to be a property of all genes in the gene-set, rather than being driven by the most highly enriched gene, *CACNA1C*, alone. Taken together, these findings illustrate that GSA-MiXeR provides the granularity required to map GWAS findings to potentially more informative neurobiological processes which can be tested experimentally, thus facilitating better characterization of the pathobiology of schizophrenia with potential for identifying new druggable targets and clinical sub-groups.

We found that fold enrichments for schizophrenia are markedly lower than for height. This may be due to the clinical heterogeneity and higher polygenicity in schizophrenia, which likely comprise of multiple underlying pathologies^21^. Another factor may be the limited applicability of current gene-set definitions to schizophrenia^5^, as suggested by the large variation in fold enrichment estimates for individual genes within enriched gene sets, which often also included a subset of non-enriched genes (Supplementary Table 8, Supplementary Figure 4).

Previous methods, such as AI-MiXeR^18^, RSS-E^33^ and RSS-NET^34^ have computed enrichment in certain regions of the genome. However, they do not build a comprehensive model including all known human genes, a key advantage of GSA-MiXeR. To achieve this, the GSA-MiXeR model incorporates several parameters describing the distribution of additive genetic effects, fitting those parameters from the GWAS summary statistics. Furthermore, GSA-MiXeR evaluates fold enrichment of a gene-set versus a comprehensive baseline model, which accounts for differential enrichment of functional categories yet without gene-specific effects – an enrichment model which cannot be implemented using a fixed effects approach^35^.

The GSA-MiXeR tool has some limitations. First, it does not output statistical significance of observed fold enrichments. As such, the GSA-MiXeR tool alone cannot be used for statistically valid selection of gene-sets. Indeed, fold enrichment estimates for smaller gene-sets are subject to higher uncertainty and larger sampling variance, which naturally explains why MAGMA and other methods for competitive GSA tend to prioritize larger gene-sets. To justify our exploratory analysis, we confirmed that ranking gene-sets according to GSA-MiXeR fold enrichment is at least as stable (between the discovery and the independent replication dataset) as ranking according to MAGMA p-values. Additionally, we report the standard deviation of parameter estimates across 20 runs of the GSA-MiXeR model, each constrained to a random subset of genetic variants during the fitting procedure. Nevertheless, we advocate that the main usage of the GSA-MiXeR tool is as a secondary analysis of previously identified gene-sets, such as through MAGMA analysis. Second, GSA-MiXeR does not handle continuous-valued genomic annotations as implemented in extended sLDSC^36^. In the future we’re planning to extend GSA-MiXeR analysis to cover genes located on non-autosomes (including sex chromosomes and mitochondrial DNA), and provide reference panel for GSA-MiXeR analysis for non-European populations.

To conclude, GSA-MiXeR estimates fold enrichment and implicates gene-sets with higher biological specificity than current standards. The gene-sets promoted by GSA-MiXeR can provide new insights into the pathobiology of complex diseases, with potential for identification of new drug targets and the development of pathway-specific polygenic risk scores. This may help to advance the classification, diagnosis, and treatment of complex polygenic disorders.

## Data Availability

The datasets analyzed during the current study are available for download from the following URLs: GWAS from Psychiatric Genomics Consortium, https://www.med.unc.edu/pgc/results-and-downloads/downloads; GWAS on UK Biobank, https://github.com/Nealelab/UK_Biobank_GWAS; HRC reference data, https://ega-archive.org/datasets/EGAD00001002729 (upon application); UK Biobank accessed via application 27412, https://bbams.ndph.ox.ac.uk/ams/ (upon application); 1000 Genomes Phase3 data, http://ftp.1000genomes.ebi.ac.uk/vol1/ftp/release/20130502/; MsigDB v7.5, https://www.gsea-msigdb.org/gsea/msigdb/; functional categories, https://alkesgroup.broadinstitute.org/LDSCORE/;

## Ethics statement

Each sample was collected with the participants’ written informed consent and with approval by local institutional review boards. The use of summary statistics for genetic analysis was evaluated by The Norwegian Institutional Review Board: Regional Committees for Medical and Health Research Ethics (REC) South-East Norway and found that no additional ethical approval was required because no individual data were used. Individual level data from the TOP study were analysed under ethical approvals from Norwegian REC (ref. 2009/2485), Data Inspectorate (ref. 03/02051), and The Norwegian Directorate of Health (ref. 05/5821).

## Code availability

MiXeR software and a tutorial example on how to use it will be available online upon publication (https://github.com/precimed/gsa-mixer, v2.0); PLINK software, https://www.cog-genomics.org/plink/2.0/; MAGMA software v1.09b, https://ctg.cncr.nl/software/magma; pipeline to simulate synthetic GWAS data from genotypes: https://github.com/precimed/simu (v0.9.3).

## Acknowledgements

The authors were funded by the Research Council of Norway (#324499, #324252, #273291, #223273, #276082), EU H2020 RealMent (#964874), KG Jebsen Stiftelsen, South East Norway Health Authority (#2022-073). This research has been conducted using the UK Biobank Resource under Application Number 27412. This work used the Extreme Science and Engineering Discovery Environment (XSEDE), including Expanse and OASIS resources provided by San Diego Supercomputer Center at UC San Diego through allocation IBN200001. This work also used the TSD (Tjeneste for Sensitive Data) facilities, owned by the University of Oslo, operated and developed by the TSD service group at the University of Oslo, IT-Department (USIT), using resources provided by UNINETT Sigma2 - the National Infrastructure for High Performance Computing and Data Storage in Norway. C.d.L. was funded by Hoffman-La Roche.

## Author’s contributions

O.F. conceived the study; O.F., A.A.S., and K.O. pre-processed the data; O.F. and D.v.d.M performed all analyses, with conceptual input from G.H., C.d.L., O.A.A. and A.M.D; O.F. and G.H. drafted the manuscript; all authors contributed to and approved the final manuscript.

## Online Methods

### GSA-MiXeR’s full and baseline models

Under the assumptions of a simple additive genetic model, the contribution of the genotype to the phenotype is modelled as a sum of contributions across genetic variants: 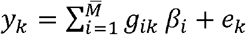, where *y*_*k*_ is a quantitative phenotype or disease liability of *k* -th individual; *g*_*ik*_ is the number of reference alleles for the *i*-th variant (centered to zero mean, but not variance standardized); *β*_*i*_ is per-allele effect size (known also as an additive genetic effect of allele substitution), index *i* runs over 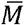 variants. Parameters *β*_*i*_ and *e*_*k*_ are scaled so that phenotype *y* has unit variance.

Following previous work^16-19^, GSA-MiXeR builds on causal mixture model^37^ with a spike-and-slab prior distribution 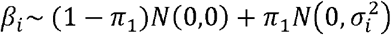, where *N*(0,0) is probability mass at zero (Dirac delta function), *N*(*μ, σ*^2^) is normal distribution, *π*_1_ gives prior probability of a variant to have non-zero effect size, corresponding effect size variance 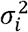 is allowed to vary across genetic variants. To reduce the number of effective parameters being optimized, GSA-MiXeR parametrizes 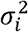 based on gene-sets, functional categories, allele frequency and LD score as follows:

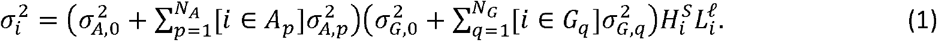

Here, 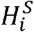 term allows the modeling of architectures dependent on allele frequency, with *H*_*i*_ = 2*f*_*i*_ (1− *f*_*i*_) denoting heterozygosity of *i*-th variant (*f*_*i*_ denoting minor allele frequency of *i*-th variant), and the *S* parameter controlling the effect size distribution along the allele frequency spectrum. Similarly, 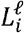 term allows the modeling of LD-dependent genetic architectures, where *L*_*i*_ denotes the LD score of the *i*-th variant and the *ℓ* parameter controls the effect size distribution with respect to LD score. Each of the other two multiplicative factors in equation (1) control differential enrichment across functional categories and across gene-sets, respectively. Index *p* runs across functional categories 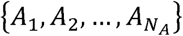 where parameter 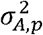 represents the contribution of *p*-th functional category to the variance of the *i*-th genetic variant (if that variant belongs to the category, as indicated by [*i* ∈ *A*_*p*_] term in the formula, where square brackets maps true or false predicate to 1 or 0 – a notation known as Iverson Bracket). If genetic variant *i* belongs to multiple categories, the variances of those categories are added together. Similarly, index *q* runs across gene-sets 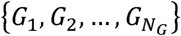 each contributing its own effect size variance 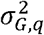 for genetic variants that belong to *G*_*q*_. Each gene-set may, in principle, include just a single gene, allowing GSA-MiXeR to model enrichment of individual genes. Parameters 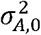 and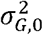 allow for non-zero variance 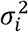 for genetic variants that do not belong to any functional category or gene-set.

For the baseline GSA-MiXeR model we utilized a reduced model (*N*_*G*_ = 2), with gene-sets *G*_1_ and *G*_2_ covering all known protein-coding and pseudo-genes, respectively, in addition to 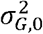 parameter which allows for a different effect size variance in intergenic regions. For the full model we included *N*_*G*_ = 44,247 genes (18,201 protein-coding genes and 26,046 pseudo-genes), but at the same time we choose to disable modeling the annotation categories. In other words, for the baseline model we include annotation categories but exclude gene-sets, while for the full model we exclude annotation categories but include gene-sets. An ideal approach would be to incorporate variable variance across genes into the baseline model, either through constraining already fitted enrichment of annotation categories, or by using previously estimated enrichments as a starting point during joint optimization of annotation and gene-set enrichments. However, due to a purely technical limitation in GSA-MiXeR optimization procedure, the joint optimization of annotation and gene-set enrichment is always performed from scratch. In certain runs this was found to cause convergence issues, thus for the main results we used the model where parameter estimates were not affected by this. Also, while GSA-MiXeR implementation allows fitting all parameters from the data, including the *S* parameter, the main results were obtained with explicitly specified value *S* = −0.25, in line with LDAK recommendation^14^.

### Gene enrichment model

The following expression gives the definition of SNP-based heritability (for a unit-variance phenotype):

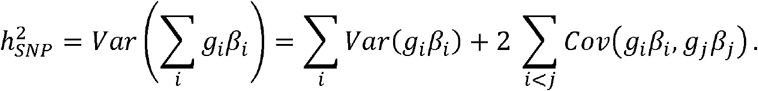

The second term vanishes under the assumptions of the random effects model, leading to heritability being partitioned across variants, and allowing to define heritability of a gene-set G as

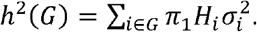

Fold enrichment of heritability can be computed as a ratio of 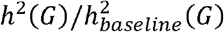, with the numerator estimated using full GSA-MiXeR model, and the denominator estimated using baseline GSA-MiXeR model. However, we observed that fitting one parameter for each gene may lead to slight inflation of heritability estimates, an issue which did not manifest itself when fitting the baseline model. To account for this we compute the fraction of heritability per gene-set, *h*^2^(*G*)/*h*^2^, and define fold enrichment as the ratio between the full model and the baseline model:

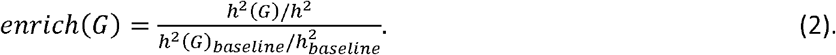

### Log-likelihood computation and its gradients

Let *z*_*j*_ be the GWAS z-score of *j*-th variant out of in total *M* variants tested for association. We assume that association is driven by the underlying additive effects *β*_*i*_, however the set of GWAS variants does not need to be the same as the set of the underlying variants (hence we use different subscripts, *i* and *j*, to distinguish between them). Inference of GSA-MiXeR parameters is based on optimizing log-likelihood function 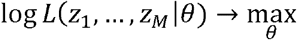 of observing a set of GWAS summary statistics given model parameters, while controlling for the LD structure among variants and their allele frequencies. Log-likelihood computation is based on previously derived methodology^16-18^, which established the following relationship between genetic effects (*β*_*i*_) and GWAS z-scores (*z*_*j*_), highlighting that *z*_*j*_ depends on of *β*_*i*_ all *i*-th variants tagged by *j*-th variant due to LD:

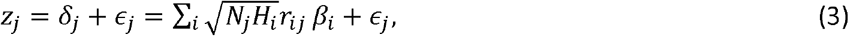

where *N*_*j*_ is sample size at the *j*-th variant, *r*_*ij*_ is the LD allele correlation between *i*-th underlying and *j*-th GWAS variants, and *ϵ*_*j*_ is the normally distributed residual 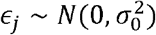, with variance 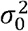 that accounts for potential inflation in GWAS summary statistics. Our two approaches for computing the log likelihood using “fast” and “full” models are described in the Supplementary Note. The “fast” model is based on the method of moments, approximating *p*(*z*_*j*_) is a way that preserves second and fourth moments of the distribution, thus preserving variance and kurtosis; the third moment (skewness) is irrelevant because of symmetry of distribution. The “full” model performs more accurate sampling from the prior distribution *p*(*β*_*i*_) at the expense of longer computation times. Based on these methods of evaluating log-likelihood, we found an efficient computation procedure for computing the gradients 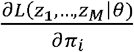 and 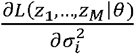 in the context of “fast” and “full” models. Finally, from equation (1) one can compute the Jacobian matrixes 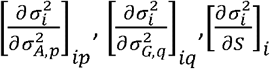 and 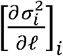, thus allowing the computation of log-likelihood gradients with respect to the parameters of the final model: 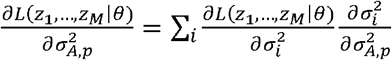, and similarly for all other parameters (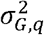, *S, ℓ* and *π*_1_). This can be seen as an equivalent to error back propagation in a neural network, with 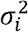 and *π*_*i*_ representing one layer the first layer of the network, and 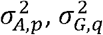 representing the second layer. The total amount of computations needed to calculate the entire gradient vector (for *π*_*i*_ and 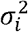 across all variants) is similar to that needed for computing the value of the likelihood function itself. This made it feasible to apply first-order optimization methods while avoiding computational burden despite adding tens of thousands of parameters to the model. For comparison, previous applications of the MiXeR model^16-19^ had a fixed number of 3, 5, 9 and 12 parameters, respectively, and were fitted with zero-order methods including Differential evolution^38^, Nelder–Mead^39^, and Brent’s method^40^.

### Log-likelihood optimization

The log-likelihood function log *L* (*z*_1_, …,*z*_*M*_|*θ*) = ∑_j_ *w*_*j*_ log*p* (*z*_*j*_|*θ*) is weighted across variants using random pruning (64 iterations at LD r2=0.1) as described in ref.^16^. The rationale for weighting SNPs is to avoid over-counting evidence from large LD blocks, in a way that is similar to `--w-ld` option in sLDSC. However, instead of deriving weights based on inverse total LD score, we define weights as probabilities of each variant to remain after random pruning procedure.

Log-likelihood optimization utilizes the Adam algorithm^20^, using parameters *β*_1_ = 0.9, *β*_2_ = 0.99, *ϵ* = 10^−8^. Optimization utilizes N=100 epochs, each of which passes over 22 batches (one batch per chromosome, re-shuffled in a random order for each epoch). The step-size parameter was gradually reduced from *α* = 0.064 down to *α* = 0.0001 by reducing the value by a factor of 2 after each 10 epochs.

Parameters of the GSA-MiXeR model were split into two groups, which are optimized separately. First, the Adam algorithm is applied to optimize 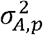 and 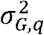 parameters, using constrained values *π*_1_ = 1 (infinitesimal model), *S* = −0.25, *ℓ* = −0.25. As such, variance parameters 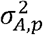 and 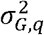 were computed under infinitesimal model (*π*_1_ = 1). Secondly, after constraining 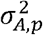 and 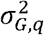 to those found at the previous step, the Adam procedure is used to optimize *π*_1,_*S* and *ℓ* parameters, starting with initial approximations *π*_1_ = 0.001, *S* = −0.25, and *ℓ*= −0.25. Further details of the optimization procedure are available in the Supplementary Note.

### HRC and UKB references (LD structure)

We prepared two reference panels available for GSA-MiXeR analysis using Haplotype Reference Consortia (HRC) and UK Biobank (UKB) datasets.

Our HRC reference has 11,980,511 variants and 23,152 samples after basic QC procedure. During sample QC, we select individuals with EUR ancestry (including Finish population) defined from the first two principal components of 1000 genomes Phase3 data. Further, we prune related individuals using KING software (“`king --unrelated –degree 2`”). For variant QC, we use “`plink --maf 0.001 --hwe 1e-10 --geno 0.1`” parameters, and further exclude markers without RS IDs and excluding duplicated RS IDs.

Our UKB reference has 12,926,669 variants and 337,145 subjects, derived from the UK Biobank imputed v3 dataset. During sample QC, we select unrelated individuals with white British ancestry, remove sex chromosome aneuploidy, and exclude participants who withdrew their consent. Variant QC was applied as follows: “`plink --maf 0.001 --hwe 1e-10 --geno 0.1`”, in addition to filtering variants with imputation INFO score below 0.8 and excluding variants with duplicated RS IDs.

In both references, calculation of allelic correlation (LD r2) was done from hard calls separately for each chromosome with a window size of 10 MB and using a LD r2 threshold of 0.01.

To perform the fitting procedure, we selected variants with MAF above 0.05, then randomly down-sample to a set of 3 million variants and pruned the resulting set of variants based on a LD r2 0.8 threshold. The above procedure was repeated 20 times leading to 20 sets of variants with an average of 655,150 variants per set for UKB reference, and 747,021 variants for HRC reference. The fit procedure for each model was also repeated 20 times (once for each set of variants), and the variability of parameter estimates (such as e.g. gene-set fold enrichment) across these runs was used to derive the standard errors.

### Functional categories and gene-sets

For functional categories, we used the set of 75 binary annotations designed for stratified LD score regression (baselineLD_v2.2).

To define gene boundaries, we used NCBI Entrez database, starting from a set of 61,837 genes which covered both protein-coding (N=19.608 genes) and pseudo genes (N=42,229 genes). After keeping only genes with known coordinates in primary assembly of GRC37 human reference (annotation release 105.20201022), excluding genes on non-autosomes, and excluding genes without variants in MiXeR’s UKB or HRC references we obtained a final list of 44,247 genes (N=18,201 protein-coding genes and N=26,046 pseudo-genes). The boundaries of each gene were extended by 10 KB up- and down-stream, both for the GSA-MiXeR and for the MAGMA analyses.

To define gene-sets, we used GO terms from MsigDB v7.5 (biological processes - c5.bp; cellular component - c5.cc; and molecular function - c5.mf; resulting in 10,402 gene-sets in total). This set was extended using the SynGO database (https://syngoportal.org/), which added 73 gene-sets after excluding those comprising 5 genes or less. Among 10,475 resulting gene-sets there were 19,366 unique genes present, of which 18,245 were present among the list of 44,247 genes available for analysis. To avoid reporting results for highly overlapping gene-sets, we pruned gene-sets which overlap with Dice coefficient of 0.8 or above, retaining smaller gene set from each overlapping pair. This procedure excluded 1760 gene-sets from analysis, and the final list used in analysis contained 8715 gene-sets.

To validate whether gene-set enrichment was largely driven by a single gene, we performed additional leave-one-out analysis for all gene-sets with up to 25 genes, where enrichment was computed after removing one gene at a time, and reporting the gene whose exclusion led to lowest enrichment.

### Summary statistics and individual-level data

For height, we selected the imputed-v3 release for Height (N=360,388) from Neale lab’s analysis (see URLs). For SCZ, we used summary statistics from EUR subset (53,386 cases, 77,258 controls) from PGC wave3 GWAS^8^. In the replication analysis of SCZ, we used sub-study data to perform variance-based meta-analysis on a randomly selected half of the cohorts. To validate prediction, we re-run GSA-MiXeR using summary statistics from PGC wave3 GWAS excluding the Thematically Organized Psychosis (TOP) cohort from Norway with N=743 schizophrenia cases and N=1074 controls, which we use to evaluate accuracy of the polygenic risk scores.

### Simulations

Simulations used a panel of N=337,145 subjects and M=12,926,669 variants from UK Biobank. For each run, we used PLINK to obtain GWAS summary statistics of a quantitative phenotype *y*_*k*_ = ∑_*i*_ *g*_*ik*_ *β*_*i*_ *+ e*_*k*_, where *β*_*i*_ were drawn from a specific model (depending on simulation scenario, as described below), and *e*_*k*_ residual was drawn from a normal distribution with zero mean and variance chosen in a way that sets heritability *h*^2^ = *var* (*G β*) / *var (y)* to a predefined level. A subset of causal variants in simulations were selected by randomly pruning all variants at LD r2=0.1 threshold so that causal variants were in weak LD with each other. This ensured that each genomic region had a well-defined heritability, as for a quantitative phenotype *y = ∑* _*i*_ *g* _*i*_ *β*_*i*_ *+ ϵ* the variance of the genetic component under a fixed effects model is given by 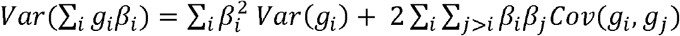, where the second component is negligible if causal variants are not in LD with each other. The remaining component allows the definition of genetic variance in a specific region *g* as 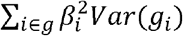.

To generate genetic effects *β*_*i*_, we consider two regions in a genome: *G* (representing the set of all genes) and *g* ⊂ *G*, representing the subset of N=1, 10, or 100 enriched genes. Let *H*_*G*_ and *H*_*g*_ be total heterozygosity for variants in *G* and *g*, respectively. To generate a trait with pre-defined fold enrichments *f*(*g*), we draw *β*_*i*_ from a normal distribution 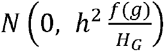 for variants in enriched gene-set *g*, and from 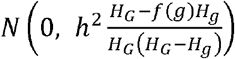 for the remaining variants in *G*\*g*. As shown in the supplementary note, this translates to a true fold enrichment equal to pre-selected *f*(*g*) value. In simulations, we used a basic model for MAF- and LD-dependency (*S* = 0, *ℓ* = 0) also constraining respective GSA-MiXeR parameters to 0, and did not simulate enrichments across functional categories.

We chose to keep the sample size constant throughout all simulations as the power depends on z-scores, which in turn depend on the product of the sample size and heritability. The simulations with N=337,145 and h2=0.1 yield results equivalent to simulations with N=1,000,000 for a trait with SNP heritability of h2=0.03, or to N=100,000 for trait a with h2=0.3. As such, these simulation results are representative of today’s large GWAS studies.

### MAGMA analysis

To ease comparison, MAGMA v1.09b analysis was conducted using the same reference, gene and gene-set definitions as the GSA-MiXeR analysis, except for selecting a random subset of N=1,000 individuals from the reference genotypes. MAGMA commands were as follows:

~~~
magma --annotate window=10 --snp-loc bfile.bim --gene-loc <gene> --out bfile
magma --bfile bfile --pval <sumstats> --gene-annot bfile.genes.annot --out <out>.magma
magma --gene-results <out>.magma.genes.raw --set-annot <geneset> ---out <out>
~~~

## Display Items

### Supplementary Figures

**1.**
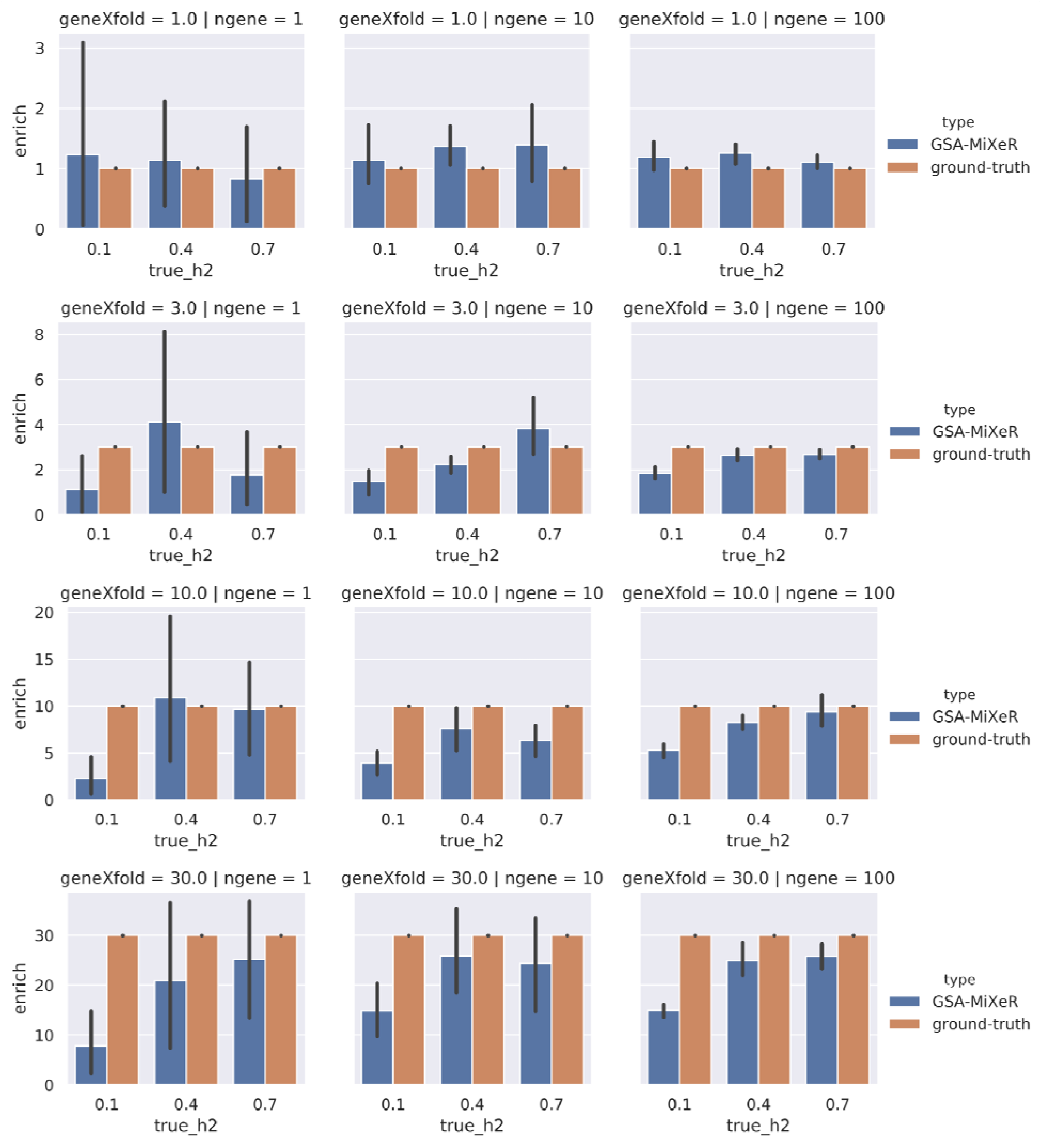
Accuracy of GSA-MiXeR fold enrichment estimates for gene-sets. Accuracy of GSA-MiXeR fold enrichment estimates across multiple simulation scenarios, varying SNP-heritability (h2=0.1, 0.4 and 0.7, on the horizontal axis), and the number genes that were enriched (1, 10, or 100 genes, in columns). The first row of figures corresponds to a scenario without enrichment (geneXfold = 1.0). The remaining three rows correspond to scenarios with 3-, 10- and 30-fold enrichment, respectively. Each simulation was performed 10 times, showing standard deviation among the runs as an error bar.

**2.**
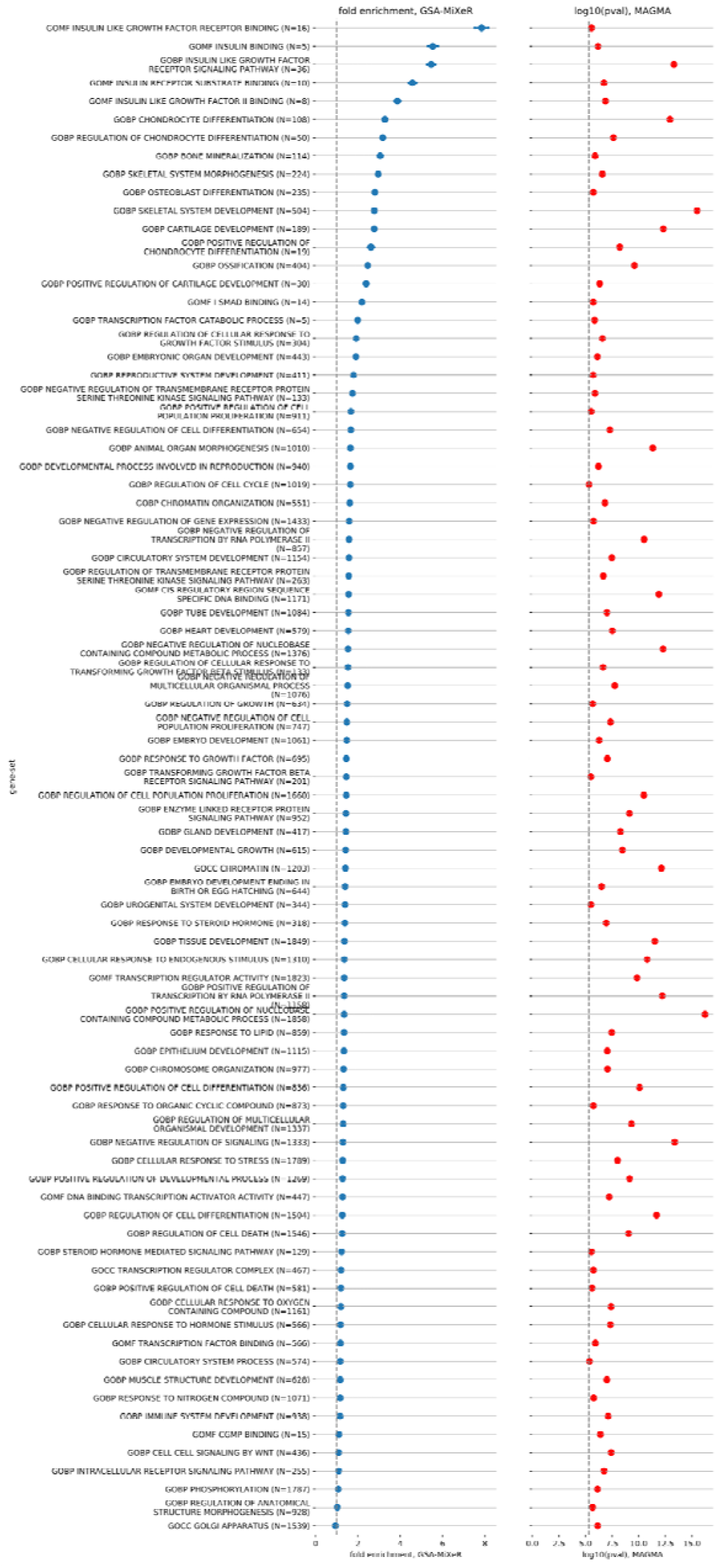
Gene-sets associated with height. Gene-sets associated with height, sorted according to GSA-MiXeR fold enrichment estimate (left-hand panel) or according to MAGMA enrichment p-value (right-hand panel). Appearance of the figure similar to Figure 1.

**3.**
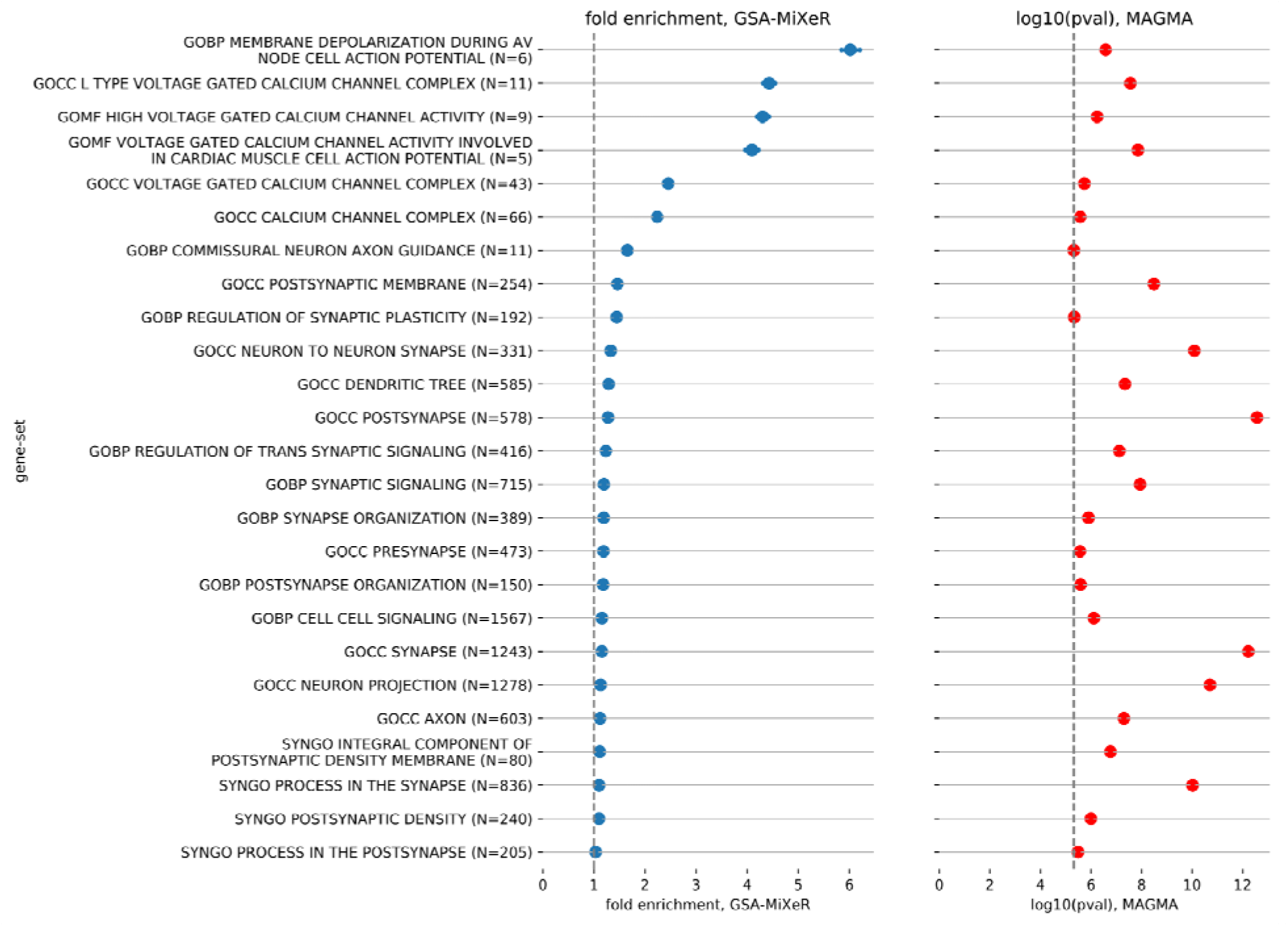
Gene-sets associated with schizophrenia. Gene-sets associated with schizophrenia, sorted according to GSA-MiXeR fold enrichment estimate (left-hand panel) or according to MAGMA enrichment p-value (right-hand panel). Appearance of the figure is similar to Figure 1.

**4.**
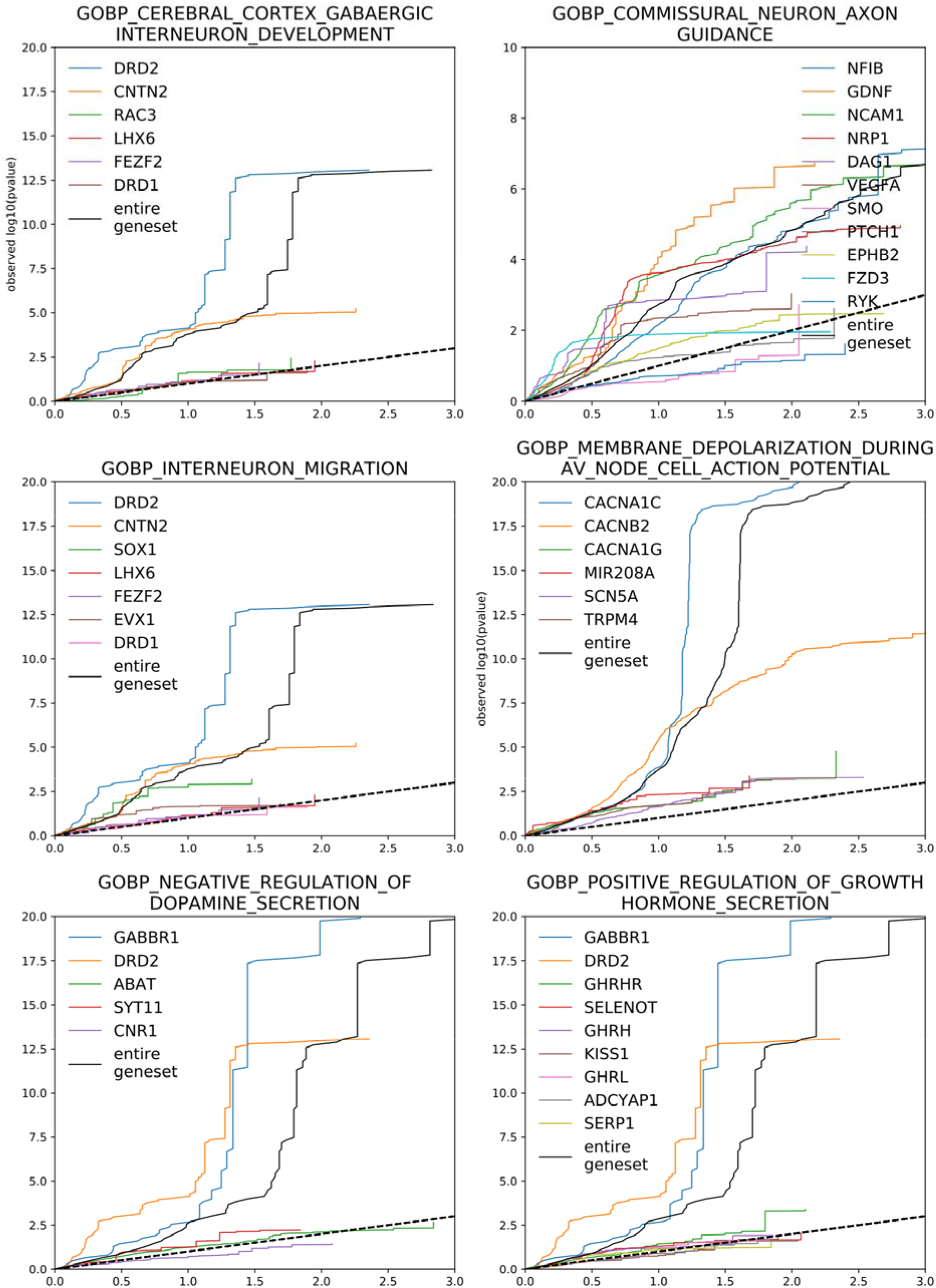
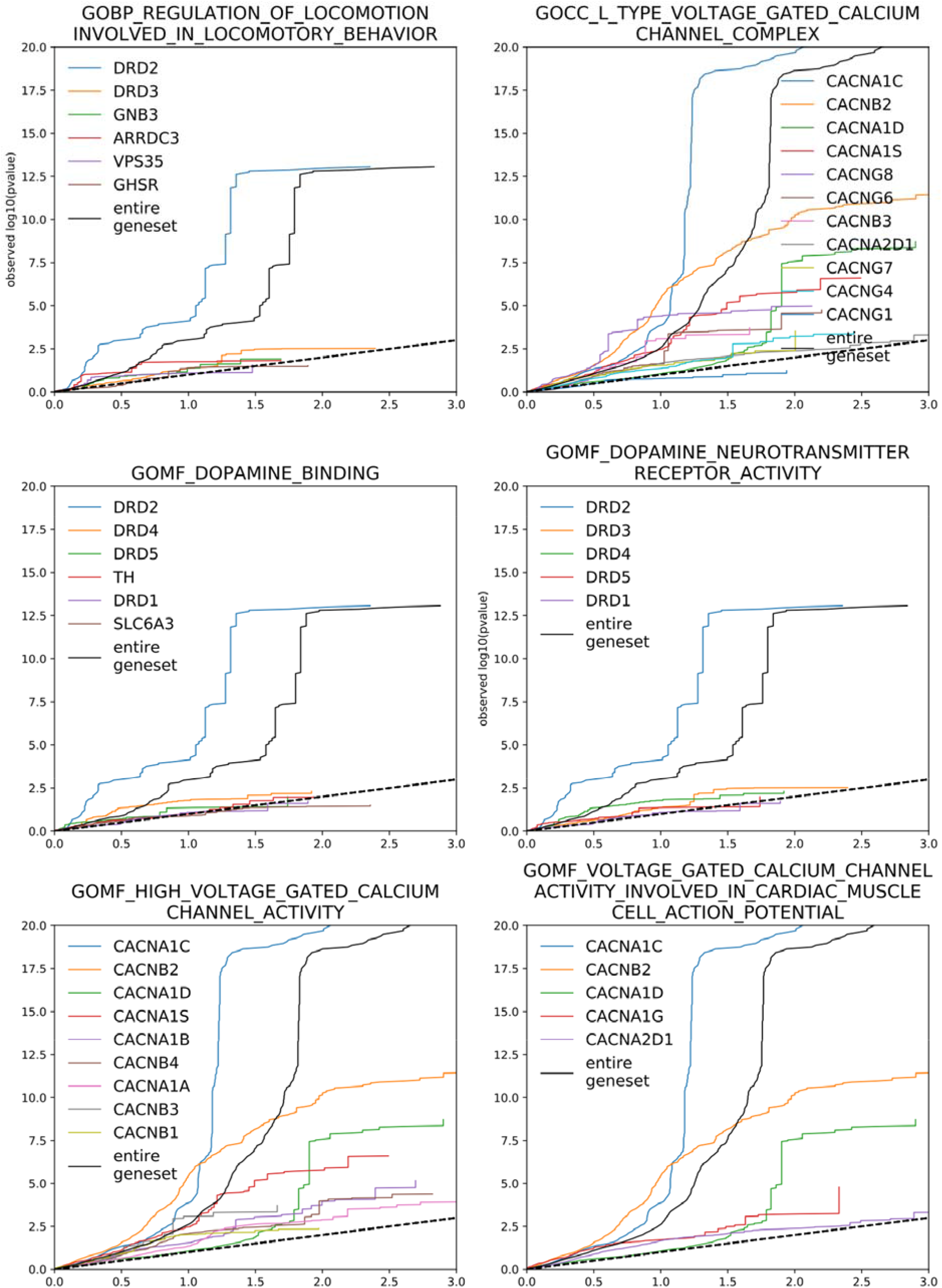
QQ-plots for schizophrenia gene-sets. QQ-plots for selected gene-sets with highest enrichment in schizophrenia (SCZ). Gene-sets were selected either based on MAGMA p-values below 0.05 / N (adjusted for the number of gene-sets), or based on GSA-MiXeR fold enrichment above 15, but only for gene-sets with no more than 25 genes. For each gene-sets, the QQ plots were partitioned by gene, highlighting large heterogeneity of genes within enriched gene-sets.

**5.**
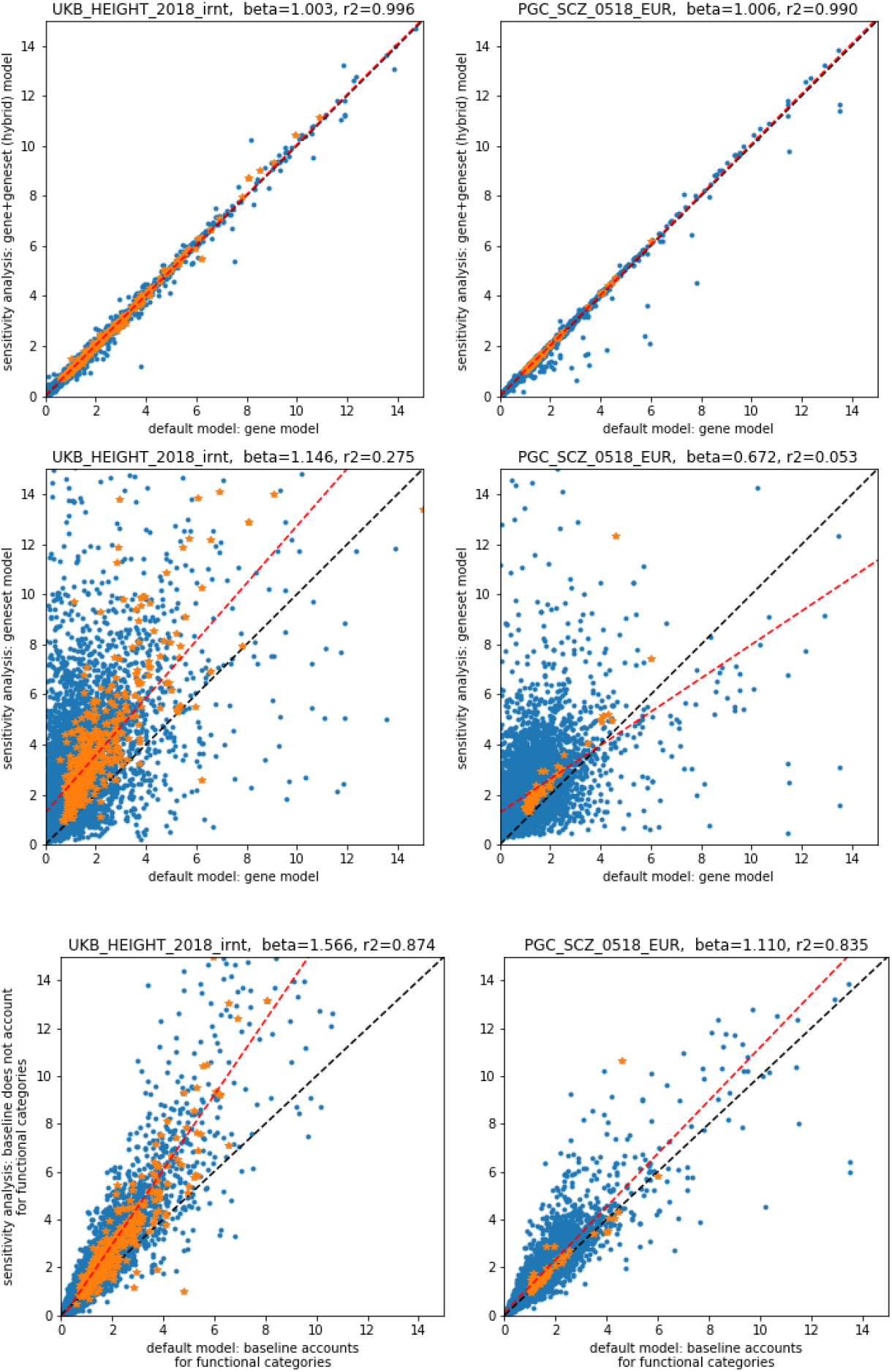
Sensitivity analysis of modeling assumptions for Height and schizophrenia. Sensitivity analysis showing the effect of modeling assumptions on the resulting fold enrichment estimates, using real GWAS summary statistics for height and schizophrenia (SCZ; in columns). Each figure shows a scatter plot of estimated fold enrichments using two different models. On all figures, the horizontal axis corresponds to the model used in the main analysis (each gene was allowed its own effect size variance parameter, and fold enrichment is computed against a “baseline” model which allowed for differential enrichment of functional annotations, but no enrichment of genes). The vertical axis corresponds to several alternative models: “gene-set-level” model (first row), “hybrid” model (second row) or “gene-level” model where baseline didn’t allow for enrichment in functional categories (last row). The “gene-set” model allowed each gene-set to have its own effect size variance. The “hybrid” model combined the “gene-level” and “gene-set-level” models, allowing both genes and gene-sets to have their specific effect size variance parameters. Finally, the last model was equivalent to “gene-level” model, except we did not model differential enrichment across functional categories. Gene-sets marked with a star marker (in orange color) pass MAGMA significance threshold (FDR<0.05), all other gene-sets are shown with point marker (in blue color). Dashed red line shows regression line between two fold enrichment estimates (the default and alternative models), with beta coefficient and R2 shown in the header of each figure.

**6.**
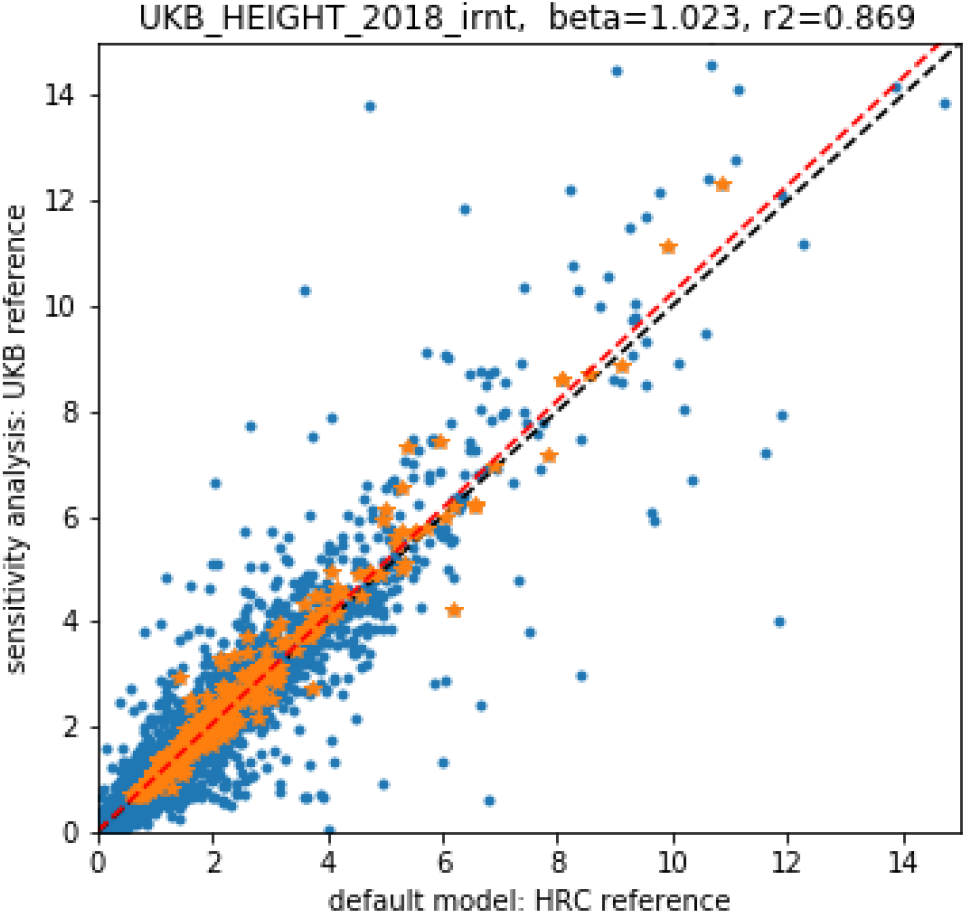
Sensitivity analysis for out-of-sample LD reference. Sensitivity analysis to assess the effects of out-of-sample LD reference on fold enrichment estimates (left panel). The appearance of the figures is similar to the previous figure.

**7.**
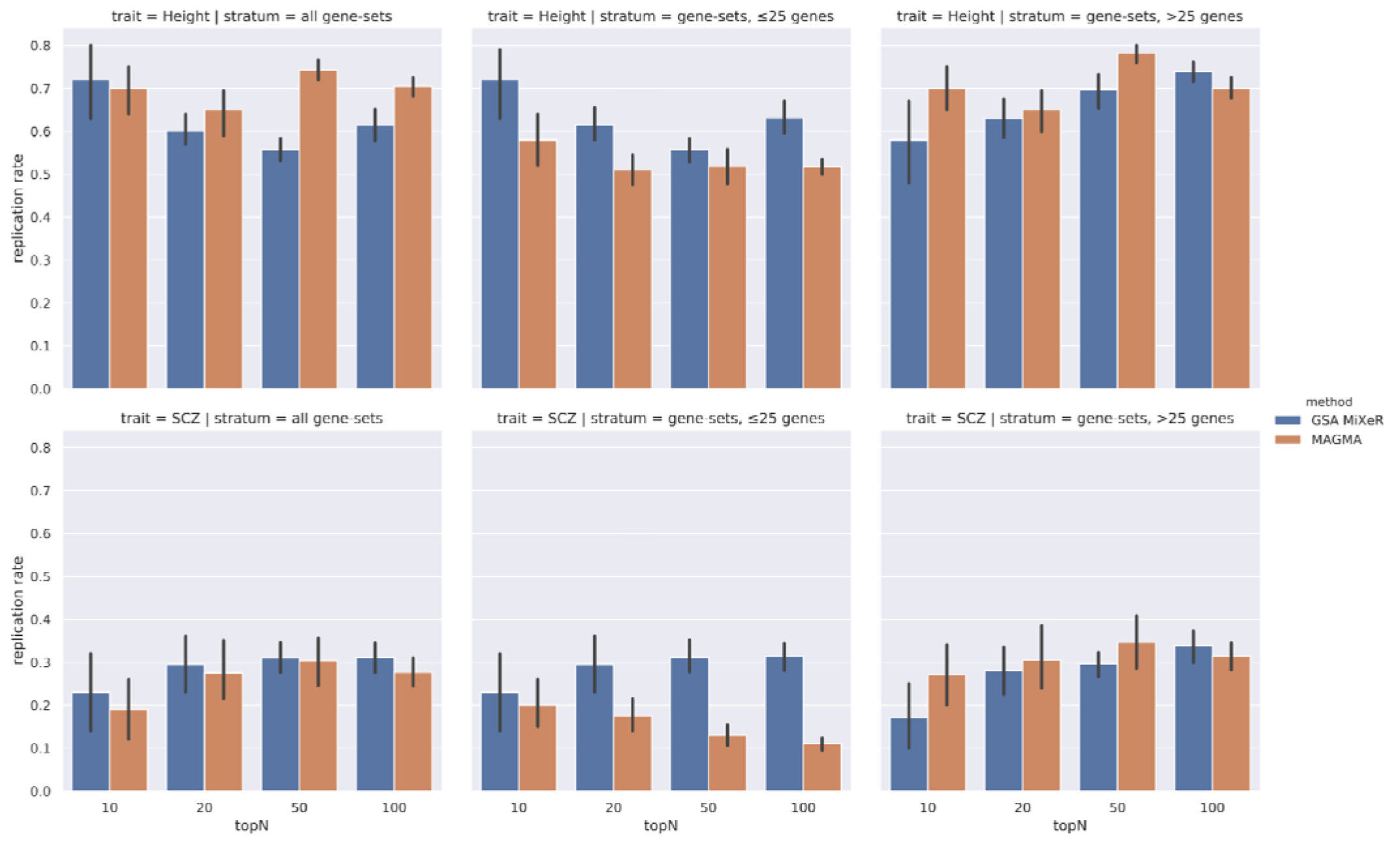
Replication analysis using schizophrenia (PGC sub-studies) and height (UK Biobank) Extended replication analysis showing replication rate for all gene-sets (first column), gene-sets with up to 25 genes (middle column), and gene-sets with more than 25 genes (third column). First row of figures corresponds to schizophrenia (SCZ), second row to height. Figure appearance is as described on Fig. 3.

### List of Supplementary Tables

1. Accuracy of GSA-MiXeR fold enrichment estimates
2. Rank of the enriched gene-set from GSA-MiXeR and from MAGMA analysis
3. Gene-set enrichment for Height, filtered by MAGMA enrichment p-value, and ordered by GSA MiXeR fold enrichment
4. Gene-set enrichment for Height, filtered and ordered by MAGMA enrichment p-value
5. Gene-set enrichment for schizophrenia, filtered by MAGMA enrichment p-value, and ordered by GSA MiXeR fold enrichment
6. Gene-set enrichment for schizophrenia, filtered and ordered by MAGMA enrichment p-value
7. Results from exploratory analysis showing gene-set enrichment for schizophrenia, ordered by GSA MiXeR fold enrichment, without filtering on MAGMA enrichment p-value
8. Enrichment for schizophrenia at the level of individual genes, for a subset of most enriched gene-sets

