## Supplementary Note for "Improved functional mapping with GSA-MiXeR implicates biologically specific gene-sets and estimates enrichment magnitude"

#### SUPPLEMENTARY NOTE 1.

##### The GSA-MiXeR model

Assuming simple additive genetic model, a quantitative phenotype $y \in \mathbb{R}$ is modeled as a linear combination of genotype dosages $g_i$ with weights $\beta_i$:

$$y = \sum_{i=1}^{\bar{M}} \beta_i g_i + e,$$

where index $i$ runs over $\bar{M}$ genetic variants, weights $\beta_i \in R$ are known as *additive effects of allele substitution*, and $e \sim N(0, 1 - h^2)$ term is a normally distributed residual, which reflects contributions from the environment and non-additive genetic effects. The quantitative phenotype $y$ is assumed to be centered and scaled, $E[y] = 0, \mathrm{Var}[y] = 1$, implying that the variance $\mathrm{Var}[\sum_i \beta_i g_i]$ equals to trait's SNP heritability, $h^2 \in [0, 1]$. Genotype dosages $g_i$ are assumed to be centered but not scaled, i.e. $g_i \in \{0 - 2f_i, 1 - 2f_i, 2 - 2f_i\}$, where $f_i$ is allele frequency of the $i$-th variant.

In the GSA-MiXeR model, we postulate a spike-and-slab prior on $\beta_i$:

$$p(\beta_i) \sim (1 - \pi_1)N(\beta_i|0, 0) + \pi_1 N(\beta_i|0, \sigma_i^2),$$

where $\pi_1 \in [0, 1]$ parameter defines weights in a two-component mixture distribution. $\pi_1$ is interpreted as prior probability of a genetic variant to have a non-zero effect. $N(x|\mu, \sigma^2)$ denotes the Gaussian distribution over random variable $x$, except for a special case $N(x|0, 0)$ which indicates probability mass at 0. Parameter $\sigma_i^2$ defines the variance, with each variant having its own $\sigma_i^2$ value. Under these conditions, SNP-heritability of a geneset $G$ is given by $h_G^2 = \pi_1 \sum_{i \in G} H_i \sigma_i^2$, where $H_i = 2f_i(1 - f_i)$ denoting heterozigosity of $i$-th variant.

The GSA-MiXeR model parametrizes $\sigma_i^2$ as follows:

$$\sigma_i^2 = \Big(\sigma_{A,0}^2 + \sum_{p=1\ldots N_a} [i \in A_p]\sigma_{A,p}^2\Big)\Big(\sigma_{G,0}^2 + \sum_{q=1\ldots N_g} [i \in G_q]\sigma_{G,q}^2\Big) H_i^S L_i^\ell. \quad (1)$$

Here $H_i^S$ term allows to model allele frequency dependent architectures, and the $S$ parameter controlling the effect size distribution w.r.t. allele frequency. Similarly, $L_i^\ell$ term allows to model linkage disequilibrium (LD) dependent architectures, where $L_i$ denotes the total LD score of $i$-th variant, and $\ell$ parameter controlling effect size distribution w.r.t. LD score. Each of the other two multiplicative factors in (1) control differential enrichment across genomic annotations and across gene sets. Index $p$ runs across annotation categories $\{A_1, A_2, \ldots, A_{Na}\}$, with parameter $\sigma^2_{A,p}$ representing the contribution of $p$-th annotation category to the variance of $i$-th genetic variant, given that $i$-th variant belongs to $p$-th category, as specified in (1) via indicator variable $[i \in A_p]$, using square brackets notation to transform a logical statement from true or false into 1 and 0, respectively. If genetic variant $i$ belongs to multiple annotation categories, the variances parameters of those annotations with be added together in an additive manner. Similarly, index $q$ runs across gene sets gene sets $\{G_1, G_2, \ldots, G_{Ng}\}$, with parameter $\sigma^2_{G,q}$ representing the contribution of $p$-th gene set. Parameters $\Sigma^2_{A,0}$ and $\Sigma^2_{G,0}$ allow for non-zero variance $\sigma_i^2$ for genetic variants that do not blong to any functional category or gene-set. In order to simplify the notation in the rest of the Supplementary Note, we introduce fake annotation category $A_0$ and gene sets $G_0$ which both consists of all SNPs.

On a technical note, GSA-MiXeR software also supports non-additive interaction of overlapping annotations or gene sets, replacing additive model:

$$\sum_{p=0 \ldots N_a} [i \in A_p]\sigma^2_{A,p} \quad \text{and} \quad \sum_{q=0 \ldots N_g} [i \in G_q]\sigma^2_{G,q}$$

with "smooth-max" model, as follows:

$$(\sum_{p=0 \ldots N_a} [i \in A_a]\sigma^{2p_a}_{A,p})^{1/p_a} \quad \text{and} \quad (\sum_{q=0 \ldots N_G} [i \in G_g]\sigma^{2p_g}_{G,q})^{1/p_g},$$

with parameters $p_a$ and $p_g$ controlling "smooth-max" behavior for overlapping annotations and gene sets, respectively. Note that $p_a$ and $p_g$ parameters of the GSA-MiXeR are unrelated to the $p$ notation used here to index annotation categories in the above formulas. By default GSA-MiXeR implementation will use an additive model across overlapping annotations ($p_a = 1$) and across gene sets ($p_g = 1$). Setting these parameters to e.g. $p_a = 5$ will closely resemble a model where only a single annotation with the largest variance parameter $\sigma_a^2$ (the largest among all annotations containing variant $i$) will contribute to $\sigma_i^2$. Equivalently, setting $p_g = 5$ will trigger similar logic for overlapping gene sets.

#### Log-likelihood computation ("full" z-score model)

To infer parameters of the GSA-MiXeR model we use z-scores ($z_j$) from GWAS summary statistics, where index $j = 1, \ldots, M$ runs over GWAS tag SNPs (imputed or genotyped). Note that the set of GWAS tag SNPs is not necessary the same as the set of genetic variants used in modeling additive genetic effects $\beta_i$, with $i = 1, \ldots, \bar{M}$.. In the GSA-MiXeR implementation, the set of genetic variants is formed by SNPs of the Haplotype Reference Consortia (HRC) reference panel, while a subset of HRC

SNPs that are also present in GWAS summary statistics is used as a set of tag SNPs. Further details about QC procedures applied to HRC and GWAS variants for filtering low quality SNPs are described in the Online Methods section of the manuscript. The following equations related additive effects of allele substitution $\beta_i$ to $z$-scores from GWAS summary statistics.

*Let $\hat{\beta}'_j$ be GWAS estimate of the marginal effect size for $j$-th tag SNP, assessed via univariate linear regression, and $z_j$ be the corresponding z-score, $z_j = \hat{\beta}'_j / \hat{se}(\beta'_j)$. Then*

$$\begin{aligned} & z_j = \delta_j + \epsilon_j, \\ & \delta_j = \sum_{i=1,\ldots,\bar{M}} a_{ij}\beta_i, \ \textit{where}\ a_{ij} = \sqrt{N_j H_i} r_{ij}, \\ & \epsilon_j \sim \mathcal{N}(0, \sigma_0^2), \end{aligned} \tag{2}$$

*where $N_j$ is the number of subjects with non-missing genotype information on $j$-th variant; $H_i = 2f_i(1-f_i)$ is heterozygosity of $i$-th variant; $r_{ij} = corr(\mathbf{v_i}, \mathbf{v_j})$ is an LD allele count correlation between genotype vectors $\mathbf{v_i}$ and $\mathbf{v_j}$ (two column-vectors running across individuals, containing dosages $g_i$ and $g_j$, respectively), and parameter $\sigma_0^2$ accounts for non-polygenic inflation in GWAS z-scores.*

In the absence of covariates, the least square estimate $\hat{\beta}'_j$ can be expressed as

$$\hat{\beta}'_j = \frac{\mathbf{v_j^T y}}{\mathbf{v_j^T v_j}} = \beta_j + \sum_{i \neq j} \hat{\xi}_{ij}\beta_i + \frac{\mathbf{v_j^T e}}{\mathbf{v_j^T v_j}} = \sum_i \sqrt{\frac{H_i}{H_j}} r_{ij}\beta_i + \frac{\epsilon_j}{\sqrt{H_j N_j}},$$

where $\hat{\xi}_{ij} = \mathbf{v_i^T v_j} / \mathbf{v_j^T v_j} = \hat{\zeta}_{ij} / \hat{\zeta}_{jj}$, with $\hat{\zeta}_{ij} = \mathbf{v_i^T v_j}/N$ being an estimate of the covariance between $i$-th and $j$-th variants:

$$\hat{\zeta}_{ij} \simeq \sqrt{2f_i(1-f_i)}\sqrt{2f_j(1-f_j)} r_{ij}.$$

Here the symbol "$\simeq$" denotes asymptotic equality as $n \to \infty$. Then using $\hat{se}(\beta'_j) = 1/\sqrt{H_j N_j}$ result in (2).

Let $\theta = \{\pi_1, \sigma_i^2, \sigma_0^2\}$ be the vector of parameters of the GSA-MiXeR model, where we simplify notation allowing each SNP to have its own $\sigma_i^2$ (but as described above, $\sigma_i^2$ is in turn parametrized as in (1)). We now introduce latent variables $u_i \in \{0, 1\}$ following Bernouli distribution, $p(u_i) = Bern(u_i|\pi_1)$, so that full probabilistic model can be written as follows:

$$\begin{aligned} & p(z_j, \vec{\beta}, \vec{u}|\theta) = p(z_j|\vec{\beta}, \theta) \cdot p(\vec{\beta}|\vec{u}, \theta) \cdot p(\vec{u}|\theta)), \\ & p(z_j|\beta_1, \ldots, \beta_M, \theta) = N\Big(z_j \Big| \sum_{i=1}^{M} a_{ij}\beta_i, \sigma_0^2\Big), \\ & p(\beta_i|u_i = 0, \theta) = N(\beta_i|0, 0), \quad p(\beta_i|u_i = 1, \theta) = N(\beta_i|0, \sigma_i^2), \\ & p(u_i|\theta) = Bern(u_i|\pi_1). \end{aligned}$$

A tricky part here is that $z_j$ may depend on multiple $\beta_i$. After observing $\vec{z} = (z_1, \ldots, z_M)^T$, we are aiming to do inference on $\theta$ by max. likelihood:

$$p(\vec{z}|\theta) = \prod_j \int_u \int_\beta p(z_j, \vec{\beta}, \vec{u}, \theta) du d\beta \to \max_\theta$$

In GSA-MiXeR, we approximate the above integral by drawing $K = 20000$ samples from $p(u_i)$ for each latent variable $u_i$. For $j$-th variant and $k$-th sampling iteration, let $U_{jk}$ be the set of variants with $u_i = 1$. For each realization of latent variables the distribution over $p(z_j|U_{jk}, \theta)$ became normal zero-mean distribution with a simple analytical formula for its variance:

$$p(z_j|U_{jk}, \theta) = N(z_j|0, \Sigma^2_{jk}),$$
$$\Sigma^2_{jk} = \sigma^2_0 + \sum_{i \in U_{jk}} a_{ij}\sigma^2_i.$$

On a technical note, we would like to point out that in practice it is impossible to store all elements of the $a_{ij}$ matrix ($a_{ij} = \sqrt{N_j H_i} r_{ij}$) due to its size. Nevertheless, most of its elements are close to zero, and in GSA-MiXeR we use sparse matrix formats to store all $a_{ij}$ with $r^2_{ij} \geq r^2_{min}$, by default $r^2_{min} = 0.01$, considering all pairs of variants within each chromosome regardless of the distance between them, but neglecting potential correlations between different chromosomes. However, the truncated parts of the LD structure (with $r^2_{min} < 0.01$) may result in a slightly inflated $\sigma^2_0$ parameter estimates. To reduce this effect, GSA-MiXeR computes partial LD scores $\tilde{L}_j = \sum_{i:\, r^2_{ij} < r^2_{min}} a^2_{ij}$, and allow them to contribute to $\Sigma^2_{jk}$ under the assumptions of an infinitesimal model with a new variance parameter $\sigma^2_{0L}$ to be estimated from the data:

$$\Sigma^2_{jk} = \sigma^2_0 + \tilde{L}_j \sigma^2_{0L} + \sum_{i \in U_{jk}} a_{ij}\sigma^2_i.$$

To clarify how this works, let's consider an infinitesimal model with $\pi_1 = 1$ (all additive genetic effects have non-zero variance) and $\sigma^2_i = \sigma^2_\beta$ (equal variance across all genetic variants). Under these assumptions we can use the above formulas with $u_i = 1$ across all variants to derive $p(z_j|\theta) = N(z_j|0, \sigma^2_0 + \sum_i a^2_{ij}\sigma^2_\beta)$. In this formula $L_j = \sum_i a^2_{ij} = \sum_i N_j H_i r^2_{ij}$ plays a role of LD score, with linear relationship between $E(z^2)$ and $L_j$ under the assumptions of an infinitesimal model. In GSA-MiXeR, partial LD scores $\tilde{L}_j$ are, by default, computed across variants with $0.0001 \leq r^2_{ij} < 0.01$, and user has possibility to constraint $\sigma^2_{0L}$ to 0, or let GSA MiXeR fit $\sigma^2_{0L}$ parameter from the data. By default, $\sigma^2_{0L}$ is set zero 0, and this setting was used throughly all analyses in this work.

Final expression for log-likelihood in GSA-MiXeR is as follows:

$$\log L = \sum_j w_j \log p_j = \sum_j w_j \log \Big( \frac{1}{K} \sum_k q(z_j; 0, \sigma^2_0 + \tilde{L}_j \sigma^2_{0L} + \sum_{i \in U_{jk}} a_{ij}\sigma^2_i) \Big) \quad (3)$$

where weights $w_j$ are induced by random-pruning technique to avoid over-counting contribution from large LD blocks, and $q(z; 0, s^2) = \phi(z; 0, s^2) = \frac{1}{\sqrt{2\pi}s} e^{-z^2/2s^2}$ stands for the probability density function of centered normal distribution, except for z-scores that are very large. For large z-scores that exceed certain threshold $|z_j| \geq z_{max}$, the log-likelihood is computed based on right-censoring, i.e. with $q(z_j; 0, s^2) = 2\Phi(-z_{max}; 0, s^2) = erfc\left(\frac{z_{max}}{\sqrt{2}s}\right)$. A typical value for $z_{max}$ is 5.45, which corresponds to conventional genome-wide significance threshold $\alpha = 5 \times 10^{-8}$.

#### Log-likelihood gradients

To maximize log-likelihood w.r.t. parameters $\theta$, GSA-MiXeR employ Adam algorithm, a stochastic gradient-based optimization, which require a gradient of the log likelihood w.r.t. all parameters. By inspecting (3) we observe that it is trivial to compute gradients w.r.t. $\sigma_0^2$, $\sigma_{0L}^2$, and $\sigma_i^2$:

$$\begin{aligned}
\frac{\partial \log L}{\partial \sigma_i^2} &= \sum_j \frac{w_j}{Kp_j} \sum_{k:\, i \in U_{jk}} q'_{jk} a_{ij}^2, \\
\frac{\partial \log L}{\partial \sigma_0^2} &= \sum_j \frac{w_j}{Kp_j} \sum_k q'_{jk}, \\
\frac{\partial \log L}{\partial \sigma_{0L}^2} &= \sum_j \frac{w_j \tilde{L}_j}{Kp_j} \sum_k q'_{jk},
\end{aligned} \tag{4}$$

where

$$\begin{aligned}
q_{jk} &= q(z_j; 0, s^2), \quad \text{with} \quad s^2 = \sigma_0^2 + \tilde{L}_j \sigma_{0L}^2 + \sum_{i \in U_{jk}} a_{ij} \sigma_i^2, \\
q'_{jk} &= \frac{\partial \phi(z_j; 0, s^2)}{\partial s^2} = \frac{z^2 - s^2}{2s^4} q_{jk} \;\text{ when }\; |z_j| < z_{max}, \;\text{ otherwise} \\
q'_{jk} &= \frac{\partial 2\Phi(-z_{max}; 0, s^2)}{\partial s^2} = \frac{z_{max} e^{\frac{z_{max}^2}{2s^2}}}{\sqrt{2\pi} s^3} = \frac{z_{max}}{s^2} \phi(z_{max}; 0, s^2).
\end{aligned}$$

Generally, $s^2$ in the above formulas should appear as $s_{jk}^2$, i.e. indexed by $j$ and $k$ indices; those indices were omitted in the formulas above in favor of less heavy notation. When implemented in GSA-MiXeR software, these formulas result in a numeric vector with length $M + 2$, with two last elements corresponding to $\sigma_0^2$ and $\sigma_{0L}^2$ gradient, and the remaining elements corresponding to $M$ gradients one for each genetic variant, with $M$ typically being around $10^7$. Interestingly, computation of all these derivatives jointly take approximately the same time as computation of the log likelihood function, because for each $\sigma_i^2$ the gradient only depends on a smaller set of GWAS tag SNPs that are in LD with $i$. Another good illustration for this is error back propagation in neural networks, which also take a similar time to computing forward pass over the neural network.

To conclude the computation of log-likelihood gradients we need to consider all parameters of the GSA-MiXeR model that needs to be optimized. These parameters are as follows:

- $\pi_1$ - the polygenicity parameter;
- $\sigma_i^2$ - the variance parameter of each SNP, which in turn depend on $S$ and $\ell$ (parameters of effect size distribution w.r.t. allele frequency and LD), $\sigma^2_{A,p}$ (a set of variance parameters, one for each annotation category) and $\sigma^2_{G,q}$ (a set of variance parameters, one for each gene set);
- $\sigma_0^2$ - inflation parameter;
- $\sigma^2_{0L}$ - inflation capturing the truncated parts of the LD matrix.

To avoid having boundaries on these parameters during optimization, we further parametrize $\pi_1$ using logistic function, $\pi_1(x) = \frac{e^x}{1+e^x}$, with $x$ here being the actual parameter used during optimization; all non-negative variance parameters are further parametrized with an exponential function, $\sigma^2(A,p) = e^{x_p}$ and $\sigma^2(G,q) = e^{y_q}$; parameters $S$ and $\ell$ are unbounded, and thus are used directly during optimization.

Now, we further re-parametrize $\sigma_i^2$ as follows, to reduce collinearity among parameters:

$$\sigma_i^2 = \frac{\sigma_\beta^2}{\pi_1}\Big(\sum_{p=0\ldots N_a}[i \in A_p]e^{p_a x_p}\Big)^{\frac{1}{pa}}\Big(\sum_{q=0\ldots N_g}[i \in G_q]e^{p_g y_q}\Big)^{\frac{1}{pg}}\frac{H_i^S}{\frac{1}{\bar{M}}\sum_t H_t^S}\frac{L_i^\ell}{\frac{1}{\bar{M}}\sum_t L_t^\ell}, \quad (5)$$

where $t = 1, \ldots, \bar{M}$ runs across all SNPs. The above formula differs from equation (1) only by a factor $\frac{\sigma_\beta^2}{\pi_1}\frac{1}{\frac{1}{\bar{M}}\sum_t H_t^S}\frac{1}{\frac{1}{\bar{M}}\sum_t L_t^\ell}$ that is the same for all SNPs, thus it does not change relative effect size distribution across genetic variants. Here we introduced a global $\sigma_\beta^2$ parameter which allows to simultaneously adjust all $\sigma_i^2$ up or down, which plays a role if one fits the GSA-MiXeR model with constrained variance parameters for annotation categories and gene sets. Further, we divided all $\sigma_i^2$ by polygenicty parameter $\pi_1$, making sure that polygenicity parameter itself does not affect heritability estimate $h^2 = \sum_i \pi_i H_i \sigma_i^2$. Finally, we normalize the formula by mean $H_i^S$ over genetic variants, and also by mean $L_i^\ell$ over genetic variants, so that changes in $S$ and $\ell$ parameters also have a minimal effect on the heritability estimate. The idea behind parametrization (5) is that heritability estimate $h^2 = \sum_i \pi_i H_i \sigma_i^2$ has a substantial impact on log-likelihood, and fine-tuning of other parameters that control intricate aspects of the genetic architecture is more robust when total heritability is constrained to a given value.

Now everything is set to finalize gradient computation.

The $\pi_1$ parameter is treated specially, in a sense that $\frac{\partial \log L}{\partial \pi_1}$ is estimated numerically using finite difference

$$\frac{\log L(x+\Delta x) - \log L(x - \Delta x)}{2x},$$

where step $\Delta x$ changes adaptively depending on the iteration, and is set to the value used in the Adam algorithm.

We already derived gradients for $\sigma_0^2$ and $\sigma_{0L}^2$, except for the parametrization $\sigma_0^2(x)$ which uses an exponential function, $\sigma_0^2(x) = e^x$, so we need to use corresponding gradient w.r.t $x$, i.e. $\frac{\partial \log L}{\partial x} = \frac{\partial \log L}{\partial \sigma_0^2} \times \frac{\partial \sigma_0^2(x)}{\partial x} = \sigma_0^2 \frac{\partial \log L}{\partial \sigma_0^2}$, and similarly for $\sigma_{0L}^2$.

All of the remaining parameters $(S, \ell, \sigma^2_\beta(x), \sigma^2_{A,p}(x_p), \sigma^2_{G,q}(y_q))$ enter into log likelihood only via $\sigma^2_i$, thus we will now derive $\frac{\partial\sigma^2_i}{\partial S}$, $\frac{\partial\sigma^2_i}{\partial \ell}$, $\frac{\partial\sigma^2_i}{\partial x}$, $\frac{\partial\sigma^2_i}{\partial x_p}$, $\frac{\partial\sigma^2_i}{\partial y_q}$, and then calculate final gradient as follows:

$$\frac{\partial \log L}{\partial S} = \sum_i \frac{\partial \log L}{\partial \sigma^2_i}\frac{\partial \sigma^2_i}{\partial S},$$

and similarly for all other parameters.

Starting with $\frac{\partial \log L}{\partial S}$ and $\frac{\partial \log L}{\partial \ell}$, let us introduce shorthand notations $T(S) = \sum_t H^S_t/\bar{M}$ and $T(\ell) = \sum_t L^\ell_t/\bar{M}$. Then

$$\Big(\frac{H^S_i}{T(S)}\Big)' = \frac{\frac{\partial H^S_i}{\partial S}T(S) - H^S_i T'(S)}{T^2(S)} = \frac{H^S_i}{T(S)}\frac{(\log H_i)T(S) - \big(\sum_t (\log H_t)H^S_t/M\big)}{T(S)}.$$

Hence

$$\frac{\partial\sigma^2_i}{S} = \sigma^2_i \frac{T(S)\log H_i - \big(\sum_t (\log H_t)H^S_t/\bar{M}\big)}{T(S)},$$

and similarly

$$\frac{\partial\sigma^2_i}{\ell} = \sigma^2_i \frac{T(\ell)\log L_i - \big(\sum_t (\log L_t)L^\ell_t/\bar{M}\big)}{T(L)}.$$

Note that both $T(S)$, $T(\ell)$ as well as $\sum_t (\log H_t)H^S_t$ and $\sum_t (\log L_t)L^\ell_t$ are independent of $i$, thus computation of these derivatives takes linear time w.r.t. the number of variants $M$.

Turning to $\sigma^2_{A,p}$ and $\sigma^2_{G,q}$, which are in turn parameterized by exponential functions $\sigma^2_{A,p}(x_p) = e^{x_p}$ and $\sigma^2_{G,q}(y_q) = e^{y_q}$, let us first consider a "smooth-max" function $F(x_1, \ldots, x_N) = \Big(\sum_{u=1}^N e^{px_u}\Big)^{\frac{1}{p}}$ and explore the derivative $\frac{\partial F}{\partial x_t}$:

$$\frac{\partial F(x_1,\ldots,x_N)}{x_t} = \frac{1}{p}\big(\sum_u e^{px_u}\big)^{\frac{1}{p}-1} p e^{px_t} = F(x_1,\ldots,x_N)^{1-p} e^{px_t} = F(x_1,\ldots,x_N)\frac{e^{px_t}}{\sum_u e^{px_u}}.$$

As such,

$$\frac{\partial\sigma^2_i}{\partial x_p} = \sigma^2_i \frac{[i \in A_p]e^{p_a x_p}}{\sum_u [i\in A_u]e^{p_a x_u}} = \sigma^2_i \frac{[i \in A_p]\sigma^{2p}_{A,p}}{\sum_u [i\in A_u]\sigma^{2p_a}_{A,u}}.$$

In a matrix form, $\frac{\partial\sigma^2_i}{x_p}$ follows the same sparsity structure as $[i \in A_p]$ matrix with $\bar{M}$ rows (one row per SNP) and $N_a$ columns (one column per annotation), with each row of this matrix being multiplied by $\sigma^2_i / \sum_u [i \in A_u]\sigma^{2p_a}_{A,u}$ value (note this value only depends on $i$, but not on annotation $p$), and each column being multiplied by $\sigma^{2p_a}_{A,p}$ (a value that only depends on annotation $p$, but not on $i$). Such rows-wise and column-wise multiplications can be implemented using matrix-matrix produce with a diagonal matrix. Also, note that "smooth-max" function, being also a $||x||_p$ p-norm, scales linearly w.r.t. scale of its argument, so we are free to re-scale $\sigma^2_u$ by dividing it to its mean to ensure numeric stability even with single precision float-point numbers.

Similarly to $\frac{\partial\sigma^2_i}{\partial x_p}$, for the gene sets we can derive that

$$\frac{\partial\sigma^2_i}{\partial y_q} = \sigma^2_i \frac{[i \in G_q]\sigma^{2p}_{G,q}}{\sum_u [i\in G_u]\sigma^{2p_g}_{G,u}}.$$

And finally, for the $\sigma^2_\beta$ that is also parametrized by an exponential function $\sigma^2_\beta(x) = e^x$, we have $\frac{\partial \sigma^2_i}{\partial x} = \sigma^2_i$. This concludes all pieces needed for efficient gradient computation of the log likelihood function w.r.t. the parameters of the GSA-MiXeR model.

##### Gaussian approximation ("fast" z-score model)

In this section we will assume $\beta_i \sim (1-\pi_i)N(0,0) + \pi_i N(0, \sigma^2_i)$, allowing for $\pi_i$ to vary across SNPs, in addition to varying $\sigma^2_i$ across SNPs as modeled previosuly.

**Lemma**. Raw moments of $E\beta^2_i$ and $E\beta^4_i$ are as follows:

$$\begin{aligned} E\beta^2_i &= \pi_i \sigma^2_i, \\ E\beta^4_i &= 3\pi_i \sigma^4_i \end{aligned} \tag{6}$$

Let $X_i$ be a set of independent random variables, and $Y = \sum_i \pi_i X_i$ be mixture distribution. Then all raw moments $E[Y^n]$ are given by a linear combination $\sum_i \pi_i E[X^n_i]$. This trivial fact follows directly from the definition of the expectation operator: $E[Y^n] = \int_{-\infty}^{\infty} x^n dF_Y(x) = \int_{-\infty}^{\infty} x^n \sum_i \pi_i dF_{X_i}(x) = \sum_i \pi_i E[X^n_i]$. ■

**Lemma**. Let $\delta_j = \sum_i a_{ij}\beta_j$, where $a_{ij} = \sqrt{N_j}\sqrt{H_i} r_{ij}$. Then raw moments of $E\delta^2_j$ and $E\delta^4_j$ are as follows:

$$\begin{aligned} E\delta^2_j &= \sum_i a^2_{ij} E\beta^2_i = \sum_i a^2_{ij} \pi_i \sigma^2_i \\ E\delta^4_j - 3(E\delta^2_j)^2 &= \sum_i a^4_{ij} \Big(E\beta^4_i - 3(E\beta^2_i)^2\Big) = \sum_i 3a^4_{ij}\pi_i(1-\pi_i)\sigma^4_i \end{aligned} \tag{7}$$

First equation $E\delta^2_j = \sum_i a^2_{ij} E\beta^2_i$ is simply because $\beta_i$ are independent zero-mean random variables.

Let $A_i$ and $B_i$ be some arbitrary expressions. Then $(\sum_i B_i)^2 = \sum_i B^2_i + 2\sum_{i<j} B_i B_j$, and $(\sum_i A_i)^4 = \sum_i A^4_i + 6\sum_{i<j} A^2_i A^2_j + \cdots$, where omitted terms contain at least one odd power of $A_i$. Consequently, if $A_i$ are independent zero-mean random variables, then $E[(\sum_i A_i)^4] = E[\sum_i A^4_i + 6\sum_{i<j} A^2_i A^2_j] = \sum_i E[A^4_i] + 6\sum_{i<j} E[A^2_i]E[A^2_j]$. Using the above expression for $(\sum_i B_i)^2$ with $B_i = E[A^2_i]$, we get $2\sum_{i<j} E[A^2_i]E[A^2_j] = (\sum_i E[A^2_i])^2 - \sum_i (E[A^2_i])^2$. Therefore,

$$E[(\sum_i A_i)^4] = \sum_i E[A^4_i] + 3\Big((\sum_i E[A^2_i])^2 - \sum_i (E[A^2_i])^2\Big).$$

Replacing $(\sum_i E[A^2_i])$ with $E[(\sum A_i)^2]$, we get

$$E[(\sum_i A_i)^4] - \Big(E[(\sum A_i)^2]\Big)^2 = \sum_i \Big(E[A^4_i] - 3(E[A^2_i])^2\Big).$$

Using expression $A_i = a_{ij}\beta_i$, we get

$$E\delta^4_j - 3(E\delta^2_j)^2 = \sum_i a^4_i \Big(E\beta^4_i - 3(E\beta^2_i)^2\Big). \quad ■$$

**Lemma**. Let $A$ and $B$ be two numeric values, $\tilde{\delta}_j = p_0 N(0,0) + p_1 N(0,s^2)$ so that

$$p_0 = \frac{B}{B+3A^2}, \quad p_1 = \frac{3A^2}{B+3A^2}, \quad s^2 = \frac{B+3A^2}{3A}. \tag{8}$$

Then, by trivial computation, $E\tilde{\delta}_j^2 = A$, and $E\tilde{\delta}_j^4 - 3(E\tilde{\delta}_j^2)^2 = B$. ■

Substituting $A_j \equiv E\delta_j^2 = \sum_i a_{ij}^2 \pi_i \sigma_i^2$ and $B_j \equiv E\delta_j^4 - 3(E\delta_j^2)^2 = \sum_i 3a_{ij}^4 \pi_i (1-\pi_i)\sigma_i^4$ into (8) yields explicit formulas defining $p_{j0}$, $p_{j1}$ and $s_j^2$ for the distribution $\tilde{\delta}_j = p_{0j} N(0,0) + p_{1j} N(0, s_j^2)$ which approximates true distribution for $\delta_j = \sum_i a_{ij}\beta_i$, in a sense that $E\delta_j^2 = E\tilde{\delta}_j^2$ and $E\delta_j^4 = E\tilde{\delta}_j^4$. From these expressions one can derive explicit formulas for computation of the gradients $\frac{\partial p(z_j)}{\partial \pi_i}$ and $\frac{\partial p(z_j)}{\partial \sigma_i^2}$.

#### Log-likelihood optimization

Log-likelihood optimization in GSA-MiXeR utilize Adam algorithm (1), using parameters $\beta_1 = 0.9$, $\beta_2 = 0.99$, $\epsilon = 10^{-8}$. In total optimization utilize $N = 50$ epochs, each of which passes over 22 batches, one per chromosome, in a random order. The stepsize parameter is gradually reduced from $\alpha = 0.064$ down to $\alpha = 0.0001$, reducing its value by a factor of 2 after each 5 epochs.

Parameters of the GSA-MiXeR model are split into two groups which are optimized separately. First, Adam algorithm is applied to optimize $\sigma^2_{A,p}$ and $\sigma^2_{G,q}$ parameters, using constrained values $\pi_1 = 1$, $S = -0.25$, $\ell = -0.25$. Secondly, constraining $\sigma^2_{A,p}$ and $\sigma^2_{G,q}$ parameters to those found at the previous step, Adam procedure is used to optimize $\pi$, $S$ and $\ell$ parameters, starting with initial approximations $\pi_1 = 0.001$, $S = -0.25$ and $\ell = -0.25$. Since $\sigma^2_{A,p}$ and $\sigma^2_{G,q}$ are fixed at this step, we optimize $\sigma^2_\beta$ parameter to adjust for slight changes in heritability resulting from changes in $S$ and $\ell$ parameters. The main inflation parameter $\sigma_0^2$ is optimized at both steps. The secondary inflation parameter $\sigma^2_{0L}$, if requested by the user, is also optimized at both steps.

#### Simulations with pre-defined levels of fold enrichment

Let $G$ and $g$ be two regions in a genome, and $g$ be a subset of $G$. For example, $G$ can be the set of all genes, while $g$ be a subset of genes. Let $H_G$ and $H_g$ be the total heterozigosity (i.e. $2p(1-p)$ where $p$ is minor allele frequency) combined across all causal variants in regions $G$ and $g$, respectively.

Let effect size variance for causal SNPs in $G \backslash g$ be $\sigma_G^2$, while effect size variance for SNPs in $g$ be $(\sigma_G^2 + \sigma_g^2)$. Let's also assume that there are no causal variants outside $G$. Then total heritability of a trait simulated according to the above model is $h^2 = H_G\sigma_G^2 + H_g\sigma_g^2$. Under null model (no enrichment) let $\sigma^2_{null}$ be the effect size variance for causal variants in $G$. Then heritability under null model is $h^2 = H_G\sigma^2_{null}$.

Heritability within the region $g$ can be expressed as $h^2(g) = H_g(\sigma_g^2 + \sigma_G^2)$ (under

**Algorithm 1:** Adam algorithm for GSA-MiXeR inference

**Input:** $\alpha_1, \ldots, \alpha_N$: Stepsize for each epoch
**Input:** $\beta_1, \beta_2 \in [0, 1)$: Exponential decay rates for the moment estimates
**Input:** $f(\theta, X)$: Stochastic objective function with parameters $\theta$ on batch $X$
**Input:** $\theta_0$: Initial parameter vector
$m_0 \leftarrow 0$ (Initialize 1st moment vector)
$v_0 \leftarrow 0$ (Initialize 2nd moment vector)
$\leftarrow 0$ (Initialize timestep)
**for** $n$ ***in*** $1, \ldots, N$ **do**
  **for** $X$ ***in*** $shuffle(X_1, \ldots, X_{22})$ **do**
    $t \leftarrow t + 1$
    $g_t \leftarrow \nabla_\theta f_t(\theta_{t-1}, X)$ (Get gradients w.r.t. stochastic objective at time $t$)
    $m_t \leftarrow \beta_1 \cdot m_{t-1} + (1 - \beta_1) \cdot g_t$ (Update biased 1st moment estimate)
    $v_t \leftarrow \beta_2 \cdot v_{t-1} + (1 - \beta_2) \cdot g_t^2$ (Update biased 2nd raw moment estimate)
    $\hat{m}_t \leftarrow m_t/(1 - \beta_1^t)$ (Compute bias-corrected 1st moment estimate)
    $\hat{v}_t \leftarrow v_t/(1 - \beta_2^t)$ (Compute bias-corrected 2nd raw moment estimate)
    $\theta_t \rightarrow \theta_{t-1} - \alpha_n \cdot \hat{m}_t/(\sqrt{\hat{v}_t} + \epsilon)$ (Update parameters)
  **end**
**end**
**Result:** $\theta_t$ (Resulting parameters)

full model) or $H^2(g) = H_g \sigma^2_{null}$ (under null model). Therefore, fold enrichment of heritability is expected to be $f(g) = (\sigma^2_g + \sigma^2_G)/\sigma^2_{null}$.

Using the above equations, and assuming that $H_g, H_G, h^2$ and $f(g)$ are known parameters one can derive that

$$\begin{aligned} \sigma^2_G &= h^2 \frac{H_G - f(g)H_g}{H_G(H_G - H_g)} \\ \sigma^2_G + \sigma^2_g &= h^2 f(g)/H_G, \end{aligned} \tag{9}$$

(under a constraint that $f(g)$ does not exceed $H_G/H_g$ ratio). We will use the above formulas to simulate a trait with pre-defined heritability and fold enrichment in a region $g$ over a wider region $G$. Note the ratio has the following formula:

$$\frac{\sigma^2_G + \sigma^2_g}{\sigma^2_G} = f(g) \frac{H_G - H_g}{H_G - f(g)H_g}$$

Finally, $h^2(g) = H_g(\sigma^2_g + \sigma^2_G) = h^2 f(g) H_g / H_G$. This last formula can be applied also to individual genes within $g$, which can be written as $\sum_i H_i \beta_i^2$ across causal SNPs. After drawing $\beta_i$ from normal distribution we can re-scale $\beta_i$ by a constant factor to achieve expected $h^2(g)$.

### Extended authors list: *Mapping genomic loci implicates genes and synaptic biology in schizophrenia*, Nature, 2022.

Vassily Trubetskoy[1†], Antonio F Pardiñas[2†], Ting Qi[3,4], Georgia Panagiotaropoulou[1], Swapnil Awasthi[1], Tim B Bigdeli[5,6,7], Julien Bryois[8], Chia-Yen Chen[9,10,11], Charlotte A Dennison[2], Lynsey S Hall[2], Max Lam[12,13,14], Kyoko Watanabe[15], Oleksandr Frei[16,17,18], Tian Ge[11,12,19], Janet C Harwood[2], Frank Koopmans[20], Sigurdur Magnusson[21], Alexander L Richards[2], Julia Sidorenko[3], Yang Wu[3], Jian Zeng[3], Jakob Grove[22,23,24], Minsoo Kim[25], Zhiqiang Li[26,27], Georgios Voloudakis[28], Wen Zhang[28,29], Mark Adams[30], Ingrid Agartz[16,31,32], Elizabeth G Atkinson[10,12], Esben Agerbo[22,33], Mariam Al Eissa[34], Margot Albus[35], Madeline Alexander[36], Behrooz Z Alizadeha[37,38], Köksal Alptekin[39,40], Thomas D Als[22,23,24], Farooq Amin[41], Volker Arolt[42], Manuel Arrojo[43], Lavinia Athanasiu[16,17], Maria Helena Azevedo[44], Silviu A Bacanu[45], Nicholas J Bass[34], Martin Begemann[46], Richard A Belliveau[12], Judit Bene[47], Beben Benyamin[48,49,50], Sarah E Bergen[8], Giuseppe Blasi[51], Julio Bobes[52,53,54], Stefano Bonassi[55], Alice Braun[1], Rodrigo Affonseca Bressan[56,57], Evelyn J Bromet[58], Richard Bruggeman[37,59], Peter F Buckley[60], Randy L Buckner[61], Jonas Bybjerg-Grauholm[22,62], Wiepke Cahn[63,64], Murray J Cairns[65,66,67], Monica E Calkins[68], Vaughan J Carr[69,70,71], David Castle[72,73], Stanley V Catts[74,75], Kimberley D Chambert[12], Raymond CK Chan[76,77], Boris Chaumette [78,79], Wei Cheng[80], Eric FC Cheung[81], Siow Ann Chong[13,82], David Cohen[83,84,85], Angèle Consoli[83,84], Quirino Cordeiro[86], Javier Costas[87], Charles Curtis[88,89], Michael Davidson[90], Kenneth L Davis[91], Lieuwe de Haan[92,93], Franziska Degenhardt[94], Lynn E DeLisi[19,95], Ditte Demontis[22,23,24], Faith Dickerson[96], Dimitris Dikeos[97], Timothy Dinan[98,99], Srdjan Djurovic[100,101], Jubao Duan[102,103], Giuseppe Ducci[104], Frank Dudbridge[105], Johan G Eriksson[106,107,108], Lourdes Fañanás[109,110], Stephen V Faraone[111], Alessia Fiorentino[34], Andreas Forstner[94,112], Josef Frank[113], Nelson B Freimer[114,115], Menachem Fromer[116], Alessandra Frustaci[117], Ary Gadelha[56,57], Giulio Genovese[12], Elliot S Gershon[118], Marianna Giannitelli[83,84], Ina Giegling[119], Paola Giusti-Rodríguez[120], Stephanie Godard[121], Jacqueline I Goldstein[10], Javier González Peñas[110,122], Ana González-Pinto[110,123], Srihari Gopal[124], Jacob Gratten[3,125], Michael F Green[126,127], Tiffany A Greenwood[128], Olivier Guillin[129,130,131], Sinan Gülöksüz[132,133], Raquel E Gur[68], Ruben C Gur[68], Blanca Gutiérrez[134], Eric Hahn[135], Hakon Hakonarson[136], Vahram Haroutunian[91,137,138], Annette M Hartmann[119], Carol Harvey[72,139], Caroline Hayward[140], Frans A Henskens[141], Stefan Herms[142], Per Hoffmann[142], Daniel P Howrigan[10,143], Masashi Ikeda[144], Conrad Iyegbe[145], Inge Joa[146], Antonio Julià[147], Anna K Kähler[8], Tony Kam-Thong[148], Yoichiro Kamatani[149,150], Sena Karachanak-Yankova[151,152], Oussama Kebir[78], Matthew C Keller[153], Brian J Kelly[141], Andrey Khrunin[154], Sung-Wan Kim[155], Janis Klovins[156], Nikolay Kondratiev[157], Bettina Konte[119], Julia Kraft[1,158], Michiaki Kubo[159], Vaidutis Kučinskas[160], Zita Ausrele Kučinskiene[160], Agung Kusumawardhani[161], Hana Kuzelova-Ptackova[162], Stefano Landi[163], Laura C Lazzeroni[164,165], Phil H Lee[12,166], Sophie E Legge[2], Douglas S Lehrer[167], Rebecca Lencer[42], Bernard Lerer[168], Miaoxin Li[169], Jeffrey Lieberman[170], Gregory A Light[128,171], Svetlana Limborska[154], Chih-Min Liu[172,173], Jouko Lönnqvist[174,175], Carmel M Loughland[176], Jan Lubinski[177], Jurjen J Luykx[132,178,179,180], Amy Lynham[2], Milan Macek Jr[181], Andrew Mackinnon[182,183], Patrik KE Magnusson[8], Brion S Maher[184], Wolfgang Maier[185], Dolores Malaspina[91,186], Jacques Mallet[187], Stephen R Marder[188], Sara Marsal[147], Alicia R Martin[10,12,189], Lourdes Martorell[190], Manuel Mattheisen[23,191,192,193], Robert W McCarley[194,195], Colm McDonald[196], John J McGrath[33,197,198], Helena Medeiros[199,200], Sandra Meier[191,201], Bela Melegh[202],

Ingrid Melle[16,17], Raquelle I Mesholam-Gately[19,203], Andres Metspalu[204], Patricia T Michie[205], Lili Milani[204], Vihra Milanova[206], Marina Mitjans[46], Espen Molden[207,208], Esther Molina[209], María Dolores Molto[110,210,211], Valeria Mondelli[89,212], Carmen Moreno[110,122], Christopher P Morley[213], Gerard Muntané[190,214], Kieran C Murphy[215], Inez Myin-Germeys[216], Igor Nenadić[217,218], Gerald Nestadt[219], Liene Nikitina-Zake[156], Cristiano Noto[56,57], Keith H Nuechterlein[126], Niamh Louise O'Brien[34], F Anthony O'Neill[220], Sang-Yun Oh[221,222], Ann Olincy[223], Vanessa Kiyomi Ota[57,224], Christos Pantelis[139,225,226,227], George N Papadimitriou[97], Mara Parellada[110,122], Tiina Paunio[228,229], Renata Pellegrino[136], Sathish Periyasamy[197,230], Diana O Perkins[231], Bruno Pfuhlmann[232], Olli Pietiläinen[12,233,234], Jonathan Pimm[34], David Porteous[235], John Powell[236], Diego Quattrone[88,89,237], Digby Quested[238,239], Allen D Radant[240,241], Antonio Rampino[51], Mark H Rapaport[242], Anna Rautanen[148], Abraham Reichenberg[91], Cheryl Roe[243], Joshua L Roffman[244], Julian Roth[245], Matthias Rothermundt[42], Bart PF Rutten[132], Safaa Saker-Delye[246], Veikko Salomaa[247], Julio Sanjuan[110,211,248], Marcos Leite Santoro[57,224], Adam Savitz[124], Ulrich Schall[66,249], Rodney J Scott[65,250,251], Larry J Seidman[19,203], Sally Isabel Sharp[34], Jianxin Shi[252], Larry J Siever[91,253], Engilbert Sigurdsson[254,255], Kang Sim[256,257,258], Nora Skarabis[1], Petr Slominsky[154], Hon-Cheong So[259,260], Janet L Sobell[199], Erik Söderman[32], Helen J Stain[261,262], Nils Eiel Steen[17,263], Agnes A. Steixner-Kumar[46], Elisabeth Stögmann[264], William S Stone[265,266], Richard E Straub[267], Fabian Streit[113], Eric Strengman[268], T Scott Stroup[170], Mythily Subramaniam[13,82], Catherine A Sugar[126,269], Jaana Suvisaari[247], Dragan M Svrakic[270], Neal R Swerdlow[128], Jin P Szatkiewicz[120], Thi Minh Tam Ta[271,272], Atsushi Takahashi[132,273], Chikashi Terao[273], Florence Thibaut[274,275], Draga Toncheva[151,276], Paul A Tooney[65,66,67], Silvia Torretta[51], Sarah Tosato[277], Gian Battista Tura[278], Bruce I Turetsky[68], Alp Üçok[279], Arne Vaaler[280,281], Therese van Amelsvoort[89,132], Ruud van Winkel[132,282], Juha Veijola[283,284], John Waddington[285], Henrik Walter[286], Anna Waterreus[287,288], Bradley T Webb[45], Mark Weiser[289], Nigel M Williams[2], Stephanie H Witt[113], Brandon K Wormley[45], Jing Qin Wu[290], Zhida Xu[291], Robert Yolken[292], Clement C Zai[293,294], Wei Zhou[27], Feng Zhu[295,296], Fritz Zimprich[264], Eşref Cem Atbaşoğlu[186,297], Muhammad Ayub[298], Christian Benner[234], Alessandro Bertolino[51], Donald W Black[299], Nicholas J Bray[2], Gerome Breen[88], Nancy G Buccola[300], William F Byerley[301], Wei J Chen[302,303], C Robert Cloninger[270], Benedicto Crespo-Facorro[304,305], Gary Donohoe[196], Robert Freedman[223], Cherrie Galletly[306,307,308], Michael J Gandal[25], Massimo Gennarelli[309,310], David M Hougaard[22,62], Hai-Gwo Hwu[173,311], Assen V Jablensky[288], Steven A McCarroll[12], Jennifer L Moran[12,244], Ole Mors[22,312], Preben B Mortensen[22,33], Bertram Müller-Myhsok[313,314,315], Amanda L Neil[316], Merete Nordentoft[22,317], Michele T Pato[318,319], Tracey L Petryshen[166], Matti Pirinen[234,320,321], Ann E Pulver[219], Thomas G Schulze[193,322,323,324], Jeremy M Silverman[91,253], Jordan W Smoller[12,166], Eli A Stahl[116,325,326], Debby W Tsuang[240,241], Elisabet Vilella[190], Shi-Heng Wang[327], Shuhua Xu[328,329,330], Indonesia Schizophrenia Consortium[*], PsychENCODE[*], Psychosis Endophenotypes International Consortium[*], The SynGO Consortium[*], Rolf Adolfsson[331], Celso Arango[110,122], Bernhard T Baune[42,226,227], Sintia Iole Belangero[57,224], Anders D Børglum[22,23,24], David Braff[128,171], Elvira Bramon[332], Joseph D Buxbaum[91], Dominique Campion[129,130], Jorge A Cervilla[333], Sven Cichon[334,335,336], David A Collier[337], Aiden Corvin[338], Marta Di Forti[88,89,237], Enrico Domenici[341], Hannelore Ehrenreich[46], Valentina Escott-Price[342,343], Tõnu Esko[204,325], Ayman H Fanous[7,344,345], Anna Gareeva[346,347], Micha Gawlik[245], Pablo V Gejman[102,103], Michael Gill[338], Stephen J Glatt[348], Vera Golimbet[157], Kyung Sue Hong[349], Christina M Hultman[8], Steven E Hyman[12,233], Nakao Iwata[144], Erik G Jönsson[32,263], René S Kahn[63,91], James L Kennedy[293,294], Elza

Khusnutdinova[347,350], George Kirov[2], James A Knowles[351,352], Marie-Odile Krebs[78], Claudine Laurent-Levinson[83,84], Jimmy Lee[353,354], Todd Lencz[14,355,356], Douglas F Levinson[164], Qingqin S Li[124], Jianjun Liu[357,358], Anil K Malhotra[14,355,356], Dheeraj Malhotra[359], Andrew McIntosh[30], Andrew McQuillin[34], Paulo R Menezes[360], Vera A Morgan[287,288], Derek W Morris[196], Bryan J Mowry[197,230], Robin M Murray[89,361], Vishwajit Nimgaonkar[362], Markus M Nöthen[94], Roel A Ophoff[114,363,364], Sara A Paciga[365], Aarno Palotie[234,366,367], Carlos N Pato[318,319], Shengying Qin[27,368], Marcella Rietschel[113], Brien P Riley[45], Margarita Rivera[369,370], Dan Rujescu[119], Meram C Saka[297], Alan R Sanders[102,103], Sibylle G Schwab[371,372], Alessandro Serretti[373], Pak C Sham[374,375,376], Yongyong Shi[27,377], David St Clair[378], Hreinn Stefánsson[21], Kari Stefansson[21], Ming T Tsuang[379,380], Jim van Os[381,382], Marquis P Vawter[383], Daniel R Weinberger[267], Thomas Werge[384,385,386,387], Dieter B Wildenauer[388], Xin Yu[389,390], Weihua Yue[389,390,391], Peter A Holmans[2], Andrew J Pocklington[2], Panos Roussos[28,392], Evangelos Vassos[88,89,393], Matthijs Verhage[394,395], Peter M Visscher[3], Jian Yang[3,4,396], Danielle Posthuma[395], Ole A Andreassen[16,17], Kenneth S Kendler[45], Michael J Owen[2], Naomi R Wray[3,197], Mark J Daly[10,143,234], Hailiang Huang[10,12,189], Benjamin M Neale[10,12], Patrick F Sullivan[8,120,231], Stephan Ripke[1,10,367‡], James TR Walters[2‡], Michael C O'Donovan[2‡], Schizophrenia Working Group of the Psychiatric Genomics Consortium[*]

[1]Department of Psychiatry and Psychotherapy, Charité - Universitätsmedizin, Berlin, Germany. [2]MRC Centre for Neuropsychiatric Genetics and Genomics, Division of Psychiatry and Clinical Neurosciences, Cardiff University, Cardiff, UK. [3]Institute for Molecular Bioscience, University of Queensland, Brisbane QLD, Australia. [4]School of Life Sciences, Westlake University, Hangzhou Zhejiang, China. [5]Department of Psychiatry and the Behavioral Sciences, State University of New York, Downstate Medical Center, New York NY, USA. [6]Institute for Genomic Health, SUNY Downstate Medical Center, New York NY, USA. [7]Department of Psychiatry, Veterans Affairs New York Harbor Healthcare System, New York NY, USA. [8]Department of Medical Epidemiology and Biostatistics, Karolinska Institutet, Stockholm, Sweden. [9]Biogen, Cambridge MA, USA. [10]Analytic and Translational Genetics Unit, Massachusetts General Hospital, Boston, MA USA. [11]Psychiatric and Neurodevelopmental Genetics Unit, Center for Genomic Medicine, Massachusetts General Hospital, Boston MA, USA. [12]Stanley Center for Psychiatric Research, Broad Institute of MIT and Harvard, Cambridge MA, USA. [13]Research Division, Institute of Mental Health, Singapore, Republic of Singapore. [14]Division of Psychiatry Research, Zucker Hillside Hospital, Glen Oaks NY, USA. [15]Department of Complex Trait Genetics, Center for Neurogenomics and Cognitive Research, Amsterdam Neuroscience, Vrije Universiteit Amsterdam, 1081 HV, Amsterdam, The Netherlands. [16]NORMENT Centre, Division of Mental Health and Addiction, University of Oslo, Oslo, Norway. [17]Division of Mental Health and Addiction, Oslo University Hospital, Oslo, Norway. [18]Center for Bioinformatics, Department of Informatics, University of Oslo, Oslo, Norway. [19]Department of Psychiatry, Harvard Medical School, Boston MA, USA. [20]Department of Molecular and Cellular Neurobiology, Center for Neurogenomics and Cognitive Research, Faculty of Science, Amsterdam Neuroscience, Vrije Universiteit, Amsterdam, The Netherlands. [21]deCODE Genetics/Amgen Inc., Reykjavik, Iceland. [22]The Lundbeck Foundation Initiative for Integrative Psychiatric Research (iPSYCH), Aarhus, Denmark. [23]Department of Biomedicine and Centre for Integrative Sequencing (iSEQ), Aarhus University, Aarhus, Denmark. [24]Center for

Genomics and Personalized Medicine, Aarhus, Denmark. [25]Department of Psychiatry, Semel Institute, David Geffen School of Medicine, University of California, Los Angeles CA, USA. [26]Affiliated Hospital of Qingdao University & Biomedical Sciences Institute, Qingdao University, Qingdao Shi, China. [27]Bio-X Institutes, Key Laboratory for the Genetics of Developmental and Neuropsychiatric Disorders (Ministry of Education), Collaborative Innovation Center for Brain Science, Shanghai Jiao Tong University, Shanghai, China. [28]Department of Psychiatry, Pamela Sklar Division of Psychiatric Genomics, Friedman Brain Institute, Department of Genetics and Genomic Science and Institute for Data Science and Genomic Technology, Icahn School of Medicine at Mount Sinai, New York NY, USA. [29]Department of Genetics & Genomic Sciences and Institute for Genomics and Multiscale Biology, Icahn School of Medicine at Mount Sinai, New York NY, USA. [30]Division of Psychiatry, Centre for Clinical Brain Sciences, University of Edinburgh, Royal Edinburgh Hospital, Edinburgh, UK. [31]Department of Psychiatric Research, Diakonhjemmet Hospital, Oslo, Norway. [32]Centre for Psychiatry Research, Department of Clinical Neuroscience, Karolinska Institutet & Stockholm Health Care Services, Stockholm Region, Stockholm, Sweden. [33]National Centre for Register-based Research, Aarhus University, Aarhus, Denmark. [34]Molecular Psychiatry Laboratory, Division of Psychiatry, University College London, London, UK. [35]Comedicum Lindwurmhof, Munich, Germany. [36]Center for Depression, Anxiety and Stress Research, McLean Hospital, Belmont MA, USA. [37]University Medical Center Groningen, University Center for Psychiatry, Rob Giel Research Center, Unviersity of Groningen, Groningen, The Netherlands. [38]Department of Epidemiology, University Medical Center Groningen, Unviersity of Groningen, Groningen, The Netherlands. [39]Department of Psychiatry, Dokuz Eylül University School of Medicine, Izmir, Turkey. [40]Department of Neuroscience, Dokuz Eylül University Graduate School of Health Sciences, Izmir, Turkey. [41]Department of Psychiatry and Behavioral Sciences, Emory University, Atlanta GA, USA. [42]Department of Psychiatry, University of Münster, Münster, Germany. [43]Servizo de Psiquiatría, Complexo Hospitalario Universitario de Santiago de Compostela, Servizo Galego de Saúde (SERGAS), Santiago de Compostela Galicia, Spain. [44]Institute of Medical Psychology, Faculty of Medicine, University of Coimbra, Coimbra, Portugal. [45]Virginia Institute for Psychiatric and Behavioral Genetics, Department of Psychiatry, Virginia Commonwealth University, Richmond VA, USA. [46]Clinical Neuroscience, Max Planck Institute of Experimental Medicine, Göttingen, Germany. [47]Department of Medical Genetics, Medical School, University of Pécs, Pécs, Hungary. [48]Australian Centre for Precision Health, University of South Australia Cancer Research Institute, University of South Australia, Adelaide SA, Australia. [49]UniSA Allied Health & Human Performance, University of South Australia, Adelaide SA, Australia. [50]South Australian Health and Medical Research Institute, Adelaide SA, Australia. [51]Department of Basic Medical Science, Neuroscience and Sense Organs, University of Bari 'Aldo Moro', Bari, Italy. [52]Área de Psiquiatría-Universidad de Oviedo, Hospital Universitario Central de Asturias (HUCA), Asturias, Spain. [53]Instituto de Investigación Sanitaria del Principado de Asturias (ISPA), Asturias, Spain. [54]Centro de Investigación Biomédica en Red de Salud Mental, Oviedo, Asturias, Spain. [55]Unit of Clinical and Molecular Epidemiology, IRCCS San Raffaele Pisana, and San Raffaele University, Rome, Italy. [56]Department of Psychiatry, Universidade Federal de Sao Paulo, Sao Paulo SP, Brazil. [57]Laboratory of Integrative Neuroscience, Universidade Federal de Sao Paulo, Sao Paulo SP, Brazil. [58]Department of Psychiatry and Behavioural Health, Stony Brook University, Stony Brook NY, USA. [59]University of Groningen, Department of Clinical and Developmental

Neuropsychology, Groningen, The Netherlands. [60]School of Medicine, Virginia Commonwealth University,, Richmond VA, USA. [61]Department of Psychology, Harvard University, Cambridge MA, USA. [62]Center for Neonatal Screening, Department for Congenital Disorders, Statens Serum Institut, Copenhagen, Denmark. [63]University Medical Center Utrecht, Department of Psychiatry, Rudolf Magnus Institute of Neuroscience, Utrecht, The Netherlands. [64]Altrecht, General Menthal Health Care, Utrecht, The Netherlands. [65]School of Biomedical Sciences and Pharmacy, University of Newcastle, Callaghan NSW, Australia. [66]Hunter Medical Research Institute, Newcastle NSW, Australia. [67]Centre for Brain & Mental Health Research, The University of Newcastle, Callaghan NSW, Australia. [68]Department of Psychiatry, University of Pennsylvania, Philadelphia PA, USA. [69]School of Psychiatry, University of New South Wales, Sydney NSW, Australia. [70]Department of Psychiatry, Monash University, Melbourne, Australia. [71]Neuroscience Research Australia, Sydney, Australia. [72]Department of Psychiatry, The University of Melbourne, Parkville VIC, Australia. [73]St Vincent's Hospital, 41 Victoria Parade, Fitzroy VIC, Australia. [74]Brain and Mind Centre, The University of Sydney, Sydney NSW, Australia. [75]School of Medicine, University of Queensland, Herston QLD, Australia. [76]Institute of Psychology, Chinese Academy of Science, Beijing, China. [77]Department of Psychology, University of Chinese Academy of Sciences, Beijing, China. [78]INSERM U1266, Institute of Psychiatry and Neuroscience of Paris, Université de Paris, GHU Paris Psychiatrie & Neurosciences, Paris, France. [79]Department of Psychiatry, McGill University, Montreal, Canada. [80]Department of Computer Science, University of North Carolina, Chapel Hill NC, USA. [81]Castle Peak Hospital, Hong Kong, China. [82]Saw Swee Hock School of Public Health, National University of Singapore, Singapore, Republic of Singapore. [83]Faculté de Médecine Sorbonne Université, Groupe de Recherche Clinique n°15 - Troubles Psychiatriques et Développement (PSYDEV), Department of Child and Adolescent Psychiatry, Hôpital Universitaire de la Pitié-Salpêtrière, Paris, France. [84]Centre de Référence des Maladies Rares à Expression Psychiatrique, Department of Child and Adolescent Psychiatry, AP-HP Sorbonne Université, Hôpital Universitaire de la Pitié-Salpêtrière, Paris, France. [85]Institut des Systèmes Intelligents et de Robotique (ISIR), CNRS UMR7222, Sorbonne Université, Campus Pierre et Marie Curie, Faculté des Sciences et Ingénierie, Paris, France. [86]Department of Psychiatry, Irmandade da Santa Casa de Misericórdia de São Paulo, Rua Dona Veridiana, São Paulo SP, Brazil. [87]Instituto de Investigación Sanitaria (IDIS) de Santiago de Compostela, Complexo Hospitalario Universitario de Santiago de Compostela (CHUS), Servizo Galego de Saúde (SERGAS), Santiago de Compostela Galicia, Spain. [88]Social, Genetic and Developmental Psychiatry Centre, Institute of Psychiatry, Psychology and Neuroscience, King's College London, London, UK. [89]National Institute for Health Research (NIHR) Maudsley Biomedical Research Centre at South London and Maudsley NHS Foundation Trust and King's College London, London, UK. [90]University of Nicosia Medical School, Nicosia, Cyprus. [91]Department of Psychiatry, Icahn School of Medicine at Mount Sinai, New York NY, USA. [92]Department of Psychiatry, Academic Medical Centre, University of Amsterdam, Amsterdam, The Netherlands. [93]Arkin, Institute for Mental Health, Amsterdam, The Netherlands. [94]Institute of Human Genetics, University of Bonn, Bonn, Germany. [95]Cambridge Health Alliance, Cambridge MA, USA. [96]Sheppard Pratt Health System, Baltimore MD, USA. [97]First Department of Psychiatry, Medical School, National and Kapodistrian University of Athens, Eginition Hospital, Athens, Greece. [98]Department of Psychiatry and Neurobehavioural Sciences, University College Cork, Cork, Ireland. [99]APC Microbiome Ireland, University College Cork, Cork, Ireland.

[100]NORMENT Centre, Department of Clinical Science, University of Bergen, Bergen, Norway. [101]Department of Medical Genetics, Oslo University Hospital, Oslo, Norway. [102]Center for Psychiatric Genetics, NorthShore University HealthSystem, Evanston IL, USA. [103]Department of Psychiatry and Behavioral Neurosciences, The University of Chicago, Chicago IL, USA. [104]Department of Mental Health, ASL Rome 1, Rome, Italy. [105]Department of Health Sciences, University of Leicester, Leicester, UK. [106]Department of General Practice and Primary Health Care, University of Helsinki and Helsinki University Hospital, Helsinki, Finland. [107]Folkhälsan Research Center, Helsinki, Finland. [108]Department of Obstetrics & Gynecology, Yong Loo Lin School of Medicine, National University of Singapore, Singapore, Republic of Singapore. [109]Department of Evolutionary Biology, Ecology and Environmental Sciences, Faculty of Biology, University of Barcelona, Barcelona, Spain. [110]Centro de Investigación Biomédica en Red en Salud Mental (CIBERSAM), Madrid, Spain. [111]Departments of Psychiatry and Neuroscience and Physiology, SUNY Upstate Medical University, Syracuse NY, USA. [112]Centre for Human Genetics, University of Marburg, Marburg, Germany. [113]Department of Genetic Epidemiology in Psychiatry, Central Institute of Mental Health, Medical Faculty Mannheim, University of Heidelberg, Mannheim, Germany. [114]Department of Human Genetics, David Geffen School of Medicine, University of California, Los Angeles BA, USA. [115]Department of Psychiatry and Biobehavioral Sciences, University of California, Los Angeles, Los Angeles CA, USA. [116]Division of Psychiatric Genomics, Department of Psychiatry, Icahn School of Medicine at Mount Sinai, New York NY, USA. [117]Barnet, Enfield and Haringey Mental Health NHS Trust, St.Ann's Hospital, London, UK. [118]Departments of Psychiatry and Human Genetics, University of Chicago, Chicago IL, USA. [119]Department of Psychiatry and Psychotherapy, Medical University of Vienna, Vienna, Austria. [120]Department of Genetics, University of North Carolina, Chapel Hill NC, USA. [121]Departments of Psychiatry and Human and Molecular Genetics, INSERM, Institut de Myologie, Hôpital de la Pitiè-Salpêtrière, Paris, France. [122]Department of Child and Adolescent Psychiatry, Hospital General Universitario Gregorio Marañón, School of Medicine, Universidad Complutense, Investigación Sanitaria del Hospital Gregorio Marañón, Madrid, Spain. [123]BIOARABA Health Research Institute. OSI Araba. University Hospital, University of the Basque Country, Vitoria, Spain. [124]Neuroscience Therapeutic Area, Janssen Research and Development, Titusville NJ, USA. [125]Mater Research Institute, University of Queensland, Brisbane QLD, Australia. [126]Department of Psychiatry and Biobehavioral Sciences, Geffen School of Medicine, University of California Los Angeles, Los Angeles CA, USA. [127]VA Greater Los Angeles Healthcare System, Los Angeles CA, USA. [128]Department of Psychiatry, University of California San Diego, La Jolla CA, USA. [129]INSERM, Rouen, France. [130]Centre Hospitalier du Rouvray, Rouen, France. [131]UFR santé, Université de Rouen Normandie, Rouen, France. [132]Department of Psychiatry and Neuropsychology, School for Mental Health and Neuroscience, Maastricht University Medical Centre, Maastricht, The Netherlands. [133]Department of Psychiatry, Yale School of Medicine, New Haven CT, USA. [134]Department of Psychiatry, Faculty of Medicine and Biomedical Research Centre (CIBM), University of Granada, Granada, Spain. [135]Department of Psychiatry, Charité - Universitätsmedizin, Campus Benjamin Franklin, Berlin, Germany. [136]Children's Hospital of Philadelphia, Leonard Madlyn Abramson Research Center, Philadelphia PA , USA. [137]Department of Neuroscience, Icahn School of Medicine at Mount Sinai, New York NY, USA. [138]Mental Illness Research Clinical and Education Center (MIRECC), JJ Peters VA Medical Center, New York NY, USA. [139]NorthWestern Mental Health,

Melbourne VIC, Australia. [140]MRC Human Genetics Unit, University of Edinburgh, Institute of Genetics and Molecular Medicine, Western General Hospital, Edinburgh, UK. [141]School of Medicine and Public Health, University of Newcastle, Newcastle NSW, Australia. [142]Division of Medical Genetics, Department of Biomedicine, University of Basel, Basel, Switzerland. [143]Broad Institute of MIT and Harvard, Cambridge MA, USA. [144]Department of Psychiatry, Fujita Health University School of Medicine, Toyoake Aichi, Japan. [145]Department of Psychosis Studies, Institute of Psychiatry, Psychology and Neuroscience, King's College London, London, UK. [146]Regional Centre for Clinical Research in Psychosis, Department of Psychiatry, Stavanger University Hospital, Stavanger, Norway. [147]Rheumatology Research Group, Vall d'Hebron Research Institute, Barcelona, Spain. [148]Roche Pharma Research and Early Development, Pharmaceutical Sciences, Roche Innovation Center Basel, F. Hoffman-La Roche Ltd, Basel, Switzerland. [149]Laboratory of Complex Trait Genomics, Department of Computational Biology and Medical Sciences, Graduate School of Frontier Sciences, The University of Tokyo, Tokyo, Japan. [150]Laboratory for Statistical Analysis, RIKEN Center for Integrative Medical Sciences, Yokohama Kanagawa, Japan. [151]Department of Medical Genetics, Medical University, Sofia, Bulgaria. [152]Department of Genetics, Faculty of Biology, Sofia University "St. Kliment Ohridski", Sofia, Bulgaria. [153]Institute for Behavioural Genetics, University of Colorado Boulder, Boulder CO, USA. [154]Institute of Molecular Genetics of National Research Centre "Kurchatov Institute", Moscow, Russia. [155]Department of Psychiatry, Chonnam National University Medical School, Gwangju, Korea. [156]Latvian Biomedical Research and Study Centre, Riga, Latvia. [157]Mental Health Research Center, Moscow, Russian Federation. [158]Berlin School of Mind and Brain, Humboldt-Universität zu Berlin, Berlin, Germany. [159]RIKEN Center for Integrative Medical Sciences, Yokohama Kanagawa, Japan. [160]Faculty of Medicine, Vilnius University, Vilnius, Lithuania. [161]Psychiatry Department, University of Indonesia - Cipto Mangunkusumo National General Hospital, Jakarta Pusat, DKI Jakarta, Jakarta, Indonesia. [162]Department of Psychiatry, 1st Faculty of Medicine and General University Hospital, Prague, Czech Republic. [163]Dipartimento di Biologia, Universita' di Pisa, Pisa, Italy. [164]Departments of Psychiatry and Behavioral Sciences, Stanford University, Stanford CA, USA. [165]Department of Biomedical Data Science, Stanford University, Stanford CA, USA. [166]Psychiatric and Neurodevelopmental Genetics Unit, Department of Psychiatry and Center for Genomic Medicine, Massachusetts General Hospital, Harvard Medical School, Boston MA, USA. [167]Department of Psychiatry, Wright State University, 3640 Colonel Gleen Hwy, Dayton OH, USA. [168]Department of Psychiatry, Hadassah-Hebrew University Medical Center, Jerusalem, Israel. [169]Zhongshan School of Medicine and Key Laboratory of Tropical Diseases Control (SYSU), Sun Yat-sen University, Guangzhou, China. [170]Department of Psychiatry, Columbia University, New York NY, USA. [171]VISN 22, Mental Illness Research, Education & Clinical Center (MIRECC), VA San Diego Healthcare System, San Diego CA, USA. [172]Department of Psychiatry, National Taiwan University Hospital, Taipei, Taiwan. [173]Neurobiology and Cognitive Science Center, National Taiwan University, Taipei, Taiwan. [174]Mental Health Unit, Department of Public Health Solutions, National Institute for Health and Welfare, Helsinki, Finland. [175]Department of Psychiatry, University of Helsinki, Helsinki, Finland. [176]Hunter New England Health & University of Newcastle, Newcastle NSW, Australia. [177]Department of Genetics and Pathology, International Hereditary Cancer Center, Pomeranian Medical University in Szczecin, Szczecin, Poland. [178]Department of Psychiatry, UMC Utrecht Brain Center, University Medical Centre Utrecht, Utrecht University, Utrecht, The

Netherlands. [179]Department of Translational Neuroscience, UMC Utrecht Brain Center, University Medical Center, Utrecht, Utrecht University, Utrecht, The Netherlands. [180]Second opinion outpatient clinic, GGNet mental health, Warnsveld, The Netherlands. [181]Department of Biology and Medical Genetics, 2nd Faculty of Medicine and University Hospital Motol, 150 06 Prague, Czech Republic. [182]Black Dog Institute, University of new South Wales, Randwick NSW, Australia. [183]Melbourne School of Population and Global health, University of Melbourne VIC, Australia. [184]Department of Mental Health, Bloomberg School of Public Health, Johns Hopkins University, Baltimore MD, USA. [185]Department for Neurodegenerative Diseases and Geriatric Psychiatry, University Hospital Bonn, Bonn, Germany . [186]Department of Genetics & Genomics, Icahn School of Medicine at Mount Sinai, Mount Sinai, New York NY, USA. [187]Asfalia Biologics, iPEPS-ICM, Hôpital Universitaire de la Pitié-Salpêtrière, Paris, France. [188]Semel Institute for Neurosciene, University of California, Los Angeles CA, USA. [189]Department of Medicine, Harvard Medical School, Boston MA, USA. [190]Hospital Universitari Institut Pere Mata, IISPV, Universitat Rovira i Virgili, CIBERSAM, Reus, Spain. [191]Department of Psychiatry, Dalhousie University, Halifax NS, Canada. [192]Department of Community Health and Epidemiology, Dalhousie University, Halifax Nova Scotia, Canada. [193]Institute of Psychiatric Phenomics and Genomics (IPPG), University Hospital, LMU Munich, Munich, Germany. [194]VA Boston Health Care System, Brockton MA, USA. [195]Deceased. [196]Centre for Neuroimaging, Cognition and Genomics (NICOG), National University of Ireland Galway, Galway, Ireland. [197]Queensland Brain Institute, The University of Queensland, Brisbane QLD, Australia. [198]Queensland Centre for Mental Health Research, The Park Centre for Mental Health, Queensland, Australia. [199]Department of Psychiatry and the Behavioral Sciences, Keck School of Medicine, University of Southern California, Los Angeles CA, USA. [200]College of Medicine, SUNY Downstate Health Sciences University, New York NY, USA. [201]Department of Biomedicine, Aarhus University, Aarhus, Denmark. [202]Department of Medical Genetics, University of Pécs, Pécs, Hungary. [203]Massachusetts Mental Health Center Public Psychiatry Division of the Beth Israel Deaconess Medical Center, Boston MA, USA. [204]Estonian Genome Center, Institute of Genomics, University of Tartu, Tartu, Estonia. [205]School of Psychology, University of Newcastle, Newcastle NSW, Australia. [206]First Psychiatric Clinic, Medical University, Sofia, Bulgaria. [207]Department of Pharmacy, University of Oslo, Oslo, Norway. [208]Center for Psychopharmacology, Diakonhjemmet Hospital, Oslo, Norway. [209]Department of Nursing, Faculty of Health Sciences and Biomedical Research Centre (CIBM), University of Granada, Granada, Spain. [210]Department of Genetics, Faculty of Biological Sciences, Campus of Burjassot, Universidad de Valencia, Valencia, Spain. [211]Biomedical Research Institute INCLIVA, Valencia, Spain. [212]Department of Psychological Medicine, Institute of Psychiatry, Psychology, and Neuroscience, King's College London, London, UK. [213]Departments of Public Health and Preventive Medicine, Family Medicine, and Psychiatry and Behavioral Sciences, State University of New York, Upstate Medical University, Syracuse NY, USA. [214]Institut de Biologia Evolutiva (UPF-CSIC), Departament de Ciències Experimentals i de la Salut, Universitat Pompeu Fabra, PRBB, Barcelona, Spain. [215]Department of Psychiatry, Royal College of Surgeons in Ireland, Dublin, Ireland. [216]Department for Neurosciences, Center for Contextual Psychiatry, KU Leuven, Leuven, Belgium. [217]Cognitive Neuropsychiatry Lab, Department of Psychiatry and Psychotherapy, Philipps Universität Marburg, Marburg, Germany. [218]Department of Psychiatry and Psychotherapy, Jena University Hospital, Jena, Germany. [219]Department of Psychiatry and

Behavioral Sciences, Johns Hopkins University School of Medicine, Baltimore MD, USA. [220]Centre for Public Health, Institute of Clinical Sciences, Queen's University Belfast, Belfast, UK. [221]Department of Statistics and Applied Probability, University of California at Santa Barbara, Santa Barbara CA, USA. [222]Computational Research Division, Lawrence Berkeley National Laboratory, Berkeley CA, USA. [223]Department of Psychiatry, University of Colorado Denver, Aurora CO, USA. [224]Department of Morphology and Genetics, Universidade Federal de Sao Paulo, Laboratorio de Genetica, Sao Paulo SP, Brazil. [225]Melbourne Neuropsychiatry Centre, University of Melbourne & Melbourne Health, Melbourne VIC, Australia. [226]The Florey Institute of Neuroscience and Mental Health, The University of Melbourne, Parkville, VIC, Australia. [227]Department of Psychiatry, Melbourne Medical School, The University of Melbourne, Parkville VIC, Australia. [228]Department of Public Health Solutions, Genomics and Biomarkers Unit, National Institute for Health and Welfare, Helsinki, Finland. [229]Department of Psychiatry and SleepWell Research Program, Faculty of Medicine, University of Helsinki and Helsinki University Central Hospital, Helsinki, Finland. [230]Queensland Centre for Mental Health Research, The University of Queensland, Brisbane QLD, Australia. [231]Department of Psychiatry, University of North Carolina, Chapel Hill NC, USA. [232]Clinic of Psychiatry and Psychotherapy, Weißer Hirsch, Dresden, Germany. [233]Department of Stem Cell and Regenerative Biology, Harvard University, Cambridge MA, USA. [234]Institute for Molecular Medicine Finland (FIMM), University of Helsinki, Helsinki, Finland. [235]Centre for Genomic and Experimental Medicine, Institute of Genetics and Molecular Medicine, University of Edinburgh, Western General Hospital, Edinburgh, UK. [236]Department of Basic and Clinical Neuroscience, Institute of Psychiatry, Psychology and Neuroscience, King's College London, London, UK. [237]South London and Maudsley NHS Mental Health Foundation Trust, London, UK. [238]Oxford Health NHS Foundation Trust, Warneford Hospital, Oxford, UK. [239]Department of Psychiatry, University of Oxford, Oxford, UK. [240]Department of Psychiatry and Behavioral Sciences, University of Washington, Seattle WA, USA. [241]VA Puget Sound Health Care System, Seattle WA, USA. [242]Huntsman Mental Health Institute, Department of Psychiatry, University of Utah School of Medicine, Salt Lake City UT, USA. [243]SUNY Upstate Medical University, Syracuse NY, USA. [244]Department of Psychiatry, Massachusetts General Hospital, Boston MA, USA. [245]Department of Psychiatry, Psychosomatics and Psychotherapy, Julius-Maximilians-Universität Würzburg, Wuerzburg, Germany. [246]Généthon, Evry, France. [247]THL-Finnish Institute for Health and Welfare, Helsinki, Finland. [248]Department of Psychiatry, School of Medicine, University of Valencia, Hospital Clínico Universitario de Valencia, Spain. [249]Priority Centre for Brain & Mental Health Research, The University of Newcastle, Mater Hospital, McAuley Centre, Waratah NSW, Australia. [250]Division of Molecular Medicine, NSW Health Pathology North, Newcastle, NSW, Australia. [251]Hunter Medical Research Institute, Newcastle NSW, Australia. [252]Division of Cancer Epidemiology and Genetics, National Cancer Institute, Bethesda MD, USA. [253]James J. Peters VA Medical Center, New York NY, USA. [254]Faculty of Medicine, University of Iceland, Reykjavik, Iceland. [255]Department of Psychiatry, Landspitali University hospital, Reykjavik, Iceland. [256]West Region, Institute of Mental Health, Singapore, Singapore. [257]Yoo Loo Lin School of Medicine, National University of Singapore, Singapore, Singapore. [258]Lee Kong Chian School of Medicine, Nanyang Technological University, Singapore, Singapore. [259]School of Biomedical Sciences, The Chinese University of Hong Kong, Hong Kong, China. [260]Department of Psychiatry, The Chinese University of Hong Kong, Hong Kong, China.

[261]School of Social and Health Sciences, Leeds Trinity University, Leeds, UK. [262]TIPS - Network for Clinical Research in Psychosis; Stavanger University Hospital, Stavanger, Norway. [263]NORMENT Centre, Institute of Clinical Medicine, University of Oslo, Oslo, Norway. [264]Department of Neurology, Medical University of Vienna, Vienna, Austria. [265]Harvard Medical School Department of Psychiatry at Beth Israel Deaconess Medical Center, Boston MA, USA. [266]Massachusetts Mental Health Center, Boston MA, USA. [267]Lieber Institute for Brain Development, Baltimore MD, USA. [268]Department of Medical Genetics, University Medical Centre Utrecht, Utrecht, The Netherlands. [269]Department of Biostatistics, Fielding School of Public Health, University of California Los Angeles, Los Angeles CA, USA. [270]Department of Psychiatry, Washington University, St. Louis MO, USA. [271]Department of Psychiatry, Campus Benjamin Franklin, Charité – Universitätsmedizin Berlin, Berlin, Germany. [272]Berlin Institute of Health (BIH), Berlin, Germany. [273]Laboratory for Statistical and Translational Genetics, RIKEN Center for Integrative Medical Sciences, Yokohama Kanagawa, Japan. [274]Université de Paris, Faculté de médecine, Hôpital Cochin-Tarnier, Paris, France. [275]INSERM U1266, Institut de psychiatrie et de neurosciences, Paris, France. [276]Bulgarian Academy of Science, Sofia, Bulgaria. [277]Department of Neuroscience, Biomedicine and Movement Sciences, Section of Psychiatry, University of Verona, Verona, Italy. [278]Psychiatry Unit, IRCCS Istituto Centro San Giovanni di Dio Fatebenefratelli, Brescia, Italy. [279]Department of Psychiatry, Faculty of Medicine, Istanbul University, Istanbul, Turkey. [280]Division of Mental Health, St. Olav's Hospital, Trondheim University Hospital, Trondheim, Norway. [281]Department of Mental Health, Norwegian University of Science and Technology, Trondheim, Norway. [282]KU Leuven, Department of Neurosciences, Center for Clinical Psychiatry, Leuven, Belgium. [283]Department of Psychiatry, Research Unit of Clinical Neuroscience, University of Oulu, Oulu, Finland. [284]Medical Research Center Oulu, Oulu University Hospital and University of Oulu, Oulu, Finland. [285]Molecular and Cellular Therapeutics, Royal College of Surgeons in Ireland, Dublin, Ireland. [286]Department for Psychiatry and Psychotherapy, CCM
Charité Universitätsmedizin Berlin, corporate member of Freie Universität Berlin, Humboldt-Universität zu Berlin, and Berlin Institute of Health, Berlin, Germany. [287]Neuropsychiatric Epidemiology Research Unit, School of Population and Global Health, University of Western Australia, Perth WA, Australia. [288]Centre for Clinical Research in Neuropsychiatry, The University of Western Australia, Perth WA, Australia. [289]Sheba Medical Center, Tel Hashomer, Israel. [290]School of Life and Environmental Sciences, University of Sydney, Sydney NSW, Australia. [291]Department of Psychiatry, GGz Centraal, The Netherlands. [292]Stanley Neurovirology Laboratory, Johns Hopkins University School of Medicine, Baltimore MD, USA. [293]Campbell Family Mental Health Research Institute, Centre for Addiction and Mental Health, Toronto ON, Canada. [294]Department of Psychiatry, University of Toronto, Toronto, ON, Canada. [295]Department of Psychiatry, The First Affiliated Hospital of Xi'an Jiaotong University, Xi'an, China. [296]Center for Translational Medicine, The First Affiliated Hospital of Xi'an Jiaotong University, Xi'an, China. [297]Department of Psychiatry, School of Medicine, Ankara University, Ankara, Turkey. [298]Department of Psychiatry, Queens University Kingston, 191 Portsmouth Avenue, Kingston ON, Canada. [299]Department of Psychiatry, University of Iowa Carver College of Medicine, Iowa City IA, USA. [300]School of Nursing, Louisiana State University Health Sciences Center, New Orleans LA, USA. [301]Department of Psychiatry, University of California San Francisco, San Francisco CA, USA. [302]Center for

Neuropsychiatric Research, National Health Research Institutes, Zhunan Town, Taiwan. [303]Institute of Epidemiology and Preventive Medicine, College of Public Health, National Taiwan University, Taipei, Taiwan. [304]University of Sevilla, CIBERSAM, Sevilla, Spain. [305]Hospital Universitario Virgen del Rocio, Department of Psychiatry, Universidad del Sevilla, Sevilla, Spain. [306]Discipline of Psychiatry, Adelaide Medical School, University of Adelaide, Adelaide SA, Australia. [307]Ramsay Health Care (SA) Mental Health, Ramsey MN, USA. [308]Northern Adelaide Local Health Network. [309]Department of Molecular and Translational Medicine, University of Brescia, Brescia, Italy. [310]Genetic Unit, IRCCS Istituto Centro San Giovanni di Dio Fatebenefratelli, Brescia, Italy. [311]Department of Psychiatry, College of Medicine and National Taiwan University Hospital, National Taiwan University, Taipei, Taiwan. [312]Psychosis Research Unit, Aarhus University Hospital, Aarhus, Denmark. [313]Max Planck Institute of Psychiatry, Munich, Germany. [314]Munich Cluster for Systems Neurology, Munich, Germany. [315]Department of Health Data Science, University of Liverpool, Liverpool, UK. [316]Menzies Institute for Medical Research, University of Tasmania, Hobart TAS, Australia. [317]Mental Health Services in the Capital Region of Denmark, Mental Health Center Copenhagen, University of Copenhagen, Copenhagen, Denmark. [318]Rutgers University, Robert Wood Johnson Medical School, New Brunswick NJ, USA. [319]Rutgers University, New Jersey Medical School, Newark NJ, USA. [320]Department of Mathematics and Statistics, University of Helsinki, Helsinki, Finland. [321]Department of Public Health, University of Helsinki, Helsinki, Finland. [322]Department of Psychiatry and Behavioral Sciences, SUNY Upstate Medical University, Syracuse NY, USA. [323]Department of Psychiatry and Psychotherapy, University Medical Center Göttingen, Göttingen, Germany. [324]Department of Psychiatry and Behavioral Sciences, The Johns Hopkins University, Baltimore, MD, USA. [325]Program in Medical and Population Genetics, The Broad Institute of MIT and Harvard, Cambridge MA, USA. [326]Regeneron Genetics Center, Orange CA, USA. [327]College of Public Health, China Medical University, Taichung, Taiwan. [328]State Key Laboratory of Genetic Engineering and Ministry of Education (MOE) Key Laboratory of Contemporary Anthropology, Collaborative Innovation Center of Genetics and Development, Human Phenome Institute, School of Life Sciences, Fudan University, Shanghai, China. [329]School of Life Science and Technology, ShanghaiTech University, Shanghai, China. [330]Center for Excellence in Animal Evolution and Genetics, Chinese Academy of Sciences, Kunming, China. [331]Department of Clinical Sciences, Psychiatry, Umeå University, Umeå, Sweden. [332]Division of Psychiatry, Department of Mental Health Neuroscience, University College London, London, UK. [333]Department of Psychiatry, San Cecilio University Hospital, University of Granada, Granada, Spain. [334]Institute of Medical Genetics and Pathology, University Hospital Basel, Basel, Switzerland. [335]Department of Biomedicine, University of Basel, Basel, Switzerland. [336]Institute of Neuroscience and Medicine (INM-1), Research Center Juelich, Juelich, Germany. [337]Eli Lilly and Company, Surrey, UK. [338]Neuropsychiatric Genetics Research Group, Department of Psychiatry, Trinity College Dublin, Dublin, Ireland. [339]UCL Genetics Institute, UCL, London, UK. [340]Centre for Psychiatry, Queen Mary University London, London, UK. [341]Department of Cellular, Computational and Integrative Biology, University of Trento, Trento, Italy. [342]Dementia Research Institute, Cardiff University, Cardiff, UK. [343]MRC Centre for Neuropsychiatric Genetics and Genomics, Cardiff University, Cardiff, UK. [344]Department of Psychiatry, Phoenix VA Healthcare System, Phoenix AZ, USA. [345]Banner-University Medical Center, Phoenix AZ, USA. [346]Department of Human Molecular Genetics of the Institute of

Biochemistry and Genetics of the Ufa Federal Research Center of the Russian Academy of Sciences (IBG UFRC RAS), Ufa, Russia. [347]Federal State Educational Institution of Highest Education Bashkir State Medical University of Public Health Ministry of Russian Federation (BSMU), Ufa, Russia. [348]Psychiatric Genetic Epidemiology and Neurobiology Laboratory (PsychGENe lab), Department of Psychiatry and Behavioral Sciences, SUNY Upstate Medical University, Syracuse NY, USA. [349]Department of Psychiatry, Sungkyunkwan University School of Medicine, Samsung Medical Center, Seoul, Korea. [350]Institute of Biochemistry and Genetics of the Ufa Federal Research Center of the Russian Academy of Sciences (IBG UFRC RAS), Ufa, Russia. [351]Department of Psychiatry and Zilkha Neurogenetics Institute, Keck School of Medicine at University of Southern California, Los Angeles CA, USA. [352]Department of Cell Biology, State University of New York, Downstate Medical Center, New York NY, USA. [353]Department of Psychosis, Institute of Mental Health, Singapore, Singapore. [354]Neuroscience and Mental Health, Lee Kong Chian School of Medicine, Nanyang Technological University, Singapore, Singapore. [355]Institute of Behavioral Science, Feinstein Institutes for Medical Research, Manhasset NY, USA. [356]Department of Psychiatry, Zucker School of Medicine at Hofstra/Northwell, Hempstead NY, USA. [357]Human Genetics, Genome Institute of Singapore, A*STAR, Singapore, Singapore. [358]Department of Medicine, Yong Loo Lin School of Medicine, National University of Singapore, Singapore. [359]Roche Pharma Research and Early Development, Roche Innovation Center Basel, F. Hoffman-La Roche Ltd, Basel, Switzerland. [360]Department of Preventative Medicine, Faculdade de Medicina FMUSP, University of Sao Paulo, Sao Paulo SP, Brazil. [361]Department of Psychosis Studies, Institute of Psychiatry, King's College London, London, UK. [362]Department of Psychiatry, University of Pittsburgh, Pittsburgh PA, USA. [363]Center for Neurobehavioral Genetics, Semel Institute for Neuroscience and Human Behavior, University of California, Los Angeles CA, USA. [364]Department of Psychiatry, Erasmus University Medical Center, Rotterdam, The Netherlands. [365]Early Clinical Development, Pfizer Worldwide Research and Development, Groton CT, USA. [366]Analytic and Translational Genetics Unit, Department of Medicine, Department of Neurology and Department of Psychiatry Massachusetts General Hospital, Boston MA, USA. [367]Stanley Center for Psychiatric Research, Broad Institute of MIT and Harvard, Cambridge, USA. [368]Shanghai Key Laboratory of Psychotic Disorders, Shanghai Mental Health Center, Shanghai Jiao Tong University School of Medicine, Shanghai, China. [369]Department of Biochemistry and Molecular Biology II, Faculty of Pharmacy, University of Granada, Granada, Spain. [370]Institute of Neurosciences, Biomedical Research Center (CIBM), University of Granada, Granada, Spain. [371]Faculty of Science, Medicine and Health, School of Chemistry and Molecular Bioscience, University of Wollongong, Wollongong NSW, Australia. [372]Illawarra Health and Medical Research Institute, Wollongong NSW, Australia. [373]Department of Biomedical and Neuromotor Sciences, University of Bologna, Bologna, Italy. [374]Centre for PanorOmic Sciences, LKS Faculty of Medicine, The University of Hong Kong, Hong Kong, China. [375]State Key Laboratory of Brain and Cognitive Sciences, LKS Faculty of Medicine, The University of Hong Kong, Hong Kong, China. [376]Department of Psychiatry, LKS Faculty of Medicine, The University of Hong Kong, Hong Kong, China. [377]Affiliated Hospital of Qingdao University and Biomedical Sciences Institute of Qingdao University (Qingdao Branch of SJTU Bio-X Institutes), Qingdao University, Qingdao, China. [378]Institute of Medical Sciences, University of Aberdeen, Aberdeen, UK. [379]Center for Behavioral Genomics; Department of Psychiatry; University of California, San Diego; La Jolla CA, USA. [380]Institute of Genomic

Medicine, University of California, San Diego; La Jolla CA, USA. [381]University Medical Center Utrecht, Department of Psychiatry, Utrecht, The Netherlands. [382]King's College London, King's Health Partners, Department of Psychosis Studies, Institute of Psychiatry, London, UK. [383]Department of Psychiatry & Human Behavior, School of Medicine, University of California, Irvine CA, USA. [384]Institute of Biological Psychiatry, Mental Health Services, Copenhagen University Hospital, Copenhagen, Denmark. [385]Department of Clinical Medicine, University of Copenhagen, Copenhagen, Denmark. [386]Center for GeoGenetics, GLOBE Institute, University of Copenhagen, Copenhagen, Denmark. [387]The Lundbeck Foundation Initiative for Integrative Psychiatric Research, iPSYCH, Copenhagen, Denmark. [388]School of Psychiatry and Clinical Neurosciences, The University of Western Australia, Perth WA, Australia. [389]Peking University Sixth Hospital, Peking University Institute of Mental Health, Beijing, China. [390]National Clinical Research Center for Mental Disorders & NHC Key Laboratory of Mental Health (Peking University) & Chinese Academy of Medical Sciences Research Unit (No.2018RU006), Beijing, China. [391]PKU-IDG/McGovern Institute for Brain Research, Peking University, Beijing, China. [392]Mental Illness Research, Education, and Clinical Center (VISN 2 South), James J. Peters VA Medical Center, New York NY, USA. [393]Oxford Health NHS Foundation Trust, Oxford, UK. [394]Department of Clinical Genetics, Center for Neurogenomics and Cognitive Research, University Medical Center Amsterdam, Amsterdam, The Netherlands. [395]Department of Functional Genomics, Faculty of Exact Science, Center for Neurogenomics and Cognitive Research, VU University Amsterdam and VU Medical Center, Amsterdam, The Netherlands. [396]Westlake Laboratory of Life Sciences and Biomedicine, Hangzhou Zhejiang, China.

[*]Lists of authors and their affiliations appears at the end of the paper.

[†]These authors contributed equally to this work.

[‡]These authors jointly supervised this work.

#### Indonesia Schizophrenia Consortium

Nan Dai[397,398], Qin Wenwen[397,398], DB Wildenauer[397,398], Feranindhya Agiananda[399], Nurmiati Amir[399], Ronald Antoni[399], Tiana Arsianti[399], Asmarahadi Asmarahadi[399], H Diatri[399], Prianto Djatmiko[399], Irmansyah Irmansyah[399], Siti Khalimah[399], Irmia Kusumadewi[399], Profitasari Kusumaningrum[399], Petrin R Lukman[399], Martina W Nasrun[399], NS Safyuni[399], Prasetyawan Prasetyawan[399], G Semen[399], Kristiana Siste[399], Heriani Tobing[399], Natalia Widiasih[399], Tjhin Wiguna[399], D Wulandari[399], None Evalina[399], AJ Hananto[399], Joni H Ismoyo[399], TM Marini[399], Supiyani Henuhili[399], Muhammad Reza[399], Suzy Yusnadewi[399]

[397]Western Australian Institute for Medical Research and Centre for Medical Research, University of Western Australia, Nedlands WA, Australia. [398]School of Psychiatry and Clinical Neurosciences, University of Western Australia, Crawley WA, Australia. [399]Department of Psychiatry, University of Indonesia, Jakarta, Indonesia.

#### PsychENCODE

Alexej Abyzov[400], Schahram Akbarian[401], Allison Ashley-Koch[402], Harm van Bakel[401], Michael Breen[401], Miguel Brown[403], Julien Bryois[404], Becky Carlyle[405], Alex Charney[401], Gerard Coetzee[406], Gregory Crawford[402], Stella Dracheva[401], Prashant Emani[405], Peggy Farnham[406], Menachem Fromer[143], Timur Galeev[405], Mike Gandal[407], Mark Gerstein[405], Gina Giase[408], Kiran Girdhar[401], Fernando Goes[409], Kay Grennan[403], Mengting Gu[405], Brittney Guerra[405], Gamze Gursoy[405], Gabriel Hoffman[401], Thomas Hyde[267], Andrew Jaffe[267], Shan Jiang[408], Yan Jiang[401], Amira Kefi[408], Yunjung Kim[410], Robert Kitchen[405], James Knowles[411], Fides Lay[406], Donghoon Lee[405], Mingfeng Li[405], Chunyu Liu[243], Shuang Liu[405], Eugenio Mattei[412], Fabio Navarro[405], Xinghua Pan[405], Mette A Peters[413], Dalila Pinto[401], Sirisha Pochareddy[405], Damon Polioudakis[407], Michael Purcaro[412], Shaun Purcell[401], Henry Pratt[412], Tim Reddy[402], Suhn Rhie[406], Panagiotis Roussos[401], Joel Rozowsky[405], Stephan Sanders[414], Nenad Sestan[405], Anurag Sethi[405], Xu Shi[405], Annie Shieh[408], Vivek Swarup[407], Anna Szekely[405], Daifeng Wang[405], Jonathan Warrell[405], Sherman Weissman[405], Zhiping Weng[415], Kevin White[416], Jennifer Wiseman[401], Heather Witt[406], Hyejung Won[407], Shannon Wood[406], Feinan Wu[405], Xuming Xu[405], Lijing Yao[406], Peter Zandi[409]

[400]Mayo Clinic, Rochester MN, USA. [401]Mount Sinai, New York NY, USA. [402]Duke University, Durham NC, USA. [403]University of Chicago, Chicago IL, USA. [404]Karolinska Institutet, Stockholm, Sweden. [405]Yale University, New Haven CT, USA. [406]University of Southern California, Los Angeles CA, USA. [407]University of California, Los Angeles CA, USA. [408]University of Illinois at Chicago, Chicago IL, USA. [409]Johns Hopkins University, Baltimore MD, USA. [410]University of North Carolina - Chapel Hill, Chapel Hill NC, USA. [411]SUNY Downstate Medical Center, New York NY, USA. [412]University of Massachusetts, Amherst MA, USA. [413]Sage Bionetworks, Seattle WA, USA. [414]University of California, San Francisco CA, USA. [415]University of Massachusetts Medical School, Worcester MA, USA. [416]Yong Loo Lin School of Medicine, National University of Singapore, Singapore.

#### Psychosis Endophenotypes International Consortium

Maria J Arranz[417,418], Steven Bakker[178], Stephan Bender[419,420], Elvira Bramon[417,421], David A Collier[88,337], Benedicto Crepo-Facorro[110,422], Jeremy Hall[423], Conrad Iyegbe[417], Assen V Jablensky[288], René Kahn[178], Stephen Lawrie[424], Cathryn Lewis[417], Kuang Lin[417], Don H Linszen[425], Ignacio Mata[110,422], Andrew McIntosh[424], Robin M Murray[417], Roel A Ophoff[426], Jim van Os[427,428], John Powell[417], Dan Rujescu[429,430], Muriel Walshe[417], Matthias Weisbrod[420]

[417]King's College London, London, UK. [418]Fundació de Docència i Recerca Mútua de Terrassa, Universitat de Barcelona, Barcelona, Spain. [419]Child and Adolescent Psychiatry, University of Technology Dresden, Dresden, Germany. [420]Section for Experimental Psychopathology, General Psychiatry, Heidelberg, Germany. [421]Institute of Cognitive Neuroscience, University College London, London, UK. [422]University Hospital Marqués de Valdecilla, Instituto de Formación e Investigación Marqués de Valdecilla, University of Cantabria, Santander, Spain. [423]Neuroscience and Mental Health Research Institute, Division of Psychiatry and Clinical Neuroscience, Cardiff University, Cardiff, UK. [424]Division of Psychiatry, University of Edinburgh, Edinburgh, UK. [425]Department of Psychiatry, Academic Medical Center, University of Amsterdam, Amsterdam, The Netherlands. [426]Department of Human Genetics, David Geffen School of Medicine, University of California, Los Angeles CA, USA. [427]Maastricht University Medical Centre, South

Limburg Mental Health Research and Teaching Network, EURON, Maastricht, The Netherlands. [428]Institute of Psychiatry, King's College London, London, UK. [429]Department of Psychiatry, University of Halle, Halle, Germany. [430]Department of Psychiatry, University of Munich, Munich, Germany.

#### The SynGO Consortium

Tilmann Achsel[431], Maria Andres-Alonso[432], Claudia Bagni[431], Àlex Bayés[433], Thomas Biederer[434], Nils Brose[435], Tyler C. Brown[12], John Jia En Chua[436], Marcelo P. Coba[437], L. Niels Cornelisse[438], Arthur P.H. de Jong[439], Jaime de Juan-Sanz[440], Daniela C. Dieterich[441,442], Guoping Feng[12,443], Hana L. Goldschmidt[444], Eckart D. Gundelfinger[442], Casper Hoogenraad[445], Richard L. Huganir[444], Steven E. Hyman[12,446], Cordelia Imig[447], Reinhard Jahn[448], Hwajin Jung[449], Pascal S. Kaeser[450], Eunjoon Kim[449], Frank Koopmans[395], Michael R. Kreutz[432], Noa Lipstein[451], Harold D. MacGillavry[439], Robert Malenka[452], Peter S. McPherson[453], Vincent O'Connor[454], Rainer Pielot[441,442], Timothy A. Ryan[455], Dnyanada Sahasrabudhe[395], Carlo Sala[456], Morgan Sheng[12], Karl-Heinz Smalla[441,442], August B Smit[457], Thomas C. Südhof[458], Paul D. Thomas[459], Ruud F. Toonen[395], Jan R.T. van Weering[438], Matthijs Verhage [395], Chiara Verpelli[456]

[431]Department of Fundamental Neurosciences, University of Lausanne, Lausanne, Switzerland. [432]RG Neuroplasticity, Leibniz Institute for Neurobiology, Magdeburg, Germany. [433]Molecular Physiology of the Synapse Laboratory, Biomedical Research Institute Sant Pau, Barcelona, Spain. [434]Department of Neurology, Yale School of Medicine, New Haven CT, USA. [435]Department of Molecular Neurobiology, Max Planck Institute of Experimental Medicine, Göttingen, Germany. [436]LSI Neurobiology Programme, National University of Singapore, Singapore. [437]Zilkha Neurogenetic Institute and Department of Psychiatry and Behavioral Sciences, Keck School of Medicine, University of Southern California, Los Angeles CA, USA. [438]Functional Genomics section, Department Human Genetics, Center for Neurogenomics and Cognitive Research, Amsterdam University Medical Center, Amsterdam, The Netherlands. [439]Cell Biology, Neurobiology and Biophysics, Department of Biology, Faculty of Science, Utrecht University, Utrecht, The Netherlands. [440]Sorbonne Université, Institut du Cerveau - Paris Brain Institute - ICM, Inserm, CNRS, APHP, Hôpital de la Pitié Salpêtrière, Paris, France. [441]Institute for Pharmacology and Toxicology, Medical Faculty Otto-von-Guericke University Magdeburg, Magdeburg, Germany. [442]Leibniz Institute for Neurobiology (LIN), Magdeburg, Germany. [443]McGovern Institute for Brain Research, Department of Brain and Cognitive Sciences, Massachusetts Institute of Technology (MIT), Cambridge MA, USA. [444]Solomon H. Snyder Department of Neuroscience Kavli Neuroscience Discovery Institute The Johns Hopkins University School of Medicine Baltimore, Baltimore MD, USA. [445]Department of Neuroscience, Genentech, Inc., South San Francisco CA, USA. [446]Department of Stem Cell and Regenerative Biology, Harvard University; 7 Divinity Avenue, Cambridge MA 02138. [447]Department of Neuroscience, University of Copenhagen, Copenhagen, Denmark. [448]Laboratory of Neurobiology, Max-Planck Institute for Biophysical Chemistry, Göttingen, Germany. [449]Center for Synaptic Brain Dysfunctions, Institute for Basic Science (IBS), Department of Biological Sciences, Korea Advanced Institute of Science and Technology (KAIST), Daejeon, South Korea. [450]Department of Neurobiology, Harvard Medical School, Boston MA, USA. [451]Department of Molecular Physiology and Cell Biology, Leibniz-Forschungsinstitut für Molekulare Pharmakologie, Berlin,

Germany. [452]Nancy Pritzker Laboratory, Department of Psychiatry and Behavioral Sciences, Stanford University, Stanford CA, USA. [453]Department of Neurology and Neurosurgery, Montreal Neurological Institute, McGill University, Montreal, Quebec, Canada. [454]Biological Sciences, University of Southampton, Southampton, UK. [455]Department of Biochemistry, Weill Cornell Medicine, New York NY, USA. [456]CNR Neuroscience Institute, Milan, Italy. [457]Dept. of Molecular and Cellular Neurobiology, Center for Neurogenomics and Cognitive Research, Vrije Universiteit Amsterdam, Amsterdam, The Netherlands. [458]Department of Molecular & Cellular Physiology, Howard Hughes Medical Institute, Stanford University, Stanford CA, USA. [459]Division of Bioinformatics, Dept. of Preventive Medicine, Keck School of Medicine of USC, University of Southern California, Los Angeles CA, USA.

#### Schizophrenia Working Group of the Psychiatric Genomics Consortium

Vassily Trubetskoy[1], Antonio F Pardiñas[2], Georgia Panagiotaropoulou[1], Swapnil Awasthi[1], Tim B Bigdeli[5,6,7], Charlotte A Dennison[2], Lynsey S Hall[2], Max Lam[12,13,14], Oleksandr Frei[16,17,18], Alexander L Richards[2], Jakob Grove[22,23,24], Zhiqiang Li[26,27], Mark Adams[30], Ingrid Agartz[16,31,32], Elizabeth G Atkinson[10,12], Esben Agerbo[22,33], Mariam Al Eissa[34], Margot Albus[35], Madeline Alexander[36], Behrooz Z Alizadeha[37,38], Köksal Alptekin[39,40], Thomas D Als[22,23,24], Farooq Amin[41], Volker Arolt[42], Manuel Arrojo[43], Lavinia Athanasiu[16,17], Maria Helena Azevedo[44], Silviu A Bacanu[45], Nicholas J Bass[34], Martin Begemann[46], Richard A Belliveau[12], Judit Bene[47], Beben Benyamin[48,49,50], Sarah E Bergen[8], Giuseppe Blasi[51], Julio Bobes[52,53,54], Stefano Bonassi[55], Alice Braun[1], Rodrigo Affonseca Bressan[56,57], Evelyn J Bromet[58], Richard Bruggeman[37,59], Peter F Buckley[60], Randy L Buckner[61], Jonas Bybjerg-Grauholm[22,62], Wiepke Cahn[63,64], Murray J Cairns[65,66,67], Monica E Calkins[68], Vaughan J Carr[69,70,71], David Castle[72,73], Stanley V Catts[74,75], Kimberley D Chambert[12], Raymond CK Chan[76,77], Boris Chaumette [78,79], Wei Cheng[80], Eric FC Cheung[81], Siow Ann Chong[13,82], David Cohen[83,84,85], Angèle Consoli[83,84], Quirino Cordeiro[86], Javier Costas[87], Charles Curtis[88,89], Michael Davidson[90], Kenneth L Davis[91], Lieuwe de Haan[92,93], Franziska Degenhardt[94], Lynn E DeLisi[19,95], Ditte Demontis[22,23,24], Faith Dickerson[96], Dimitris Dikeos[97], Timothy Dinan[98,99], Srdjan Djurovic[100,101], Jubao Duan[102,103], Giuseppe Ducci[104], Johan G Eriksson[106,107,108], Lourdes Fañanás[109,110], Stephen V Faraone[111], Alessia Fiorentino[34], Andreas Forstner[94,112], Josef Frank[113], Nelson B Freimer[114,115], Menachem Fromer[116], Alessandra Frustaci[117], Ary Gadelha[56,57], Giulio Genovese[12], Elliot S Gershon[118], Marianna Giannitelli[83,84], Ina Giegling[119], Paola Giusti-Rodríguez[120], Stephanie Godard[121], Jacqueline I Goldstein[10], Javier González Peñas[110,122], Ana González-Pinto[110,123], Srihari Gopal[124], Jacob Gratten[3,125], Michael F Green[126,127], Tiffany A Greenwood[128], Olivier Guillin[129,130,131], Sinan Gülöksüz[132,133], Raquel E Gur[68], Ruben C Gur[68], Blanca Gutiérrez[134], Eric Hahn[135], Hakon Hakonarson[136], Vahram Haroutunian[91,137,138], Annette M Hartmann[119], Carol Harvey[72,139], Caroline Hayward[140], Frans A Henskens[141], Stefan Herms[142], Per Hoffmann[142], Daniel P Howrigan[10,143], Masashi Ikeda[144], Conrad Iyegbe[145], Inge Joa[146], Antonio Julià[147], Anna K Kähler[8], Tony Kam-Thong[148], Yoichiro Kamatani[149,150], Sena Karachanak-Yankova[151,152], Oussama Kebir[78], Matthew C Keller[153], Brian J Kelly[141], Andrey Khrunin[154], Sung-Wan Kim[155], Janis Klovins[156], Nikolay Kondratiev[157], Bettina Konte[119], Julia Kraft[1,158], Michiaki Kubo[159], Vaidutis Kučinskas[160], Zita Ausrele Kučinskiene[160], Agung Kusumawardhani[161], Hana Kuzelova-Ptackova[162], Stefano Landi[163], Laura C Lazzeroni[164,165], Phil H Lee[12,166], Sophie E Legge[2], Douglas S Lehrer[167], Rebecca Lencer[42],

Bernard Lerer[168], Miaoxin Li[169], Jeffrey Lieberman[170], Gregory A Light[128,171], Svetlana Limborska[154], Chih-Min Liu[172,173], Jouko Lönnqvist[174,175], Carmel M Loughland[176], Jan Lubinski[177], Jurjen J Luykx[132,178,179,180], Amy Lynham[2], Milan Macek Jr[181], Andrew Mackinnon[182,183], Patrik KE Magnusson[8], Brion S Maher[184], Wolfgang Maier[185], Dolores Malaspina[91,186], Jacques Mallet[187], Stephen R Marder[188], Sara Marsal[147], Alicia R Martin[10,12,189], Lourdes Martorell[190], Manuel Mattheisen[23,191,192,193], Robert W McCarley[194,195], Colm McDonald[196], John J McGrath[33,197,198], Helena Medeiros[199,200], Sandra Meier[191,201], Bela Melegh[202], Ingrid Melle[16,17], Raquelle I Mesholam-Gately[19,203], Andres Metspalu[204], Patricia T Michie[205], Lili Milani[204], Vihra Milanova[206], Marina Mitjans[46], Espen Molden[207,208], Esther Molina[209], María Dolores Molto[110,210,211], Valeria Mondelli[89,212], Carmen Moreno[110,122], Christopher P Morley[213], Gerard Muntané[190,214], Kieran C Murphy[215], Inez Myin-Germeys[216], Igor Nenadić[217,218], Gerald Nestadt[219], Liene Nikitina-Zake[156], Cristiano Noto[56,57], Keith H Nuechterlein[126], Niamh Louise O'Brien[34], F Anthony O'Neill[220], Sang-Yun Oh[221,222], Ann Olincy[223], Vanessa Kiyomi Ota[57,224], Christos Pantelis[139,225,226,227], George N Papadimitriou[97], Mara Parellada[110,122], Tiina Paunio[228,229], Renata Pellegrino[136], Sathish Periyasamy[197,230], Diana O Perkins[231], Bruno Pfuhlmann[232], Olli Pietiläinen[12,233,234], Jonathan Pimm[34], David Porteous[235], John Powell[236], Diego Quattrone[88,89,237], Digby Quested[238,239], Allen D Radant[240,241], Antonio Rampino[51], Mark H Rapaport[242], Anna Rautanen[148], Abraham Reichenberg[91], Cheryl Roe[243], Joshua L Roffman[244], Julian Roth[245], Matthias Rothermundt[42], Bart PF Rutten[132], Safaa Saker-Delye[246], Veikko Salomaa[247], Julio Sanjuan[110,211,248], Marcos Leite Santoro[57,224], Adam Savitz[124], Ulrich Schall[66,249], Rodney J Scott[65,250,251], Larry J Seidman[19,203], Sally Isabel Sharp[34], Jianxin Shi[252], Larry J Siever[91,253], Kang Sim[256,257,258], Nora Skarabis[1], Petr Slominsky[154], Hon-Cheong So[259,260], Janet L Sobell[199], Erik Söderman[32], Helen J Stain[261,262], Nils Eiel Steen[17,263], Agnes A. Steixner-Kumar[46], Elisabeth Stögmann[264], William S Stone[265,266], Richard E Straub[267], Fabian Streit[113], Eric Strengman[268], T Scott Stroup[170], Mythily Subramaniam[13,82], Catherine A Sugar[126,269], Jaana Suvisaari[247], Dragan M Svrakic[270], Neal R Swerdlow[128], Jin P Szatkiewicz[120], Thi Minh Tam Ta[271,272], Atsushi Takahashi[132,273], Chikashi Terao[273], Florence Thibaut[274,275], Draga Toncheva[151,276], Paul A Tooney[65,66,67], Silvia Torretta[51], Sarah Tosato[277], Gian Battista Tura[278], Bruce I Turetsky[68], Alp Üçok[279], Arne Vaaler[280,281], Therese van Amelsvoort[89,132], Ruud van Winkel[132,282], Juha Veijola[283,284], John Waddington[285], Henrik Walter[286], Anna Waterreus[287,288], Bradley T Webb[45], Mark Weiser[289], Nigel M Williams[2], Stephanie H Witt[113], Brandon K Wormley[45], Jing Qin Wu[290], Zhida Xu[291], Robert Yolken[292], Clement C Zai[293,294], Wei Zhou[27], Feng Zhu[295,296], Fritz Zimprich[264], Eşref Cem Atbaşoğlu[186,297], Muhammad Ayub[298], Alessandro Bertolino[51], Donald W Black[299], Nicholas J Bray[2], Gerome Breen[88], Nancy G Buccola[300], William F Byerley[301], Wei J Chen[302,303], C Robert Cloninger[270], Benedicto Crespo-Facorro[304,305], Gary Donohoe[196], Robert Freedman[223], Cherrie Galletly[306,307,308], Massimo Gennarelli[309,310], David M Hougaard[22,62], Hai-Gwo Hwu[173,311], Assen V Jablensky[288], Steven A McCarroll[12], Jennifer L Moran[12,244], Ole Mors[22,312], Preben B Mortensen[22,33], Bertram Müller-Myhsok[313,314,315], Amanda L Neil[316], Merete Nordentoft[22,317], Michele T Pato[318,319], Tracey L Petryshen[166], Ann E Pulver[219], Thomas G Schulze[193,322,323,324], Jeremy M Silverman[91,253], Jordan W Smoller[12,166], Eli A Stahl[116,325,326], Debby W Tsuang[240,241], Elisabet Vilella[190], Shi-Heng Wang[327], Shuhua Xu[328,329,330], Rolf Adolfsson[331], Celso Arango[110,122], Bernhard T Baune[42,226,227], Sintia Iole Belangero[57,224], Anders D Børglum[22,23,24], David Braff[128,171], Elvira Bramon[332], Joseph D Buxbaum[91], Dominique Campion[129,130], Jorge A Cervilla[333], Sven Cichon[334,335,336], David A Collier[337], Aiden Corvin[338],

Marta Di Forti[88,89,237], Enrico Domenici[341], Hannelore Ehrenreich[46], Valentina Escott-Price[342,343], Tõnu Esko[204,325], Ayman H Fanous[7,344,345], Anna Gareeva[346,347], Micha Gawlik[245], Pablo V Gejman[102,103], Michael Gill[338], Stephen J Glatt[348], Vera Golimbet[157], Kyung Sue Hong[349], Christina M Hultman[8], Steven E Hyman[12,233], Nakao Iwata[144], Erik G Jönsson[32,263], René S Kahn[63,91], James L Kennedy[293,294], Elza Khusnutdinova[347,350], George Kirov[2], James A Knowles[351,352], Marie-Odile Krebs[78], Claudine Laurent-Levinson[83,84], Jimmy Lee[353,354], Todd Lencz[14,355,356], Douglas F Levinson[164], Qingqin S Li[124], Jianjun Liu[357,358], Anil K Malhotra[14,355,356], Dheeraj Malhotra[359], Andrew McIntosh[30], Andrew McQuillin[34], Paulo R Menezes[360], Vera A Morgan[287,288], Derek W Morris[196], Bryan J Mowry[197,230], Robin M Murray[89,361], Vishwajit Nimgaonkar[362], Markus M Nöthen[94], Roel A Ophoff[114,363,364], Sara A Paciga[365], Aarno Palotie[234,366,367], Carlos N Pato[318,319], Shengying Qin[27,368], Marcella Rietschel[113], Brien P Riley[45], Margarita Rivera[369,370], Dan Rujescu[119], Meram C Saka[297], Alan R Sanders[102,103], Sibylle G Schwab[371,372], Alessandro Serretti[373], Pak C Sham[374,375,376], Yongyong Shi[27,377], David St Clair[378], Ming T Tsuang[379,380], Jim van Os[381,382], Marquis P Vawter[383], Daniel R Weinberger[267], Thomas Werge[384,385,386,387], Dieter B Wildenauer[388], Xin Yu[389,390], Weihua Yue[389,390,391], Peter A Holmans[2], Panos Roussos[28,392], Evangelos Vassos[88,89,393], Danielle Posthuma[395], Ole A Andreassen[16,17], Kenneth S Kendler[45], Michael J Owen[2], Naomi R Wray[3,197], Mark J Daly[10,143,234], Hailiang Huang[10,12,189], Benjamin M Neale[10,12], Patrick F Sullivan[8,120,231], Stephan Ripke[1,10,367], James TR Walters[2], Michael C O'Donovan[2]
